# Mapping the Middle East Respiratory Syndrome (MERS) related Research – A Scoping Review (2012-2023)

**DOI:** 10.1101/2023.11.08.23298197

**Authors:** Maya Hassan, Halima Yarow, Ruth Mccabe, Sophie Von Dobschuetz, Wasiq Khan, Amal Barakat, Maria D. Van Kerkhove, Abdinasir Abu Bakar, Hala Abou El Naja

## Abstract

**Background:** Middle East respiratory syndrome (MERS), is a zoonotic disease caused by MERS coronavirus (MERS-CoV). The purpose of this scoping review was to take stock of the empirical research evidence for MERSDCoV, map the information to priority research areas as set out in existing MERS-CoV research roadmaps, identify technical areas that received less attention and set recommendations for the advancement of MERS-CoV research.

**Methods:** We undertook a scoping review for MERS-CoV, comprehensively searching the three databases PubMed, EMBASE, and CINAHL for studies published between 1 January 2012 and 24 January 2023. Two reviewers screened studies and extracted data using a pilot-tested screening form. We categorized studies into priority research areas outlined in existing roadmaps and summarized the evidence available for each category.

**Results:** A total of 1,264 records were included in the review, assigned into pre-defined categories. 33% of the included records were molecular genetics studies, followed by therapeutic studies (17.6%) and pathogenesis studies (15.6%). We found that, while there has been a substantial research effort on MERS-CoV, many technical themes pertaining to the areas of animal, human, animal-human interface, and environmental research identified by FAO, WHO, and WOAH in the past have not sufficiently been addressed to date. This includes asymptomatic human cases role in transmission, human exposure risk from dromedary products, reinfection, analyses of camel value chain and production systems, and anthropological studies characterizing interactions at the animal-human interface, in addition to studies highlighting the role of environmental factors in MERS-CoV transmission.

**Conclusion:** Our study highlights the continued need for coordinated action to better prepare for, prevent, detect, and respond to MERS-CoV. Examples include the need for enhancing collaborative surveillance, accelerating the development of MERS-CoV medical countermeasures, strengthening community protection, reducing MERS-CoV transmission at healthcare facility level and reinforcing multi-sectoral coordination using the One Health approach.

## Background

In mid-2012, a novel coronavirus strain was isolated from a Saudi Arabian patient with acute pneumonia and renal failure [1]. The virus, later named Middle East respiratory syndrome coronavirus (MERS-CoV), is a zoonotic coronavirus that can be transmitted to humans from infected dromedary camels, its reservoir host [2, 3, 4]. To date, limited onwards human-to-human transmission has been observed in healthcare settings and, to a lesser extent, communities [7]. MERS-CoV appears to be circulating widely in dromedary camel populations throughout the Middle East, Africa, and South Asia and is associated with asymptomatic infection or mild upper respiratory signs in dromedaries [5]. Although there is no perceived impact of MERS-CoV on dromedary populations, human infection is a major public health concern [5]. As per a WHO report from May 2023, MERS-CoV has been reported from 27 countries, in the Middle East, North Africa, Europe, the United States of America (USA), and Asia since its emergence in 2012, with 2,604 laboratory-confirmed human cases and 936 associated deaths globally, resulting in a case-fatality ratio (CFR) of 36% [6]. However, the infection fatality ratio is thought to be less as mild MERS-CoV cases may be missed by surveillance systems [6]. Notably, the highest numbers of human cases and deaths have been identified and reported by the Kingdom of Saudi Arabia (KSA), with 2,196 cases and 855 related deaths as of May 2023 (CFR: 39%) [6]. Although there has been no documented sustained human-to-human transmission outside of healthcare settings, MERS-CoV remains a significant public health concern due to its pandemic potential and high CFR. This, and the fact that no licensed therapeutics or vaccines are available [7], resulted in the inclusion of MERS-CoV in WHO’s list of pathogens with epidemic potential, prioritized for research and development in emergency contexts [8].

The number of MERS cases reported to WHO has declined substantially since the beginning of the COVID-19 pandemic. There are several non-exclusive hypotheses which may explain this, including reduced MERS-CoV testing as epidemiological surveillance focused on SARS-CoV-2, improved public health and social measures (PHSM) implemented to control the spread of SARS-CoV-2 also reducing opportunities for MERS-CoV to infect and spread, and possible cross-protective immunity from SARS-CoV-2 or MERS-CoV infection or SARS-CoV-2 vaccination [9]. While attention to MERS-CoV declined during the COVID-19 public health emergency of international concern (PHEIC), 8 years of preparedness and response activities for MERS-CoV globally provided a critical foundation for the early COVID-19 response when technical guidance documents, including for surveillance, case investigation, infection prevention and control (IPC), and clinical care, were quickly adapted and existing laboratory and expert networks and mathematical models immediately utilized [9]. The number of cases of other respiratory diseases, such as MERS, may increase again as public health and social measures are lifted after the COVID-19 global PHEIC was lifted as there is the risk of a reduction in the use of standard and enhanced IPC measures in healthcare settings [10]. This highlights the importance of and urgent need sustain gains made during COVID-19 for re-strengthening prevention, preparedness, readiness, and response efforts and bridging the gap in MERS-CoV research.

Since the emergence of MERS-CoV in 2012, global stakeholders such as WHO, the Food and Agriculture Organization (FAO), and the World Organization for Animal Health (WOAH) have been advocating for research in human and animal populations and at the animal-human interface. To highlight key research areas, identify gaps and measure progress towards addressing them, the FAO-WHO-WOAH tripartite has developed and continuously updated roadmaps for research and development (R&D) to prevent and mitigate the impact of MERS. These include a roadmap for MERS diagnostics, vaccines, and therapeutics in 2015, a roadmap for MERS diagnostics in 2018, and a broader roadmap for the public health research agenda on MERS in 2018 [11, 12]. FAO, WHO, and WOAH regularly conduct global technical meetings on MERS-CoV to follow up on progress against the research agendas [18, 19]. Over the past 11 years, researchers have published many reviews to describe major advances in understanding MERS-CoV in human and animal populations and to address FAO-WHO-WOAH roadmap research priorities [13, 14]. However, there has been no formal comprehensive mapping of MERS-CoV studies to date. To take stock of the empirical research evidence for MERS]CoV in humans and animals, map the information to priority research areas as set out in existing MERS-CoV research roadmaps such as from the FAO-WHO-WOAH technical meeting held in Geneva on 25-27 September 2017 (see Table 1) [18], identify MERS-CoV-related technical areas tha received less attention and develop recommendations for the advancement in MERS-CoV research, we undertook a scoping review of the MERS]CoV literature published since 2012 [17].

**Table 1:**
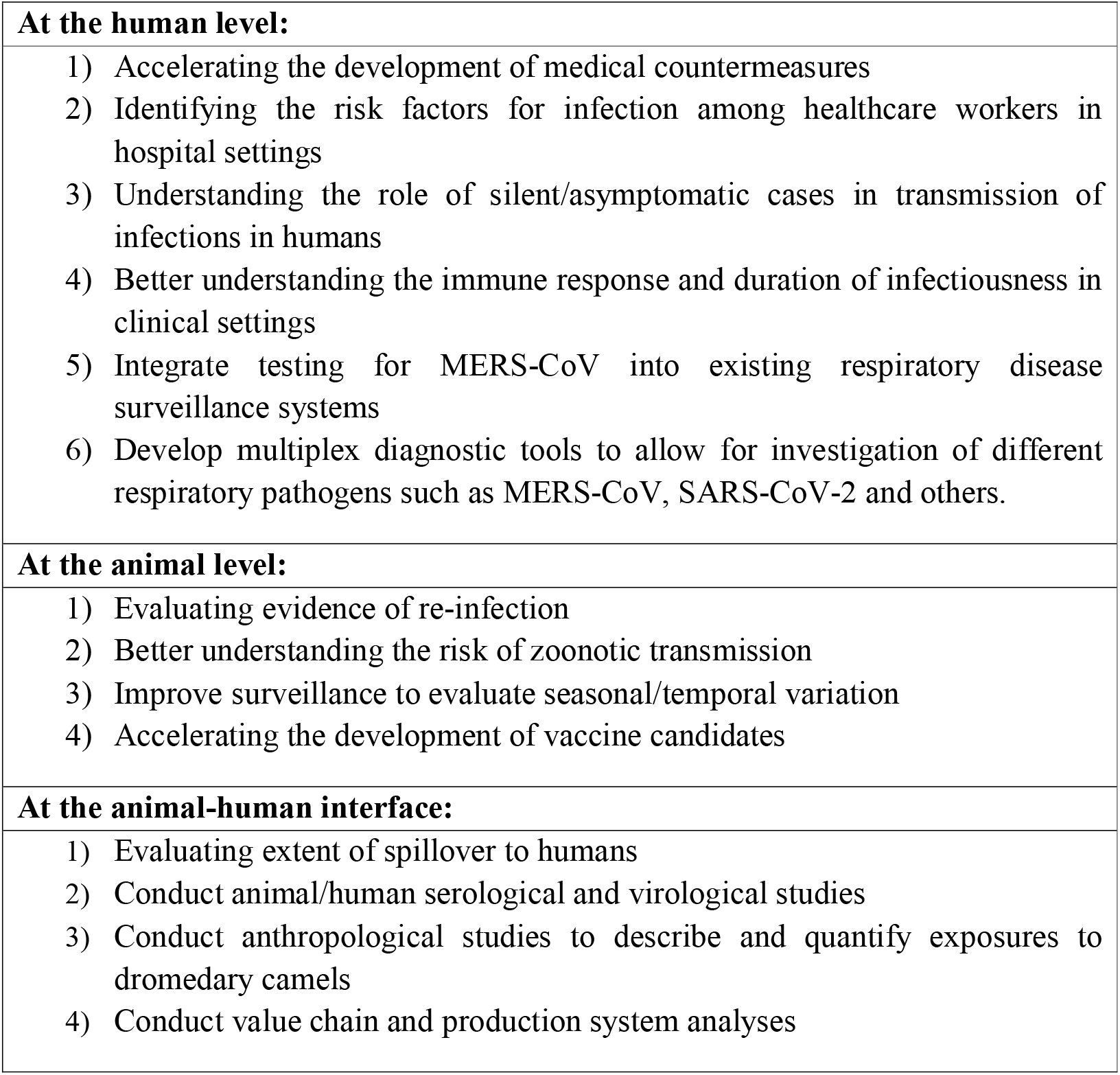
Research priorities set by FAO-WHO-WOAH technical meeting held in Geneva on 25-27 September 2017.

|  |
| --- |
| **At the human level:** |
| 1) Accelerating the development of medical countermeasures<br>2) Identifying the risk factors for infection among healthcare workers in hospital settings<br>3) Understanding the role of silent/asymptomatic cases in transmission of infections in humans<br>4) Better understanding the immune response and duration of infectiousness in clinical settings<br>5) Integrate testing for MERS-CoV into existing respiratory disease surveillance systems<br>6) Develop multiplex diagnostic tools to allow for investigation of different respiratory pathogens such as MERS-CoV, SARS-CoV-2 and others. |
| **At the animal level:** |
| 1) Evaluating evidence of re-infection<br>2) Better understanding the risk of zoonotic transmission<br>3) Improve surveillance to evaluate seasonal/temporal variation<br>4) Accelerating the development of vaccine candidates |
| **At the animal-human interface:** |
| 1) Evaluating extent of spillover to humans<br>2) Conduct animal/human serological and virological studies<br>3) Conduct anthropological studies to describe and quantify exposures to dromedary camels<br>4) Conduct value chain and production system analyses |

## Methodology

We conducted a scoping review on MERS-CoV research related to animals and humans since 2012. A scoping review generates an overview of available evidence for a given topic, irrespective of data quality, and is an excellent tool to convey the depth of a body of literature [15, 16, 20]. In this review, we used an “evidence maps” design to comprehensively search a broad topic to identify knowledge gaps, research priorities, and needs while presenting results in a user-friendly format, using visual graphs, figures and a searchable database [17, 21]. The results of evidence mapping are used to inform ongoing discussions related to research and funding priorities organized by WHO [21].

The scoping review used the Arksey and O’Malley framework [22], comprised of the following six steps: 1) identifying the research question, 2) searching for relevant studies, 3) selecting studies, 4) charting the data, 5) collating, summarizing, and reporting the results, and 6) consulting with stakeholders to inform and validate study findings [15]. We followed the standard methodology as outlined in the Preferred Reporting Items for Systematic Reviews and Meta-Analyses extension for Scoping Reviews (PRISMA-ScR) (see online Supplemental file 1) [22].

### Eligibility criteria

► **Type of study:** We included all peer-reviewed studies, except for seminars, protocols, commentaries, editorials, correspondences, letters to editors, news, viewpoints, guidelines, abstracts, and reviews. The reason for excluding reviews is to focus on primary research areas only.
► **Scope:** We included studies related to MERS-CoV research in animals and humans in all health-related areas.
► **Setting:** We included studies from any geographical setting.
► **Language:** We only included studies written in English. Nonetheless, we also documented the number of studies reported in other languages.
► **Date:** We included studies published between 1 January 2012 and 24 January 2023.

### Search strategy

We searched PubMed, EMBASE, and CINAHL electronic databases, from 1 January 2012 until 24 January 2023. We developed the search strategy with the assistance of an experienced information specialist and based on the Peer Review of Electronic Search Strategies (PRESS) guidelines [23]. The search combined terms for MERS-CoV and included both subject headings and keywords. We restricted the search to English language studies. The detailed search strategy is provided in the online Supplemental file 2.

### Study selection

We completed the selection process in two subsequent stages:

► **Title and abstract screening:** Two reviewers used the eligibility criteria to screen titles and abstracts of identified citations. Full texts judged as eligible by at least one of the two reviewers were then extracted.
► **Full-text screening:** The same two reviewers screened the full texts for eligibility. Any disagreement was resolved by discussion or consultation with a third reviewer. Before selecting eligible publications, an extraction calibration exercise using a random sample of 20 citations was conducted to minimize between-reviewer bias, therefore improving selection process validity.

### Data extraction

Two reviewers extracted data from eligible records using a standardized and pilot-tested screening form developed by the research team. Any disagreement was resolved, similarly as in screening phase, by discussion or consultation with a third reviewer. A first extraction round using a randomly selected sample of 20 citations was conducted by both reviewers in parallel to ensure consistency of data extracted across reviewers. Information extracted from each study included:

► **General characteristics:** paper title, first-author name; journal; date of publishing; first-author institution; first-author institution country; funding agency; study design; scope level (animal, human, animal-human interface or environment); study settings; human populations; and animal populations.
► **Study themes/outcomes:** a team of four researchers established an exhaustive list of all study themes that may be found in MERS-CoV literature, informed by the research priorities set by the FAO-WHO-WOAH technical meeting that was held in Geneva on 25-27 September 2017 (see Table 1) [18]. The themes are: molecular genetics; laboratory diagnostics; seroprevalence; surveillance systems; outbreak investigation; transmissibility; spillover; therapeutics (drug-related or others); vaccinology and immunization; pathogenesis; comorbidities; anthropology and social behavior; outbreak preparedness and response; infection prevention and control (IPC); impact on health system\burden of disease (BoD) and economic impact; case management; mathematical models for MERS; MERS-CoV and SARS-CoV-2 cross-reactive immunity; silent/asymptomatic; and One Health approach.

### Data analysis

We conducted a descriptive analysis of study characteristics according to pre-defined themes to examine the existing evidence and identify neglected MERS-CoV-related technical areas. We conducted this scoping review following the PRISMA-ScR Checklist [22] (**see** online Supplemental file 1).

## Results

### Search results

A total of 6,690 records were identified through the search, 2,865 of which were excluded as duplicates. Title and abstract screening removed 2,188 records for being unrelated to MERS-CoV. 91 records were excluded because only abstracts were available. Full texts of the 1,546 remaining records were retrieved, 282 of which were excluded at the full-text screening phase for the following reasons: duplication (n=6); not related to MERS-CoV (n=35); study type not of interest (n=198); and records in languages other than English (n=43). Finally, a total of 1,264 records were included in the review. **Figure 1** depicts the study flow diagram that summarizes the selection process, following PRISMA reporting guidelines [22].

**Figure 1:** PRISMA: Flowchart of the selection process for the scoping review of MERS-CoV studies and results, (1 January 2012 to 24 January 2023)

### Study characteristics

#### Sources of studies

General characteristics of the studies included in the scoping review are listed in Table 2. The results indicate that 369 scientific journals published studies related to MERS-CoV. About 50% of the studies were published in 27 journals, while 342 journals published the remaining studies. The leading journal in publishing MERS-CoV-related research was found to be the Journal of Virology with (5.6%, n=71) of the total records, followed by (5.5%, n=70) published in Emerging Infectious Diseases, (4%, n=50) published in Viruses, (3.3%, n=42) published in PLoS One, with others as shown in Table 2.

**Table 2:** Characteristics of the MERS-CoV-related studies included in the scoping review, (1 January 2012 to 24 January 2023)

| **Characteristics** |  | **Number of records** | **Percentage (%)** |
| --- | --- | --- | --- |
|  | *Journal of Virology* | 71 | 5.6 |
|  | *Emerging Infectious Diseases* | 70 | 5.5 |
|  | *Viruses* | 50 | 4 |
|  | *PLoS One* | 42 | 3.3 |
|  | *Scientific Reports* | 33 | 2.6 |
|  | *Emerging Microbes and Infections* | 31 | 2.5 |
|  | *Journal of Infection and Public Health* | 28 | 2.2 |
|  | *Virology Journal* | 25 | 2 |
|  | *Proceedings of the National Academy of Sciences of the USA* | 22 | 1.7 |
|  | *Eurosurveillance* | 21 | 1.7 |
|  | *International Journal of Infectious Diseases* | 20 | 1.59 |
| **Journal (n = 369)** | *Antiviral Research* | 19 | 1.5 |
|  | *mBio* | 18 | 1.4 |
|  | *International Journal of Environmental Research and Public Health* | 17 | 1.3 |
|  | *Nature Communications* | 17 | 1.3 |
|  | *Clinical Infectious Diseases* | 17 | 1.3 |
|  | *Journal of Infectious Diseases* | 15 | 1.2 |
|  | *BMC Infectious Diseases* | 14 | 1.1 |
|  | *American Journal of Infection Control* | 12 | 1 |
|  | *Journal of Medical Virology* | 12 | 1 |
|  | *Vaccine* | 12 | 1 |
|  | *Frontiers in Microbiology* | 11 | 0.9 |
|  | *PLoS Pathogens* | 11 | 0.9 |
|  | *Transboundary and Emerging Diseases* | 11 | 0.9 |
|  | *Frontiers in Immunology* | 10 | 0.8 |
|  | *Journal of Biological Chemistry* | 10 | 0.8 |
|  | *Journal of General Virology* | 10 | 0.8 |
|  | Other | 635 | 50.2 |
| **First-author institution country (n=59)** | USA | 241 | 19.1 |
|  | KSA | 235 | 18.6 |
|  | China | 182 | 14.4 |
|  | Republic of Korea | 180 | 14.2 |
|  | Germany | 56 | 4.4 |
|  | Netherlands | 38 | 3 |
|  | Japan | 31 | 2.4 |
|  | United Kingdom | 25 | 2 |
|  | Egypt | 21 | 1.7 |
|  | France | 20 | 1.6 |
|  | Australia | 20 | 1.6 |
|  | Taiwan | 19 | 1.5 |
|  | India | 17 | 1.3 |
|  | Spain | 16 | 1.3 |
|  | Canada | 15 | 1.2 |
|  | United Arab Emirates | 14 | 1.1 |
|  | Iran | 14 | 1.1 |
|  | Other | 120 | 9.5 |
| **Publication or posted date** | 2012 | 0 | 0 |
|  | 2013 | 39 | 3.1 |
|  | 2014 | 106 | 8.4 |
|  | 2015 | 114 | 9 |
|  | 2016 | 163 | 12.9 |
|  | 2017 | 134 | 10.6 |
|  | 2018 | 147 | 11.6 |
|  | 2019 | 143 | 11.3 |
|  | 2020 | 150 | 11.9 |
|  | 2021 | 147 | 11.6 |
|  | 2022 | 116 | 9.2 |
|  | 2023 | 5 | 0.4 |
| **Study design** | Lab trials | 721 | 57 |
|  | Cross - sectional | 172 | 13.6 |
|  | Case report & case series | 81 | 6.4 |
|  | Cohort | 78 | 6.2 |
|  | Case study | 46 | 3.6 |
|  | Descriptive survey | 34 | 2.7 |
|  | Mathematical modeling | 36 | 2.8 |
|  | Retrospective chart review (RCR) | 56 | 4.4 |
|  | Other<sup>a</sup> | 40 | 3.2 |
| **Funding** | Received funding | 950 | 75.2 |
|  | Did not receive funding | 149 | 11.7 |
|  | Not reported | 167 | 13.2 |

<sup>a</sup>:Other study designs include: case control, randomized controlled trials, case-crossover analysis, comparative analysis, natural experimental study, and quasi experimental

#### First-author institution country

This scoping review identified that 59 countries published MERS-CoV-related research as first-author institution countries. Among the first-author institution countries, the USA has the highest records of MERS-CoV-related research (19.1%, n=241), followed by KSA (18.6%, n=235), China (14.4%, n=182), Republic of Korea (14.2%, n=180), and other countries captured in Table 2. Figure 2 presents a map of first-author institution countries included in the scoping review. First-authors from 495 institutions have published on MERS-CoV, and the mapping of these institutions is presented in online Supplemental file 3.

**Figure 2:** Map of first-author countries that conducted studies on MERS-CoV, (1 January 2012 to 24 January 2023)

## Publication date

Figure 3 depicts the cumulative number of studies published yearly between 1 January 2012 and 24 January 2023. Of the 1,264 records included in the review, the highest number of papers was published in 2016 (12.9%, n=163). The number of studies published on MERS-CoV-related research did not decline after 2019.

**Figure 3:** Number of MERS-CoV-related studies across study period by publication date, (1 January 2012 to 24 January 2023)

## Design of studies

Table 3 outlines the different designs used for MERS-related studies. The most common study design was laboratory trials (57%, n=721), followed by cross-sectional studies (13.6%, n=172), case reports and case series (6.4%, n=81), and others.

**Table 3:** The frequencies of the MERS-CoV-related studies characteristics, (1 January 2012 to 24 January)

| Characteristics |  | Number of records | Percentage (%) |
| --- | --- | --- | --- |
| **Scope level\*** | Human | 1035 | 82 |
|  | Animal | 130 | 10.3 |
|  | Animal-human interface | 109 | 8.6 |
|  | Environment | 9 | 0.7 |
| **Study settings\*** | Laboratory | 730 | 57.8 |
|  | Health-care facilities | 303 | 24 |
|  | Community | 93 | 7.4 |
|  | Camel herds (farmed/nomadic herds) | 72 | 5.7 |
|  | Abattoirs /Animal markets/ Slaughterhouse | 45 | 3.6 |
|  | Camel events (races/beauty contests) | 6 | 0.5 |
|  | Other<sup>b</sup> | 59 | 4.7 |
| **Study population\*** | **Human\*** | **776** | **66.5** |
|  | Community | 322 | 41.5 |
|  | Healthcare | 223 | 28.7 |
|  | Household | 101 | 13 |
|  | Occupational animal exposure (farm workers/ live animal workers/ abattoir workers/ quarantine workers) | 74 | 9.5 |
|  | Other<sup>c</sup> | 328 | 42.3 |
|  | **Animals\*** | **391** | **33.5** |
|  | Animal models | 214 | 54.7 |
|  | Dromedary camels | 114 | 29.2 |
|  | Bats | 40 | 10.2 |
|  | Other<sup>d</sup> | 40 | 10.2 |

<sup>\*</sup>More than one option applies
<sup>b</sup>: other study settings include studies used online data to draw modelling
<sup>c</sup>: other includes human cell lines for laboratory
<sup>d</sup>: other include pigs, llamas, bactrian camel, horses, sheep, goats, cattle, donkeys, buffalo, bovine, cow, alpacas, and camelids

## Study settings

Table 3 illustrates the settings in which studies took place. (57.8%, n=730) of studies were conducted in a laboratory, followed by healthcare facilities (24%, n=303) Moreover, (7.4%, n=93) of studies were targeted at communities, followed by camel herds (5.7%, n=72), abattoir and animal markets (3.4%, n=43), and camel events including races or beauty contests (0.5%, n=6).

## Scope level

Table 3 observed frequencies of the different study characteristics by scope level. MERS-CoV studies were mostly focused on humans (82%, n=1035), followed by animals (10.3%, n=130), the animal-human interface (8.6%, n=109), and finally, only (0.7%, n=9) of studies were conducted to understand the virus in the environment. Figure 4 shows the number of MERS-related studies by scope level.

**Figure 4:** Number of MERS-CoV-related studies included in the scoping review by scope level, (1 January 2012 to 24 January 2023)

## Study populations

For studies involving humans, (41.5%, n=322) were community studies, (28.7%, n=223) involved healthcare personnel, followed by studies targeting household members (13%, n=101), and lastly studies involving occupational workers at farms, animal markets, abattoirs, or in quarantine stations (9.5%, n=74). Most studies conducted on animals involved animal models (54.7%, n=214). Moreover, (29.2%, n=114) of studies were conducted on dromedary camels, followed by studies involving bats (10.2%, n=40).

## Study themes

Different themes of the studies are characterized in Table 4, and Figure 5. Molecular genetics studies were the most common theme identified, accounting for (33%, n=417) of studies, followed by therapeutics studies (17.6%, n=222), pathogenesis studies (15.6%, n=197), and vaccinology and immunization studies (11%, n=139). Conversely, other themes have not been frequently explored such as One Health approach studies conducted with the aim of optimizing the health of people, animals, and the environment (0.7%, n=9) (see Supplemental file 3).

**Figure 5:** Number of MERS-CoV-related studies included in the scoping review by themes, (1 January 2012 to 24 January 2023)

**Table 4:** Themes of MERS-CoV-related studies included in the scoping review (1 January 2012 to 24 January)

| **Table 4: Themes of MERS-CoV-related studies included in the scoping review (1 January 2012 to 24 January 2023)** |  |  |  |
| --- | --- | --- | --- |
| **Theme** | **Theme Definition** | **Number of studies** | **Percentage (%)** |
| Molecular genetics | Studies that describe the molecular structure and the cellular activities of the virus | 417 | 33 |
| Therapeutics (drug-related or others) | Studies that test drug candidates for the treatment of MERS-CoV infection | 222 | 17.6 |
| Pathogenesis | Studies that investigate the onset of MERS-CoV disease, its progression, and maintenance | 197 | 15.6 |
| Vaccinology & Immunization | Studies that test potential vaccines for MERS-CoV infection, in addition to studies investigating the immunity to the disease after vaccination or being infected | 139 | 11 |
| Seroprevalence | Studies conducted to investigate the number of populations who tested positive for MERS-CoV based on serology specimens | 137 | 10.8 |
| Transmissibility | Studies investigating the passing of the MERS-CoV disease from one person to another person or from one animal to another animal | 105 | 8.3 |
| Outbreak investigation | Epidemiological studies that identify the source of MERS-CoV outbreak to establish control measures | 90 | 7.1 |
| Laboratory diagnostics | Studies that describe any diagnostic tests done to investigate the presence of MERS-CoV disease | 91 | 7.2 |
| Anthropology and social behavior | Studies conducted to explore population behavior towards MERS-CoV and how this might impact their health | 72 | 5.7 |
| Spillover | Studies conducted to describe the spreading of disease from animals to humans | 63 | 4.9 |
| Outbreak preparedness and response | Studies investigating the preparedness and response for MERS-CoV, including disease prioritization, and risk assessment | 60 | 4.7 |
| Infection prevention and Control (IPC) | Studies describing the approaches designed to prevent harm by infection to patients and health workers at the healthcare facilities level | 50 | 4 |
| Impact on health system\BOD & economic impact | Studies investigating the impact of MERS-CoV on health systems, and its burden, including morbidity, mortality, in addition to the economic burden | 46 | 3.6 |
| Mathematical models for MERS | Studies that help to quantify the dynamics of the disease by writing down a set of equations that mimics the reality | 40 | 3.2 |
| Comorbidities | Studies investigating the existence of any other medical condition than MERS-CoV | 40 | 3.2 |
| MERS & mental health | Studies that explore the impact of MERS-CoV on human mental health | 26 | 2.1 |
| Surveillance systems | Studies describing the surveillance systems set in place with the aim of continuous, and systematic collections, and evaluation of MERS-CoV data, and the dissemination of results | 22 | 1.7 |
| Case management | Studies examine how case managers provide health services to people infected with MERS-CoV, starting from screening to treating to evaluating. | 22 | 1.7 |
| MERS-CoV & SARS-CoV-2 cross-reactive immunity | Studies describing the relationship between previous MERS-CoV infection and the risk of SARS-CoV-2 infection as well as the relationship between previous MERS-CoV and COVID-19-related hospitalization and mortality | 20 | 1.6 |
| Silent/asymptomatic | Studies that have been done on people who tested positive for the MERS-CoV infection but never experienced symptoms | 15 | 1.2 |
| MERS & COVID-19 | Studies exploring the impact of the COVID-19 pandemic on MERS activities, and vice versa | 12 | 0.9 |
| One Health approach | Studies conducted using the one health approach with the aim of optimizing the health of people, animals, and the environment | 9 | 0.7 |
| Traditional medicine | Studies exploring the traditional health practices used by the community to maintain their well-being after being infected by MERS-CoV | 6 | 0.47 |
| MERS & pregnancy | Studies exploring the impact of MERS-CoV on pregnant women's health | 6 | 0.47 |
| Trade and animal food production | Studies that have been done to investigate the production of animal food derived from MERS-CoV animal reservoirs (e.g., meat and milk). | 5 | 0.4 |
| Reinfection | Studies investigating the reinfection possibility with MERS-CoV | 5 | 0.4 |

Table 5 maps the frequencies of MERS-CoV-related study themes over time, illustrating the dynamics of MERS-CoV-related research since its emergence in 2012. For instance, it shows an increase in studies aiming to understand the spillover mechanisms of the virus from animals to humans in 2014, in addition to an increase in studies conducted to produce therapeutics for MERS-CoV after 2019. Very few studies were conducted to examine the implementation of the One Health approach, most of which were conducted in 2019 (66.7%, n=6). Moreover, studies conducted to understand the MERS-CoV impact on health systems and its economic impact increased in 2019 (23.9%, n=11). 20 studies were published on MERS-CoV and SARS-CoV-2 cross-reactive immunity since 2020 (see Supplemental file 3).

**Table 5:** Mapping the frequencies of MERS-CoV-related studies themes included in the scoping review over the years (1 January 2012, 24 January 2023)

|  |  |  |  |  |  |  |  |  |  |  |  |  |
| --- | --- | --- | --- | --- | --- | --- | --- | --- | --- | --- | --- | --- |
| **Themes** | **2012** | **2013** | **2014** | **2015** | **2016** | **2017** | **2018** | **2019** | **2020** | **2021** | **2022** | **2023** |
| Molecular genetics | 0 | 13 | 42 | 28 | 43 | 37 | 45 | 45 | 43 | 67 | 51 | 3 |
| Therapeutics (drug-related or others) | 0 | 10 | 13 | 15 | 22 | 12 | 27 | 24 | 31 | 31 | 35 | 2 |
| Pathogenesis | 0 | 9 | 16 | 19 | 31 | 19 | 26 | 21 | 18 | 26 | 12 | 0 |
| Vaccinology & Immunization | 0 | 7 | 10 | 11 | 11 | 14 | 12 | 21 | 15 | 16 | 21 | 1 |
| Seroprevalence | 0 | 3 | 18 | 19 | 11 | 16 | 16 | 19 | 13 | 13 | 8 | 1 |
| Transmissibility | 0 | 4 | 11 | 9 | 19 | 13 | 14 | 13 | 9 | 5 | 8 | 0 |
| Outbreak investigation | 0 | 4 | 15 | 20 | 21 | 12 | 7 | 8 | 1 | 1 | 1 | 0 |
| Laboratory diagnostics | 0 | 4 | 5 | 14 | 12 | 10 | 14 | 11 | 9 | 10 | 2 | 0 |
| Anthropology and Social behavior | 0 | 0 | 0 | 4 | 11 | 18 | 13 | 10 | 11 | 4 | 1 | 0 |
| Spillover | 0 | 1 | 13 | 3 | 5 | 6 | 10 | 8 | 5 | 6 | 6 | 0 |
| Outbreak preparedness and response | 0 | 3 | 5 | 8 | 9 | 9 | 9 | 4 | 9 | 4 | 0 | 0 |
| Infection prevention and Control (IPC) | 0 | 0 | 2 | 5 | 10 | 8 | 6 | 5 | 7 | 4 | 2 | 1 |
| Impact on health system\BOD & | 0 | 1 | 4 | 4 | 6 | 4 | 5 | 11 | 6 | 5 | 0 | 0 |
| economic impact |  |  |  |  |  |  |  |  |  |  |  |  |
| Mathematical models for MERS | 0 | 0 | 4 | 5 | 8 | 4 | 4 | 4 | 5 | 2 | 4 | 0 |
| MERS & mental health | 0 | 0 | 0 | 0 | 5 | 4 | 5 | 4 | 5 | 2 | 1 | 0 |
| Surveillance systems | 0 | 0 | 5 | 3 | 6 | 1 | 3 | 0 | 4 | 0 | 0 | 0 |
| Case management | 0 | 1 | 2 | 2 | 7 | 1 | 2 | 1 | 3 | 3 | 0 | 0 |
| MERS-CoV & SARS-CoV-2 cross-reactive immunity | 0 | 0 | 0 | 0 | 0 | 0 | 0 | 0 | 4 | 9 | 7 | 0 |
| Silent/asymptomatic | 0 | 1 | 1 | 1 | 0 | 3 | 2 | 0 | 4 | 1 | 2 | 0 |
| Comorbidities | 0 | 0 | 1 | 3 | 3 | 6 | 5 | 7 | 6 | 8 | 1 | 0 |
| MERS & COVID-19 | 0 | 0 | 0 | 0 | 0 | 0 | 0 | 0 | 5 | 7 | 0 | 0 |
| One Health approach | 0 | 0 | 0 | 0 | 0 | 1 | 2 | 6 | 0 | 0 | 0 | 0 |
| Traditional medicine | 0 | 0 | 0 | 0 | 0 | 0 | 1 | 0 | 3 | 2 | 0 | 0 |
| MERS & pregnancy | 0 | 0 | 1 | 0 | 4 | 1 | 0 | 0 | 0 | 0 | 0 | 0 |
| Trade and animal food production | 0 | 0 | 2 | 0 | 0 | 1 | 0 | 0 | 1 | 0 | 1 | 0 |
| Reinfection | 0 | 0 | 0 | 1 | 1 | 2 | 0 | 0 | 1 | 0 | 0 | 0 |

Table 6 maps the frequencies of MERS-CoV-related study themes included in the scoping review by first-author institution country. The table depicts that outside the Eastern Mediterranean region (EMR), the Republic of Korea was the leading country for publications on most of the themes; for instance, transmissibility, outbreak investigation, laboratory diagnostics, and impact on the health system. However, publications from the USA exceeded the Republic of Korea when it came to therapeutics, vaccinology, pathogenesis, and MERS and SARS-CoV-2 cross-reactive immunity. Although China did not surpass the USA and Republic of Korea for most of the themes, it produced several studies on spillover mechanisms. For the EMR countries, KSA dominated the MERS-CoV-related research for all themes, except for the One Health approach, on which the United Arab Emirates also produced one study as the KSA (see Supplemental file 3).

**Table 6:**
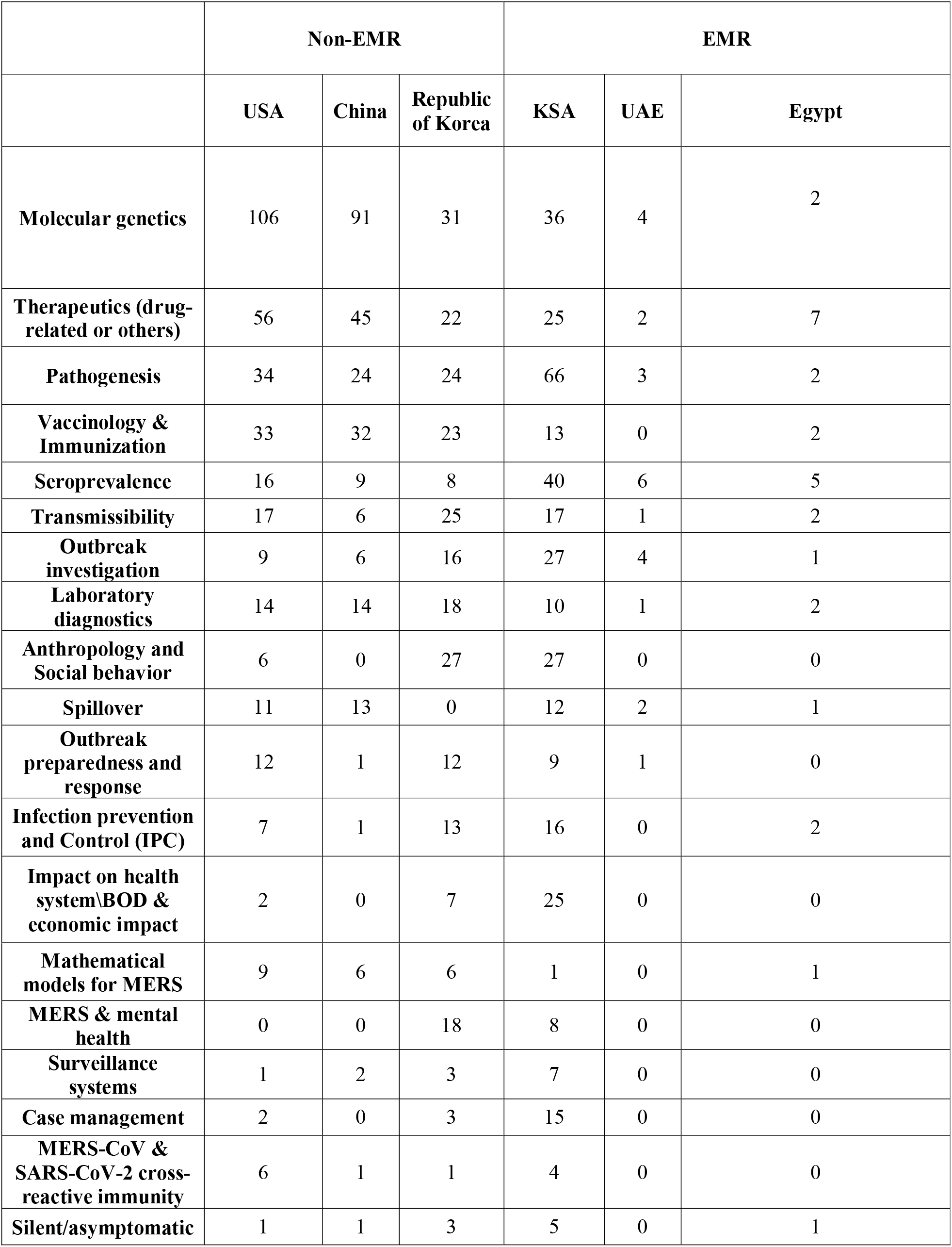
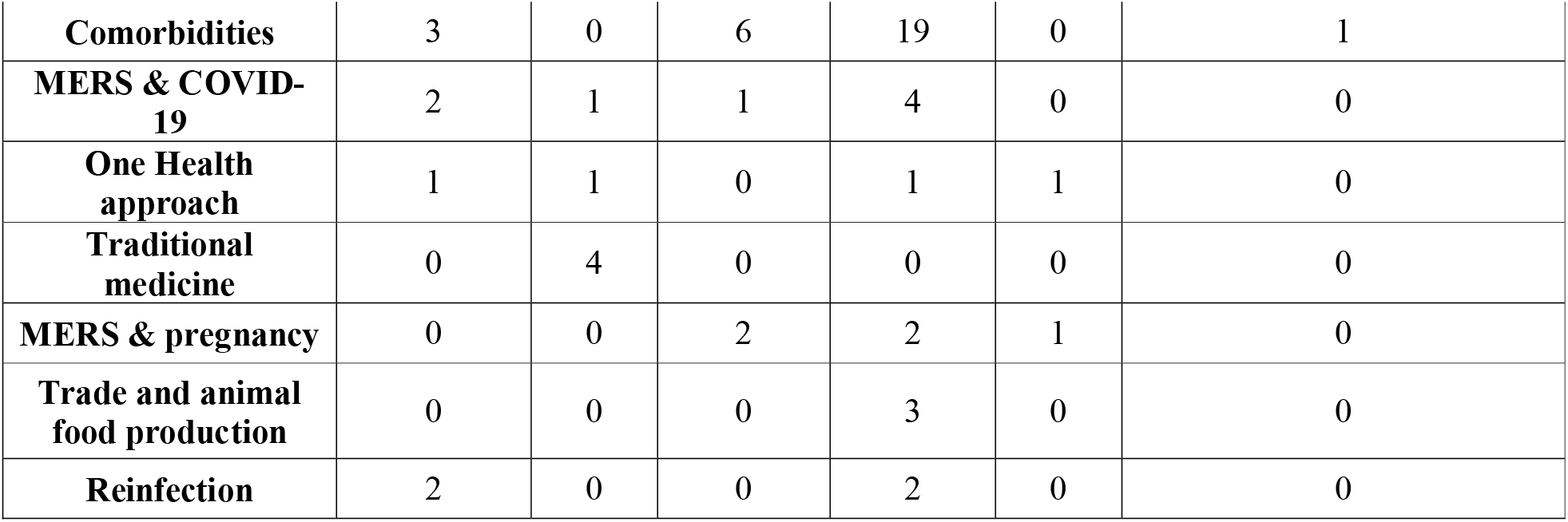
Mapping the frequencies of MERS-CoV-related studies themes included in the scoping review across countries in the EMR and Non-EMR (1 January 2012, 24 January 2023)

|  | Non-EMR |  |  | EMR |  |  |
| --- | --- | --- | --- | --- | --- | --- |
|  | USA | China | Republic of Korea | KSA | UAE | Egypt |
| **Molecular genetics** | 106 | 91 | 31 | 36 | 4 | 2 |
| **Therapeutics (drug-related or others)** | 56 | 45 | 22 | 25 | 2 | 7 |
| **Pathogenesis** | 34 | 24 | 24 | 66 | 3 | 2 |
| **Vaccinology & Immunization** | 33 | 32 | 23 | 13 | 0 | 2 |
| **Seroprevalence** | 16 | 9 | 8 | 40 | 6 | 5 |
| **Transmissibility** | 17 | 6 | 25 | 17 | 1 | 2 |
| **Outbreak investigation** | 9 | 6 | 16 | 27 | 4 | 1 |
| **Laboratory diagnostics** | 14 | 14 | 18 | 10 | 1 | 2 |
| **Anthropology and Social behavior** | 6 | 0 | 27 | 27 | 0 | 0 |
| **Spillover** | 11 | 13 | 0 | 12 | 2 | 1 |
| **Outbreak preparedness and response** | 12 | 1 | 12 | 9 | 1 | 0 |
| **Infection prevention and Control (IPC)** | 7 | 1 | 13 | 16 | 0 | 2 |
| **Impact on health system\BOD & economic impact** | 2 | 0 | 7 | 25 | 0 | 0 |
| **Mathematical models for MERS** | 9 | 6 | 6 | 1 | 0 | 1 |
| **MERS & mental health** | 0 | 0 | 18 | 8 | 0 | 0 |
| **Surveillance systems** | 1 | 2 | 3 | 7 | 0 | 0 |
| **Case management** | 2 | 0 | 3 | 15 | 0 | 0 |
| **MERS-CoV & SARS-CoV-2 cross-reactive immunity** | 6 | 1 | 1 | 4 | 0 | 0 |
| **Silent/asymptomatic** | 1 | 1 | 3 | 5 | 0 | 1 |
| **Comorbidities** | 3 | 0 | 6 | 19 | 0 | 1 |
| **MERS & COVID-19** | 2 | 1 | 1 | 4 | 0 | 0 |
| **One Health approach** | 1 | 1 | 0 | 1 | 1 | 0 |
| **Traditional medicine** | 0 | 4 | 0 | 0 | 0 | 0 |
| **MERS & pregnancy** | 0 | 0 | 2 | 2 | 1 | 0 |
| **Trade and animal food production** | 0 | 0 | 0 | 3 | 0 | 0 |
| **Reinfection** | 2 | 0 | 0 | 2 | 0 | 0 |

Table 7 shows the distribution of MERS-related study themes across the four scope levels: human, animal, animal-human interface, and environment. The table shows that scientists have produced hundreds of studies to evaluate countermeasures to decrease morbidity and mortality caused by MERS-CoV. Moreover, 81 papers were about developing multiplex diagnostic tools and platforms to simultaneously investigate multiple respiratory pathogens such as MERS-CoV and SARS-CoV-2 in humans [24, 25, 26]. Another well-researched in humans are pathogenesis studies conducted to better understand the duration of infectiousness in clinical settings, with more than 180 papers published. Also, IPC studies, aimed at decreasing infection risk factors among healthcare workers, gained relatively high interest, with 49 papers published on this topic. Seroprevalence studies accounted for 59 papers, most of which were conducted in KSA or to investigate MERS-CoV among pilgrims returning from KSA. On the other hand, the table shows that the role of silent/asymptomatic human cases in transmission received little attention to date (see Supplemental file 3).

**Table 7:** Mapping the frequencies of MERS-CoV-related studies themes included in the scoping review across the four levels (1 January 2012, 24 January 2023)

|  | **Human** | **Animal** | **Animal-human interface** | **Environment** |
| --- | --- | --- | --- | --- |
| **Molecular genetics** | 310 | 49 | 59 | 0 |
| **Therapeutics (drug-related or others)** | 215 | 3 | 4 | 0 |
| **Pathogenesis** | 186 | 7 | 5 | 0 |
| **Vaccinology & Immunization** | 128 | 13 | 2 | 0 |
| **Seroprevalence** | 59 | 53 | 25 | 0 |
| **Transmissibility** | 75 | 18 | 10 | 4 |
| **Outbreak investigation** | 83 | 3 | 5 | 1 |
| **Laboratory diagnostics** | 81 | 11 | 3 | 0 |
| **Anthropology and social behavior** | 71 | 0 | 1 | 0 |
| **Spillover** | 0 | 2 | 63 | 1 |
| **Outbreak preparedness and response** | 54 | 2 | 2 | 3 |
| **Infection prevention and Control (IPC)** | 49 | 0 | 0 | 1 |
| **Impact on health system\BOD & economic impact** | 45 | 0 | 1 | 0 |
| **Mathematical models for MERS** | 34 | 0 | 5 | 2 |
| **Comorbidities** | 39 | 0 | 2 | 0 |
| **MERS & mental health** | 26 | 0 | 0 | 0 |
| **Surveillance systems** | 22 | 0 | 0 | 0 |
| **Case management** | 22 | 0 | 0 | 0 |
| **MERS-CoV & SARS-CoV-2 cross-** | 20 | 0 | 0 | 0 |
| **reactive immunity** |  |  |  |  |
| **Silent/asymptomatic** | 10 | 3 | 3 | 0 |
| **MERS & COVID-19** | 10 | 0 | 0 | 0 |
| **One Health approach** | 0 | 0 | 9 | 2 |
| **Traditional medicine** | 3 | 0 | 3 | 0 |
| **MERS & pregnancy** | 6 | 0 | 0 | 0 |
| **Trade and animal food production** | 2 | 2 | 5 | 1 |
| **Reinfection** | 1 | 3 | 1 | 0 |

As indicated in table 7, there were extensive published studies investigating evidence of MERS-CoV circulation in animal populations through seroprevalence studies. 25 seroprevalence studies were conducted in the African region, with 10 studies in Kenya, 3 in Ethiopia, 2 each in Nigeria and Egypt, 1 each in Eastern Africa (Sudan and Somalia), Mali, Morocco, Ghana, Tunisia, Morocco, Ethiopia, and Burkina Faso, and 2 studies investigated the seroprevalence in camel populations in the whole African Region. Moreover, there were 13 publications on accelerating the development of countermeasures such as vaccine candidates. Transmissibility was another area of interest in studies targeting animal populations, with 18 studies conducted to better understand the risk of animal-to-animal transmission. On the other hand, there were only three papers published on evidence of re-infection with MERS-CoV among animals.

At the animal-human interface, there were about 60 studies evaluating the extent of spillover. On the other hand, there were only 9 papers on the One Health approach, in addition to only one anthropological study that describe and quantify the exposure of humans to camels.

## Discussion

To our knowledge, this is the first comprehensive scoping review mapping evidence generated on MERS-CoV in animal and human populations since its emergence in 2012 which highlights MERS-CoV-related tehnical areas receiving less attention and can inform future research. Several formal scoping and systematic reviews have previously been conducted, but they focused on specific topics either related to MERS-CoV in animal [13] or in human hosts [14].

Overall, there has been a substantial research effort on MERS-CoV with 1,264 studies analyzed in this review. Nevertheless, we found gaps in several technical areas. While acknowledging the time lag between research production and publication, the number of primary research produced on MERS-CoV tripled in 2014 (8.4%, n=106), as compared to 2013 (3.1%, n=39), likely triggered by a large healthcare-related outbreaks occurring in 2014 [6]. It is noteworthy that the number of research papers published on MERS-CoV did not decline during the COVID-19 pandemic, in part due to scientific interest in exploring similarities and differences between these two viruses from the same Coronavirus family, especially in the areas of molecular genetics, therapeutics, and vaccinology (Table 5) [25, 26]. Additionally, researchers started to explore if past MERS-CoV infection generates some level of cross-protective immunity against SARS-CoV-2 infection [27, 28, 29] (see Supplemental file 3).

Our review revealed that the largest percentage (57.8%, n=730) of studies were conducted in laboratory settings, mainly focusing on themes including molecular genetics, therapeutics, and vaccinology (Table 4). The smallest percentage of studies (0.5%, n=6) targeted camel events, such as races or beauty contests (Table 3).

Our review was organized into four scope levels, namely human, animal, animal-human interface and environment. The results show that at the human level, scientists made considerable efforts to investigate potential anti-MERS-CoV therapeutics [30, 31] and vaccine candidates [32, 33] to decrease morbidity and mortality. Nevertheless, no therapeutics or vaccines for MERS-CoV have been licensed to date, demonstrating that more effort should be invested to develop licensed medical countermeasures. Other research conducted at the human level included clinical studies aiming to understand the dynamics of MERS-CoV infection and its pathogenesis [34, 35], in addition to studies focusing on the development of multiplex diagnostic tools that test for multiple pathogens in parallel, including SARS-CoV-2 and MERS-CoV [23, 24, 25]. However, there is a need to now systematically integrate such multiplex tools into existing surveillance testing algorithms. 41 papers have investigated risk factors for MERS-CoV transmission among healthcare workers in hospital settings [36, 37]; however, the role of asymptomatic cases in transmission remains a research gap (See Supplemental file 3).

At the animal level, there is still a gap in studying antibody protection and potential reinfection of dromedary camels, which was one of the priority research areas highlighted by the FAO-WHO-WOAH technical meeting in 2017 [18]. On the other hand, other priorities at the animal level were addressed, such as studies investigating animal-to-animal transmission. Moreover, this review showed that more geographically representative seroprevalence studies should be conducted, particularly in regions with large camel populations. While our review revealed many publications on the evaluation of vaccine candidates for dromedary camels, advancement is still needed as no licensed animal vaccine is available to date [39, 40].

At the animal-human interface level, spillover from animals to humans has been studied in 63 studies, the most researched area. Nevertheless, more research effort is needed on applying a One Health approach in MERS prevention and control and enhance collaboration between animal, human, and environmental sectors, in addition to implementing more anthropological studies that describe the interactions of humans with camels in different geographic and cultural contexts and investigate related risks. While no specific research priorities were formulated for the environmental level by participants of the FAO-WHO-WOAH technical meeting in 2017 [18], there is a need to expand research in that area, with most of the studies to date focusing on the role of climate factors and international travel in MERS-CoV transmission [43].

This review identified 59 countries that have published MERS-related research as first-author institution countries. Although the USA is not a directly affected country, it was the leading country in terms of number of publications produced likely due to the amount of funding for pandemic preparedness available to institutions in this location. KSA, China, and Republic of Korea followed the USA in the number of first-authored MERS-related publications. Most of the 12 EMR countries that have reported MERS-CoV cases since 2012 [6] have published studies on the virus in several research areas, except for Tunisia and Bahrain (see Figure 2 and Supplemental file 3). Table 6 shows that KSA has produced most MERS-related publications among the EMR countries (18.6%, n=235). This is expected with 84% of laboratory-confirmed MERS-CoV cases in EMR being reported from KSA (2,196 cases and 855 related deaths as of May 2023) [6]. Notably, 75.2% of the studies conducted on MERS-CoV received funding, while only 11.7% did not receive any financial assistance; more advocacy on resource mobilization is needed especially targeting research that addresses key outstanding gaps.

The results of this review can be used to identify MERS-CoV-related technical areas that received less attention by published research to date, while minimizing overlap and duplication of work and efforts. By mapping all first-author institutions working on MERS-CoV research and related funding agencies, we aim to provide researchers worldwide with information and resources for planning future work on MERS-CoV, e.g. to reach out to potential collaborators or identify funders interested in supporting their research area (see Supplemental file 3).

Some limitations of this review should be noted. We only searched three databases, the minimum requirement for a scoping review. Another limitation is that we targeted only peer-reviewed publications and thus did not include records from grey literature such as websites of international organizations or governmental agencies. In addition, the decision to only include English abstracts may have introduced publication bias.

## Conclusions

This scoping review has several research and policy implications. Here we organize our conclusions and recommendations under five main categories, adapted from the topics discussed in the 2017 and 2021 FAO-WHO-WOAH technical meetings [18, 19] (see Table 8):

**Table 8:**
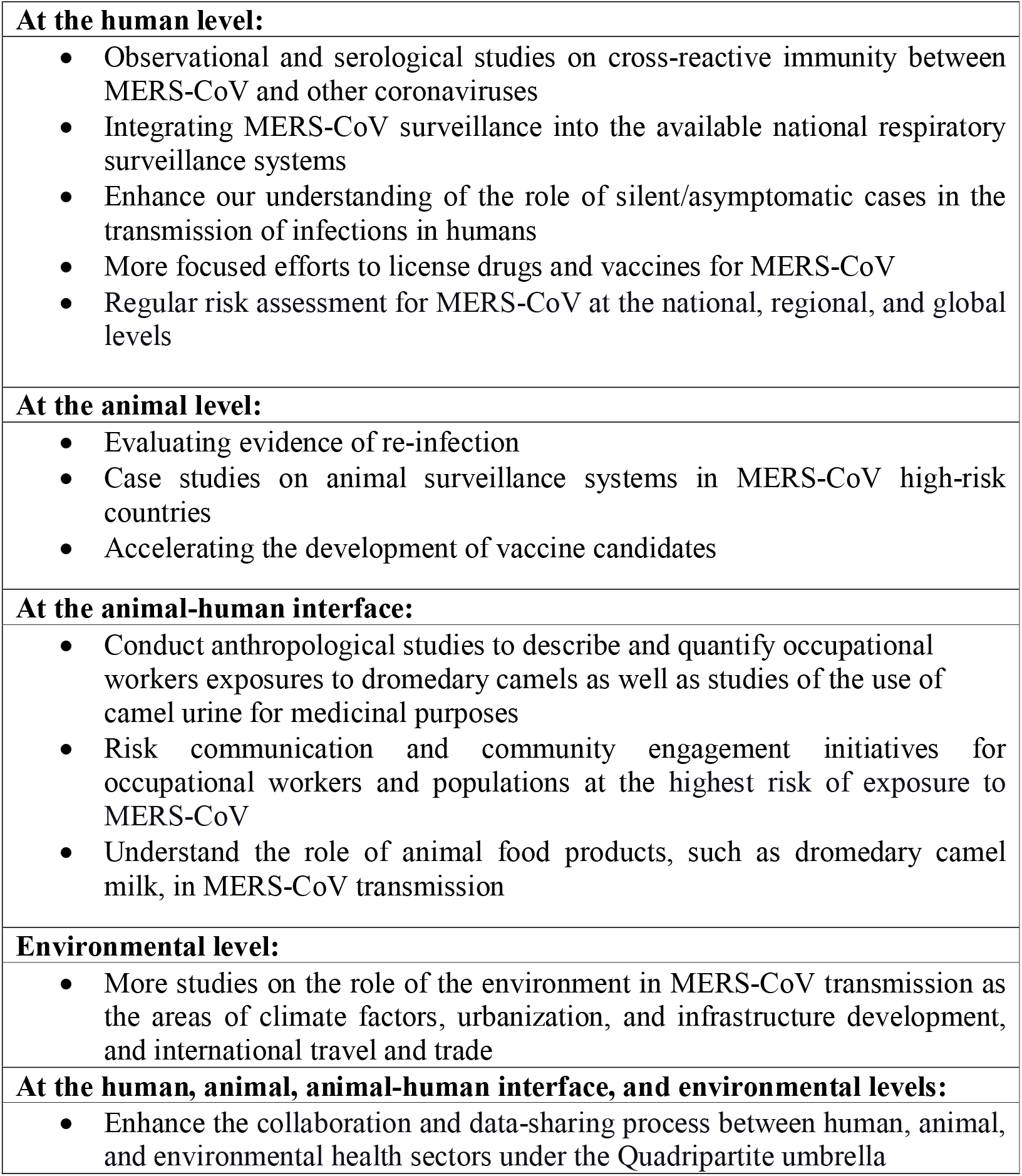
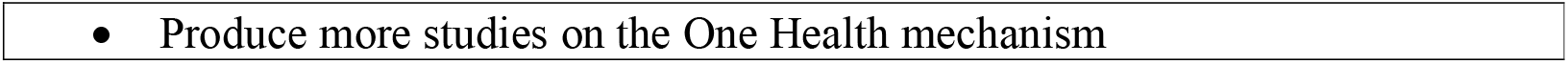
Recommendations and priority actions for MERS-CoV research.

### 1. Strengthening surveillance in animals and humans

Collaborative surveillance should be implemented by integrating MERS-CoV testing into the available national respiratory surveillance systems where appropriate and increasing laboratory detection capacity, also through strengthening the development and use of multiplex kits for SARS-CoV-2, influenza, MERS-CoV, and other respiratory viruses. It is essential to implement camel surveillance systems in EMR countries and African countries with large camel populations to allow detection and monitoring of MERS-CoV circulation and explore any patterns, such as seasonality. In addition, more longitudinal studies should be conducted in dromedary camels to better understand the natural history, shedding profile and immunity. Also, more analyses of camel value chain and production systems need to be undertaken in order to better target surveillance and risk mitigation efforts.

### 2. Infection prevention and control to reduce human infections and hospital transmission

Healthcare workers at health facilities are at higher risk of being exposed to the virus; in fact, the number of MERS-CoV hospital-acquired cases is by far exceeding that of community-acquired cases [6]. Reinforcing IPC awareness and implementation remains critical to preventing the possible spread of MERS-CoV in healthcare facilities. Moreover, to strengthen clinical care, the potential role of asymptomatic or mild cases in disease transmission should be further investigated.

### 3. Accelerating therapeutics and vaccine research and development

Despite the notable global scientific effort to develop and test vaccine and drug candidates for animals and humans, there are still no approved vaccines or therapeutics against MERS-CoV available to date; therefore, it is essential to accelerate advances for human vaccines and exert additional efforts for animal vaccine research to limit the spread of infection in animal populations and reduce the spillover risk from animals to humans. Moreover, while developing countermeasures, it is vital to initially ensure equitable access through benefit-sharing agreements.

### 4. Enhancing community engagement, community protection and risk mitigation

Our review revealed a continuing lack of anthropological and social behavioral studies that describe and quantify the occupational exposures of workers to dromedary camels; therefore, it is essential to conduct more research in this area so that effective risk-mitigating interventions can be designed. Camel workers in farms, animal markets, slaughterhouses, and abattoirs are at the highest risk of zoonotic exposure to MERS-CoV; hence, proactive risk communication and community engagement will enhance protection of the whole community. For instance, raising awareness for personal protective measures among camel workers, such as personal hygiene practices and protective clothing, boots, and facial protection, is vital. Although no confirmed MERS case has ever been in connection with Hajj, it is essential to enhance risk communication programs during these religious events of great magnitude, not only in KSA, where Hajj and Umrah take place, but also for Muslim pilgrims in their countries of origin.

### 5. Reinforcing multi-sectoral coordination

Finally, the most critical pillar for health emergency preparedness, prevention and response, and resilience in the context of zoonotic pathogens is the collaboration between human, animal, and environmental health sectors using a One Health approach. Better coordination between the different sectors by improving the data-sharing process and producing multisectoral national risk assessments and contingency plans is essential for preparedness, operational readiness, and risk reduction. Moreover, more effort should be exerted to raise awareness with the environmental sector regarding their involvement in MERS-CoV prevention and control efforts.

Human MERS-CoV cases continue to be reported to WHO. Given the pandemic potential of the virus and the high case fatality rate, the world has to remain alert and continue to further strengthen preparedness and prevention efforts. Even though research efforts have been continuing for MERS-CoV, our review found that a number of MERS-CoV-related technical areas already documented by FAO, WHO and WOAH in 2017 and 2021 have not been sufficiently addressed to date. A targeted and coordinated effort is needed to advance the MERS-CoV research agenda and close the remaining gaps.

## Declarations

## Supporting information

PRISMA Checklist

Technical appendix

## Data Availability

All data produced in the present study are available upon reasonable request to the authors

## List of abbreviations used

MERS-CoV: Middle East respiratory syndrome coronavirus
WHO: World Health Organization
PRISMA-ScR: Systematic Reviews and Meta-Analyses Extension for Scoping Reviews
BoD: Burden of Disease
FAO: Food and Agriculture Organization
WOAH: the World Organisation for Animal Health
R&D: Research and Development
CFR: Case-Fatality Ratio
EMR: Eastern Mediterranean Region

## a) Acknowledgments

Our sincere thanks to all authors, and institutions who have published studies on MERS-CoV, and contributed to bridging the research gap for this pathogen. Furthermore, we would like to extend our thanks to the Public Health Authority of Saudi Arabia, which serves as the WHO collaborating centre for MERS, for their contribution to the final draft of the manuscript. Finally, we would like to thank Ms. Gehan Al Garraya for her help in validating the search strategy.

## c) Availability of data and materials

All data relevant to the study are included in the article or uploaded as supplementary information.

## d) Authors’ contributions

WK, MDVK, HAEN, SVD and MH conceptualized the study. MH and HAEN contributed to the study design, writing of the first draft of the manuscript, and review. MH and HY collected data. MH conducted the analysis. All co-authors reviewed and validated it.

## e) Ethics approval and consent to participate

Not applicable

## f) Consent for publication

Not applicable

## List of Additional data files

- **Supplemental file 1:** Preferred Reporting Items for Systematic Reviews and Meta-Analyses Extension for Scoping Reviews (PRISMA-ScR) Checklist
- **Supplemental file 2:** Detailed search strategy for PubMed, EMBASE, CINHAL
- **Supplemental file 3:** List of supplementary materials including maps arised from the review
- **Supplemental file 4:** Technical appendix

