## Supplementary material for "Mapping the Middle East Respiratory Syndrome (MERS) related Research – A Scoping Review (2012-2023)": Figures

Records identified through database* searching:

n = 6,690

Databases*:

**CINHAL:** 856, **PubMed:** 1751, **Embase:** 4,083

**Identification**

Records after duplicate removal

(n = 3,825)

● 2,188 records excluded as not related to MERS-CoV

● 91 records excluded as only abstracts were available

**Screening**

Title and abstract screening

(n = 3,825)

Full-text records excluded (n = 282) for the following reasons:

- Duplication (n=6)
- Not related to MERS-CoV (n=35)
- Not study type of interest (n=198)
- Records in languages other than English (n=43):

Chinese (n=16), French (n=3), Japanese (n=1)

Korean (n=11), Polish (n=1), Russian (n=1)

Spanish (n=2), Deutsch (n=2), Greek (n=1)

Turkish (n=2), Nordic (n=2), Persian (n=1)

**Eligibility**

Full-text records assessed for eligibility

(n = 1,546)

Studies included

(n = 1,264)

**Included**

Figure 1: **PRISMA:** *Flowchart of the selection process for the scoping review of the Middle East Respiratory Syndrome Coronavirus studies and results, (1 January 2012 to 24 January 2023)*

|  | 100 – 300 records |
| --- | --- |
|  | 50 – 99 records |
|  | 10 – 49 records |
|  | 1 – 9 records |
|  | 0 records |

**Figure 2:** Map of first-author countries that conducted studies on MERS-CoV, (1 January 2012 to 24 January 2023)

Figure 3: Number of MERS-related articles/studies across study period by publication date, (1 January 2012 to 24 January 2023)

Cumulative number of MERS related studies (n = 1264)

| Year | Cumulative number of MERS related studies (n = 1265) |
| --- | --- |
| 2012 | 0 |
| 2013 | 39 |
| 2014 | 106 |
| 2015 | 114 |
| 2016 | 163 |
| 2017 | 134 |
| 2018 | 147 |
| 2019 | 143 |
| 2020 | 150 |
| 2021 | 147 |
| 2022 | 116 |
| 2023 | 5 |

Figure 4: Number of MERS-related articles/studies included in the scoping review by scope level, (1 January 2012 to 24 January 2023)

Number of studies (n = 1264)

| Scope Level | Cumulative number of studies (n = 1264) |
| --- | --- |
| Human | 1035 |
| Animal | 130 |
| Animal-human interface | 109 |
| Environment | 9 |

Figure 5: Number of MERS-related articles/studies included in the scoping review by themes, (1 January 2012 to 24 January 2023)

Number of studies (n=1264)

|  | Number of studies (n = 1264) |
| --- | --- |
| Molecular genetics | 417 |
| Therapeutics (drug-related or others) | 222 |
| Clinical features & complications | 197 |
| Vaccinology & Immunization | 139 |
| Seroprevalence | 137 |
| Transmissibility | 105 |
| Outbreak investigation | 90 |
| Laboratory diagnostics | 91 |
| Anthropology and Social behavior | 72 |
| Spillover | 63 |
| Outbreak preparedness and response | 60 |
| Infection prevention and Control (IPC) | 50 |
| Impact on health system\BOD & economic impact | 46 |
| Mathematical models for MERS | 40 |
| Comorbidities | 40 |
| Surveillance systems | 22 |
| MERS-CoV & SARS-CoV-2 cross-reactive immunity | 20 |
| Silent/asymptomatic | 15 |
| One Health approach | 9 |
| Others | 82 |

**Figure 5:** *Number of MERS-CoV-related studies included in the scoping review by themes, (1 January 2012 to 24 January 2023)*
