## Supplemental File 1 for "Mapping the Middle East Respiratory Syndrome (MERS) related Research – A Scoping Review (2012-2023)"

**Supplementary File 2: Search strategy**

**Databases Searched:**

Database

PubMed

Embase

CINHAL

**Search Strategy for each database:**

1. **PubMed**

| **Concept** | **MeSH Heading** | **Keywords** |
| --- | --- | --- |
| Middle east respiratory syndrome | "Middle East Respiratory Syndrome Coronavirus"[Majr:NoExp] | - MERS Virus - MERS Viruses - Virus, MERS - Middle East respiratory syndrome-related coronavirus - Middle East respiratory syndrome related coronavirus - Respiratory Syndrome, Middle East - Middle East Respiratory Syndrome Coronavirus Infection - MERS-CoV - MERS coronavirus - Merbecovirus - Merbecoviruses - MERS coronavirus infection - MERS infection - MERS virus infection - MERS-CoV infection - Middle East respiratory syndrome coronavirus infection - Middle East respiratory syndrome infection |
|  | "Middle East respiratory syndrome vaccine" [Majr:NoExp] | - ChAdOx1 MERS - ChAdOx1 MERS vaccine - Middle East respiratory syndrome vaccine ChAdOx1 MERS |
|  | "nucleocapsid protein, MERS-CoV" [Majr:NoExp] | - nucleocapsid protein, middle east respiratory syndrome coronavirus |
|  | "Nsp3 protein, Middle East respiratory syndrome coronavirus" [Majr:NoExp] | - Nsp3 protein, MERS-CoV |
|  |  | - MERS-CoV (EMC/2012) - MERS-CoV(HCoV-EMC/2012 strain) - MERS-CoV (strain EMC/2012) - MERS-CoV-EMC/2012 |
|  |  | - MERS (strain Hu/Jordan-N3/2012) |

**Keywords table:**

| **Concept** | **Keywords strategy** |
| --- | --- |
| Middle east respiratory syndrome | ("MERS"[title/abstract] OR "MERS-CoV"[title/abstract] OR "Merbecovirus*"[title/abstract] OR "middle east respiratory syndrome"[title/abstract] OR "ChAdOx1"[title/abstract]) |

| **#** | **Query** | **Results from 24 January 2023** |
| --- | --- | --- |
| **1** | "Middle East Respiratory Syndrome Coronavirus"[Majr:NoExp][title/abstract] | [1,426](https://login.research4life.org/?term=%22Middle+East+Respiratory+Syndrome+Coronavirus%22%5BMajr%3ANoExp%5D&ac=no&sort=relevance) |
| **2** | "Middle East respiratory syndrome vaccine" [Majr:NoExp][title/abstract] | 0 |
| **3** | "nucleocapsid protein, MERS-CoV" [Majr:NoExp][title/abstract] | 0 |
| **4** | "Nsp3 protein, Middle East respiratory syndrome coronavirus" [Majr:NoExp][title/abstract] | 0 |
| **5** | ("MERS"[title/abstract] OR "MERS-CoV"[title/abstract] OR "Merbecovirus*"[title/abstract] OR "middle east respiratory syndrome"[title/abstract] OR "ChAdOx1"[title/abstract]) | 9,956 |
| **6** | #1 or #2 or #3 or #4 or #5 | 10,004 |
| **7** | 2012 to 2023 | 1,751 |

2. **Embase:**

| **Concept** | **Emtree** | **Keywords** |
| --- | --- | --- |
| Middle east respiratory syndrome | 'Middle East respiratory syndrome coronavirus'/exp | - MERS Virus - MERS Viruses - Virus, MERS - Middle East respiratory syndrome-related coronavirus - Middle East respiratory syndrome related coronavirus - Respiratory Syndrome, Middle East - Middle East Respiratory Syndrome Coronavirus Infection - MERS-CoV - MERS coronavirus - Merbecovirus - Merbecoviruses - MERS coronavirus infection - MERS infection - MERS virus infection - MERS-CoV infection - Middle East respiratory syndrome coronavirus infection - Middle East respiratory syndrome infection |
|  | 'Middle East respiratory syndrome coronavirus (strain EMC/2012)'/de | - MERS-CoV (EMC/2012) - MERS-CoV(HCoV-EMC/2012 strain) - MERS-CoV (strain EMC/2012) - MERS-CoV-EMC/2012 |
|  | 'Middle East respiratory syndrome coronavirus (strain Hu/Jordan-N3/2012)'/de | - MERS (strain Hu/Jordan-N3/2012) |
|  | 'Middle East respiratory syndrome'/de | - MERS coronavirus infection - MERS infection - MERS virus infection - MERS-CoV infection - Middle East respiratory syndrome coronavirus infection - Middle East respiratory syndrome infection |
|  | 'middle east respiratory syndrome coronavirus vaccine'/de | - ChAdOx1 MERS - ChAdOx1 MERS vaccine - Middle East respiratory syndrome vaccine ChAdOx1 MERS |
|  | 'middle east respiratory syndrome vaccine'/de |  |
|  |  | - nucleocapsid protein, middle east respiratory syndrome coronavirus |
|  |  | - Nsp3 protein, MERS-CoV |

**Keywords table:**

| **Concept** | - **Keywords strategy** |
| --- | --- |
| Middle east respiratory syndrome | - ((MERS NEAR/3 (virus* OR coronavirus*)):ab,ti,kw - (MERS):ab,ti,kw - (MERS NEXT/1 CoV):ab,ti,kw - (Merbecovirus*):ab,ti,kw - (Middle East NEAR/3 respiratory):ab,ti,kw - (ChAdOx1):ab,ti,kw |

| **#** | **Query** | **Results from 24 January 2023** |
| --- | --- | --- |
| **1** | 'Middle East respiratory syndrome coronavirus'/exp | 4957 |
| **2** | 'Middle East respiratory syndrome coronavirus (strain EMC/2012)'/de | 6 |
| **3** | 'Middle East respiratory syndrome coronavirus (strain Hu/Jordan-N3/2012)'/de | 1 |
| **4** | 'Middle East respiratory syndrome'/de | 3001 |
| **5** | 'middle east respiratory syndrome coronavirus vaccine'/de | 50 |
| **6** | 'middle east respiratory syndrome vaccine'/de | 14 |
| **7** | (MERS NEAR/3 virus*):ab,ti,kw | 336 |
| **8** | (Merbecovirus*):ab,ti,kw | 22 |
| **9** | ('Middle East' NEAR/3 respiratory):ab,ti,kw | 3,830 |
| **10** | (ChAdOx1):ab,ti,kw | 1786 |
| **11** | (MERS):ab,ti,kw | 8,505 |
| **12** | (#1 or #2 or #3 or #4 or #5 or #6) and (#7 or #8 or #9 or #10 #11) | 4,083 |

3. **CINAHL**

| **Concept** | **CINAHL Subject Headings** | **Keywords** |
| --- | --- | --- |
| Middle east respiratory syndrome | MM "Middle East Respiratory Syndrome" | - MERS Virus - MERS Viruses - Virus, MERS - Middle East respiratory syndrome-related coronavirus - Middle East respiratory syndrome related coronavirus - Respiratory Syndrome, Middle East - Middle East Respiratory Syndrome Coronavirus Infection - MERS-CoV - MERS coronavirus - Merbecovirus - Merbecoviruses - MERS coronavirus infection - MERS infection - MERS virus infection - MERS-CoV infection - Middle East respiratory syndrome coronavirus infection - Middle East respiratory syndrome infection |
|  | MM "Middle East Respiratory Syndrome Coronavirus" |  |
|  |  | - ChAdOx1 MERS - ChAdOx1 MERS vaccine - Middle East respiratory syndrome vaccine ChAdOx1 MERS |
|  |  | - nucleocapsid protein, middle east respiratory syndrome coronavirus |
|  |  | - Nsp3 protein, MERS-CoV |
|  |  | - MERS-CoV (EMC/2012) - MERS-CoV(HCoV-EMC/2012 strain) - MERS-CoV (strain EMC/2012) - MERS-CoV-EMC/2012 |
|  |  | MERS (strain Hu/Jordan-N3/2012) |

**Keywords table:**

| **Concept** | **Keywords strategy** |
| --- | --- |
| Middle east respiratory syndrome | AU ( (MERS N3 virus*) OR MERS OR Merbecovirus* OR (Middle W0 East) N3 (respiratory NEXT/1 syndrome) OR ChAdOx1) ) OR TI ( (MERS N3 virus*) OR MERS OR Merbecovirus* OR (Middle W0 East) N3 (respiratory NEXT/1 syndrome) OR ChAdOx1) ) OR AB ( (MERS N3 virus*) OR MERS OR Merbecovirus* OR (Middle W0 East) N3 (respiratory NEXT/1 syndrome) OR ChAdOx1) ) |

| **#** | **Query** | **Results from 24 January 2023** |
| --- | --- | --- |
| **1** | MM "Middle East Respiratory Syndrome" | 0 |
| **2** | MM "Middle East Respiratory Syndrome Coronavirus" | 539 |
| **3** | AU ( (MERS N3 virus*) OR MERS OR Merbecovirus* OR (Middle W0 East) N3 (respiratory NEXT/1 syndrome) OR ChAdOx1) ) OR TI ( (MERS N3 virus*) OR MERS OR Merbecovirus* OR (Middle W0 East) N3 (respiratory NEXT/1 syndrome) OR ChAdOx1) ) OR AB ( (MERS N3 virus*) OR MERS OR Merbecovirus* OR (Middle W0 East) N3 (respiratory NEXT/1 syndrome) OR ChAdOx1) ) | 1,832 |
| **4** | (#1 or #2) and #3 | 910 |
| **5** | 2012 to 2023 | 856 |
