## Supplemental File 2 for "Mapping the Middle East Respiratory Syndrome (MERS) related Research – A Scoping Review (2012-2023)"

List of first-author institutions that have published on MERS-CoV globally (n=495):

| **Country** | **First author institution** | **Appendix S1: Technical Appendix Reference** |
| --- | --- | --- |
| **USA** | Baylor College of Medicine | [408] |
|  | California Department of Public Health | [675] |
|  | Centers for Disease Control and Prevention | [32], [149], [190], [213], [229], [276], [383], [434], [443], [456], [463], [565], [658], [851], [516], [683], [820], [887], [912], [989], [1085], [1160], [1190], [1214], [1220], [1263] |
|  | Collaborsations Pharmaceuticals, Inc. | [1208] |
|  | Colorado State University | [164], [397], [623], [1002], [1206] |
|  | Columbia University | [490], [814] |
|  | Cornell University | [37], [180], [539], [798] |
|  | Dana-Farber Cancer Institute | [1228] |
|  | Duke University School of Medicine | [862] |
|  | EcoHealth Alliance | [782] |
|  | Firebird Biomolecular Sciences | [331] |
|  | Florida International University | [158] |
|  | Foundation for Applied Molecular Evolution | [1093] |
|  | Fred Hutchinson Cancer Research Center | [751] |
|  | Georgia State University | [436], [702] |
|  | Global Center for Health Security | [182] |
|  | Harvard Medical School | [567] |
|  | Henry Jackson Foundation for the Advancement of Military Medicine | [553] |
|  | Jiann-Ping Hsu College of Public Health | [1229] |
|  | Johns Hopkins University | [187], [281], [636] |
|  | Kansas State University | [830] |
|  | Laboratory of Infectious Diseases | [957] |
|  | Lawrence Livermore National Laboratory | [803] |
|  | Leidos Biomedical Research Inc | [1034] |
|  | Loyola University Chicago | [44], [141], [370], [747], [962], [993], [1143], [1203] |
|  | Loyola University Chicago, Maywood | [1143] |
|  | Massachusetts Institute of Technology | [846], [870] |
|  | National Cancer Institute (NCI) | [272] |
|  | National Institutes of Health | [3], [33], [80], [99], [134], [186], [376], [379], [399], [410], [439], [452], [481], [527], [541], [596], [610], [622], [642], [648], [688], [708], [748], [765], [767], [834], [903], [954], [955], [956], [1001], [1053], [1126], [1172], [1137], [1177], [1224], [1235] |
|  | Nationwide Children's Hospital | [849] |
|  | Nebraska Biocontainment Unit | [102] |
|  | New York Blood Center | [49], [84], [563], [564], [649], [1044], [1187] |
|  | New York University | [166] |
|  | Northwestern University Feinberg School of Medicine | [252] |
|  | NYU Grossman School of Medicine | [1181] |
|  | Oklahoma State University | [106] |
|  | ORISE Fellowship Training Program | [614] |
|  | Pennsylvania State University | [28] |
|  | Purdue University | [109], [185], [314], [687], [1146], [1261] |
|  | Quantitative Biosciences Institute (QBI) | [248] |
|  | Regeneron Pharmaceuticals | [551], [929] |
|  | Saint Louis University | [1239] |
|  | Saint Louis Zoo Institute for Conservation Medicine | [1060] |
|  | Scripps Research Institute | [585] |
|  | Tennessee State University | [916] |
|  | Terumo BCT, Lakewood | [612] |
|  | Texas Tech University | [364] |
|  | Thomas Jefferson University | [882] |
|  | Tuskegee University | [24] |
|  | University of California | [958], [1036] |
|  | University of Central Florida | [455] |
|  | University of Florida | [431] |
|  | University of Georgia | [1109], [1112] |
|  | University of Illinois | [559], [635], [662] |
|  | University of Iowa | [285], [363], [349], [398], [578], [644], [783], [815], [819], [848], [959], [983], [1073], [1156] |
|  | University of Kansas | [729] |
|  | University of Maryland | [87], [88], [188], [240], [326], [513], [749], [911], [971], [1183], [1259] |
|  | University of Michigan | [884] |
|  | University of Minnesota | [23], [305], [701], [858], [991] |
|  | University of Missouri School of Medicine | [446] |
|  | University of New York | [1159] |
|  | University of North Carolina | [61], [81], [98], [175], [253], [512], [616], [847], [873], [902], [1011] |
|  | University of North Texas | [380], [969] |
|  | University of Pennsylvania | [83], [126], [728], [807] |
|  | University of Pittsburgh | [36], [40], [586], [758] |
|  | University of South Florida | [7] |
|  | University of Texas | [633], [808], [1100], [1107], [1164] |
|  | University of Texas | [171], [200], [441], [496], [583], 723], [724], [901], [906], [990], [1092], [1227] |
|  | University of Texas Rio Grande Valley | [122] |
|  | University of Utah | [16] |
|  | University of Washington | [311], [1118], [1121], [1238] |
|  | University School of Medicine Atlanta | [1037] |
|  | Vanderbilt University Medical Center | [1157] |
|  | Virginia Polytechnic Institute and State University | [1025] |
|  | Virology Division of the US Army Medical Research Institute of Infectious Diseases | [1077] |
|  | Walter Reed Army Institute of Research | [277], [1033] |
|  | Washington University School of Medicine | [6] |
|  | Wichita State University | [34], [1119] |
|  | Wistar Institute | [645] |
|  | Yale University | [856] |
|  | Zalgen Labs | [294] |
| **Saudi Arabia** | Alfarabi College of Dentistry | [667] |
|  | Almana General Hospital | [889] |
|  | Imam Abdulrahman Bin Faisal University | [580], [698], [669], [917] |
|  | Dr. Sulaiman Alhabib Medical Group | [1067] |
|  | Environmental Science and Arid Land Agriculture | [651] |
|  | Global Center for Mass Gatherings Medicine | [517] |
|  | Health Affairs in Makkah | [323] |
|  | Jazan University | [402] |
|  | Johns Hopkins Aramco Healthcare | [521], [600], [620], [629], [793], [975] |
|  | Joint Program of Postgraduate Studies of Family Medicine | [342] |
|  | Jouf University | [161], [968] |
|  | Khubash General Hospital | [320] |
|  | King Abdulaziz City for Science and Technology | [508], [510], [880], [1080], [1082], [1083] |
|  | King Abdulaziz Medical City | [533], [627], [640], [665], [706], [760], [789], [891], [892], [908] |
|  | King Abdulaziz University | [56], [107], [132], [179], [233], [265], [295], [330], [340], [346], [415], [442], [502], [587], [592], [605], [736], [754], [769], [866], [914], [966], [1063] |
|  | King Abdullah International Medical Research Center | [193], [291], [556], [529], [588], [1215], [1222] |
|  | King Abdullah University of Science and Technology | [986] |
|  | King Fahad Armed Forces Hospital | [290], [579], [730] |
|  | King Fahd General Hospital | [719], [1223] |
|  | King Fahad Medical City | [47], [96], [108], [308], [739], [839], [840] |
|  | King Fahad University Hospital | [470] |
|  | King Faisal University | [21], [70], [114], [118], [125], 129], [239], [247], [279], [341], [367], [435], [462], [486], [549], [678], [680], [696], [713], [733], [838], [945], [1008], [1039], [1052], [1076], [1078], [1079], [1134], [1178], [1180] |
|  | King Faisal Specialist Hospital & Research Center | [94], [105], [184], [519], [526], [594], [619], [693], [763], [832], [869], [904], [1018], [1019] |
|  | King Khalid University | [668] |
|  | King Khalid University Hospital | [142], [145], [871] |
|  | King Saud Bin Abdulaziz University for Health Sciences | [111], [140], [151], [218], [319], [322, [353], [381], [451], [472], [477], [478, [576], [656], [661], [704], [740], [810], [907], [1014], [1068], [1179], [1185, [1237] |
|  | King Saud University | [17], [162], [178], [197], [215], [217], [228], [327], [360], [401], [418], [433], [505], [566], [576], [603], [607], [732], [761], [773], [786], [796], [888], [925], [944], [1041], [1048], [1058] |
|  | King Saud Medical City | [422], [518], [828], [1242] |
|  | Medical Department Saudi Aramco Medical Services Organization Dhahran Saudi Arabia | [822] |
|  | Ministry of Environment, Water and Agriculture | [296], [1191] |
|  | Ministry of Health | [15], [226], [423], [476], [535], [625], [752], [777], [778], [790], [794], [795], [802], [850], [893], [1040, [1042], [1127], [1128] |
|  | Ministry of National Guard Health Affairs (MNGHA) | [1066] |
|  | Prince Sattam bin Abdulaziz University | [709] |
|  | Prince Sultan Military Medical City | [51], [196], [214], [943], [1013] |
|  | Prince Mohamed Bin Abdulaziz Hospital | [447], [937] |
|  | Princess Nourah Bint Abdulrahman University | [113], [287] |
|  | Qassim University | [318], [690] |
|  | Regional Laboratory | [234] |
|  | Riyadh Municipality Central Area Labs | [338] |
|  | Saudi Center for Disease Prevention and Control (Saudi CDC) | [210] |
|  | Sulaiman Al Rajhi Colleges | [225] |
|  | Taibah University | [738] |
|  | Taif University | [1189] |
|  | Umm Al-Qura University | [845], [976], [444] |
|  | University Campus | [537] |
|  | University of Al-Baha | [1184] |
|  | University of Dammam | [1262] |
|  | University of Tabuk | [1065] |
|  | University of Warwick | [46] |
| **Republic of Korea** | Ajou University | [949], [1110] |
|  | Armed forces medical research institute | [1260] |
|  | Asan Medical Center | [523] |
|  | Catholic University of Korea | [347] |
|  | Catholic University of Korea | [13], [150], [288], [522], [711], [750], [890] |
|  | Center for Infectious Diseases | [582] |
|  | Chonnam National University | [236], [396] |
|  | Chung-Ang University College of Medicine | [1130] |
|  | Chungbuk National University | [497], [554], [555], [824], [825], [883] |
|  | Chungnam National University | [43], [148], [153], [491], [524], [694], [1049], [1133] |
|  | Daegu-Gyeongbuk Medical Innovation Foundation | [1202] |
|  | Dankook University | [223], [386] |
|  | Division of Epidemic Intelligence Service | [1022] |
|  | Dongguk University | [1162] |
|  | Eulji University | [1167] |
|  | Ewha Woman's University | [168], [198], [1125], [1129] |
|  | Gachon University | [119], [473], [474], [475], [831] |
|  | Gwangju Women’s University | [160] |
|  | Gyeonggi Infectious Disease Control Center | [419] |
|  | Gyeonggido Business and Science Accelerator | [981] |
|  | Gyeongsang National University Hospital | [1175] |
|  | Hallym University | [359], [388], [414], [493], [725], [952], [1145], [1174] |
|  | Hanyang University | [103], [263], [530] |
|  | Health Insurance Review and Assessment Service | [29], [628], [948] |
|  | International Vaccine Institute | [204], [293] |
|  | Jeonbuk National University | [137], [270], [500], [646], [829] |
|  | Konkuk University | [50], [384], [1024] |
|  | Konyang University | [143], [967] |
|  | Kookmin University | [432], [391] |
|  | Korea Centers for Disease Control and Prevention | [18], [30], [201], [212], [255], [416], [498], [536], [1246] |
|  | Korea Institute of Oriental Medicine | [42], [174] |
|  | Korea Research Institute of Bioscience and Biotechnology | [507], [1070] |
|  | Korea Research Institute of Chemical Technology | [8], [325], [437], [560], [700], [1035], [1135] |
|  | Korea University | [54], [654], [772], [984], [1089], [1194] |
|  | Korea University College of Medicine | [450], [1251] |
|  | KR BioTech | [548] |
|  | Kyung Hee University | [998], [1154] |
|  | Kyung Hee University College of Medicine | [618] |
|  | Kyungpook National University | [457] |
|  | Libentech Co. | [352] |
|  | National Cancer Center | [155], [826] |
|  | National Medical Center | [60], [321], [471], [695], [965], [1155], [1249] |
|  | National University Hospital | [469] |
|  | Osong Medical Innovation Foundation | [1045] |
|  | Pohang University of Science and Technology | [335] |
|  | Pusan National University School of Medicine | [1122] |
|  | Samsung Medical Center | [120], [275], [404], [910] |
|  | Sejong University | [104] |
|  | Semyung University | [256] |
|  | Seoul Center for Infectious Disease Control and Prevention | [1064] |
|  | Seoul National Medical Center | [39] |
|  | Seoul National University | [260], [362], [395], [495], [584], [685], [726], [827], [1163] |
|  | Seoul National University College of Medicine | [211], [243], [666], [1005], [1087] |
|  | Seoul National University Hospital | [710], [757] |
|  | Seowon University | [458] |
|  | Sungkyunkwan University | [357], [1173] |
|  | Sungkyunkwan University School of Medicine | [157], [159], [194], [222], [393], [542], [621], [737], [746], [936], [1057], [1123] |
|  | University College of Medicine | [90], [227], [595] |
|  | University of Hong Kong | [12] |
|  | University of Science and Technology | [345] |
|  | University of Ulsan College of Medicine | [123], [139], [246], [467] |
|  | Yongin Mental Hospital | [1138] |
|  | Yonsei University | [115], [152], [195], [459], [1161], [1257] |
| **China** | Academy of Military Medical Sciences | [375], [532], [872], [875], [900], [1176] |
|  | Beijing Institute of Microbiology and Epidemiology | [66], [75], [77], [170], [262], [409], [735], [852], [997], [1072] |
|  | Beijing Institute of Pharmacology and Toxicology | [313], [366], [1124] |
|  | Beijing Institute of Radiation Medicine | [963] |
|  | Capital Medical University | [570] |
|  | Central South University | [540], [885], [1233] |
|  | Chinese Academy of Agricultural Sciences | [76], [609] |
|  | Chinese Academy of Inspection and Quarantine | [865] |
|  | Chinese Academy of Medical Sciences & Peking Union Medical College | [302], [426], [427], [552], [573], [1029] |
|  | Chinese Academy of Sciences | [20], [58], [86], [165], [368], [780], [836], [980], [1059], [1113], [1114] |
|  | Chinese Center for Disease Control and Prevention | [203], [412], [812], [886], [972], [1150], [1234] |
|  | Chinese University of Hong Kong | [951], [1188] |
|  | Fudan University | [237], [406], [449], [663], [843], [923], [1116], [1151] |
|  | Gannan Medical Universitygrid | [1104] |
|  | Guangdong Provincial Center for Disease Control and Prevention | [273], [400], [[597] |
|  | Guangdong Provincial Key Laboratory of Tropical Disease Research | [1230] |
|  | Guangxi University | [842] |
|  | Guangzhou Institute of Respiratory Health | [615], [1182] |
|  | Guangzhou University | [26], [1245] |
|  | Hebei Medical University | [647], [996], [1141] |
|  | Hong Kong Baptist University | [568] |
|  | Hong Kong Polytechnic University | [835] |
|  | Hong Kong Polytechnic University | [1247] |
|  | Hospital of Shenzhen University Health Science Center | [1038] |
|  | Huazhong Agricultural University | [425] |
|  | Hubei University | [860], [895], [652] |
|  | Huizhou Central Hospital | [221] |
|  | Hunan University | [558] |
|  | Jiaxing Municipal Center for Disease Control and Prevention | [63] |
|  | Jilin Agricultural University | [11] |
|  | Key Laboratory of the Ministry of Health for Research on Quality and Standardization of Biotech Product | [354] |
|  | Nanchang University | [1101], [1115] |
|  | National Institute for Viral Disease Control and Prevention | [35], [1140], [1196] |
|  | National Institute of Metrology | [974] |
|  | National Institutes for Food and Drug Control | [57] |
|  | North University of China | [19] |
|  | Northeast Forestry University | [753] |
|  | Northwest A&F University | [930] |
|  | Peking Union Medical College | [110], 250] |
|  | Peking University Fifth School of Clinical Medicine | [468] |
|  | School of Laboratory Medicine and Life Science and Key Laboratory of Medical Virology | [9] |
|  | Shandong University | [382], [421], [742], [743] |
|  | Shanghai Jiao Tong University | [581] |
|  | Shanxi University | [503] |
|  | South China Agricultural University | [64], [206] |
|  | Southern Medical University | [121] |
|  | State Key Laboratory of Emerging Infectious Diseases | [91], [92], [785] |
|  | Sun Yat-sen University | [511] |
|  | Tiantai County People's Hospital | [1069] |
|  | Tsinghua University | [300], [306], [571], [1071], [1098], [1102], [1106], [1108] |
|  | University of Hong Kong | [4], [22], [67], [82], [131], [177], [183], [207], [232], [261], [283], [316], [344], [373], [387], [499], [534], [545], [550], [561], [657], [660], [682], [714], [717], [718], [781], [804, [813], [855], [915], [920], [953], [979], [992], [1051], [1003], [1090], [1142], [1147], [1225], [1226], [1255] |
|  | University of Science and Technology Beijing | [27], [65] |
|  | University of Science and Technology of China | [301] |
|  | Wenzhou Medical University | [961], [994] |
|  | Wuhan Institute of Virology | [286], [348], [562] |
|  | Wuhan University | [230], [755], [933] |
|  | Xiamen University | [79], [1204] |
|  | Zhejiang Provincial Centre for Disease Control and Prevention | [244] |
|  | Zhejiang University | [45], [284] |
| **Germany** | AItona Diagnostics GmbH | [1248] |
|  | Bundeswehr Institute of Microbiology | [947] |
|  | Centre for Experimental and Clinical Infection Research | [1253] |
|  | Charité-Universitätsmedizin Berlin | [488], [1216] |
|  | German Centre for Infection Research (DZIF) | [626] |
|  | German Center for Infection Research | [689], [691] |
|  | German Primate Center-Leibniz Institute for Primate Research | [577], [632], [679], [1211] |
|  | Goethe University | [964], [686] |
|  | Institute for Infectious Diseases and Zoonoses | [189], [371] |
|  | Johannes Gutenberg University | [601], [924] |
|  | Justus Liebig University | [5], [25], [176] |
|  | Labor Prof. Gisela Enders MVZ GbR | [604] |
|  | Leibniz Institute for Primate Research | [487], [1086] |
|  | LMU University of Munich | [817], [960] |
|  | Max Planck Institute of Psychiatry | [1075] |
|  | Paul Ehrlich Institut | [55], [531] |
|  | Philipps University of Marburg | [310], [611] |
|  | University Duisburg-Essen | [68] |
|  | University Medical Center Hamburg-Eppendorf | [913], [1032] |
|  | University of Bonn Medical Centre | [116], [127], [128], [202], [220], [655], [715], [779], [909], [942], [1030], [1209], [1219], [1250] |
|  | University of Göttingen | [857], [919] |
|  | University of Lübeck | [304], [1264] |
|  | University of Veterinary Medicine Hannover Foundation | [336], [454] |
| **Netherlands** | Erasmus Medical Center | [72], [100], [154], [209], [249], [317], [361], [479], [659], [681, [774], [792], [878], [899], [999], [1020], [1047], [1054], [1084], [1210] |
|  | Leiden University Medical Center | [85], [278], [282], [464], [721], [1043], [1165] |
|  | National Institute for Public Health and the Environment | [483], [770], [805], [1221] |
|  | Netherlands Centre for Infectious Disease Control | [509] |
|  | Royal Netherlands Academy of Arts and Sciences and University Medical Center | [1] |
|  | University of Amsterdam | [879] |
|  | Utrecht University | [572], [759], [1195], [1213] |
| **Japan** | Hokkaido University | [575] |
|  | Juntendo University | [631] |
|  | Nagahama Institute of Bio-Science and Technology | [365] |
|  | National Institute of Infectious Disease | [97], [205], [337], [351], [356], [557], [608], [791], [799], [800], [818], [868], [1212] |
|  | Nihon University | [445], [1105] |
|  | Osaka University | [238], [716] |
|  | Shiga University of Medical Science | [811] |
|  | Shizuoka University | [939] |
|  | Tohoku University Graduate School of Medicine | [271] |
|  | University of Tokyo | [144], [333], [430], [569], [932], [950], [988] |
|  | Yokohama City University | [358] |
| **United Kingdom** | Animal and Plant Health Agency (APHA) | [135] |
|  | Centre for Infectious Disease Surveillance and Control | [429] |
|  | Defence Science and Technology Laboratorygrid | [257] |
|  | Glasgow University | [1017] |
|  | Heart of England NHS Foundation Trust | [93] |
|  | Imperial College London | [638], [823] |
|  | London School of Hygiene & Tropical Medicine | [1199] |
|  | Manchester University NHS Foundation Trust | [764] |
|  | Middlesex University | [130] |
|  | National Institute for Biological Standards and Control | [259] |
|  | Public Health England | [411], [1074] |
|  | Swansea University | [1144] |
|  | University Hospitals Leicester NHS Trust | [1007] |
|  | University of Cambridge | [280], [931] |
|  | University of Kent | [117] |
|  | University of Oxford | [192], [312], [599], [1031] |
|  | University of Reading | [52] |
|  | Wellcome Trust Sanger Institute | [1088], [1218] |
| **Egypt** | Ain Shams University | [1091] |
|  | Al-Azhar University | [1136] |
|  | Damietta University | [485] |
|  | Egyptian Atomic Energy Authority (EAEA) | [328] |
|  | Egyptian Ministry of Health and Population (MOHP) | [299] |
|  | Mansoura University | [1012] |
|  | National Center for Radiation Research and Technology (NCRRT) | [1232] |
|  | National Research Centre (NRC) | [74], [163], [298], [766], [797], [938], [1027], [1139] |
|  | Pharmaceutical and Drug Industries Research Institute | [650] |
|  | Port Said University | [173], [613], [1095] |
|  | World Health Organization Regional Office for the Eastern Mediterranean | [734] |
|  | Zagazig University | [216] |
| **France** | Aix-Marseille Université | [169] |
|  | Cirad UPR AGIRs | [1015] |
|  | Ecole Normale Supérieure de Lyon | [258] |
|  | Inserm france | [146] |
|  | Institut Pasteur de Lille | [242], [520], [643], [1158], [1217], [1241] |
|  | Paris Diderot University | [224] |
|  | Sorbonne Universités | [973] |
|  | Unité de coordination opérationnelle du risque épidémique et biologique | [1169] |
|  | Unité des Maladies Infectieuses et Tropicales | [390] |
|  | University of Lille | [10], [1009] |
|  | Université de Lille 2 | [219], [369], [664] |
|  | Université de Montpellier | [853] |
| **Australia** | Australian Institute of Tropical Health & Medicine | [617] |
|  | CSIRO Australian Animal Health Laboratory | [89], [453] |
|  | Charles Sturt University | [744] |
|  | James Cook University | [859] |
|  | Monash University | [671], [1166] |
|  | National Centre for Immunisation Research and Surveillance of Vaccine Preventable Diseases (NCIRS) | [147] |
|  | University of New South Wales | [78] |
|  | University of Queensland | [71], 199] |
|  | University of Sydney | [514], [970] |
|  | University of New South Wales | [136], [245], [424], [1023], [1186], [1243] |
|  | University of Tasmania | [405] |
| **Taiwan** | Academia Sinica | [403], [574], [703], [863] |
|  | Kaohsiung Medical University | [492] |
|  | National Chung Hsing University | [2], [307], [389], [1117] |
|  | National Taiwan University | [403], [593], [703], [809], [877] |
|  | National Yang-Ming University | [289], [372], [921], [1099], [1207] |
|  | Soochow University | [697], [1236] |
| **India** | Adamas University | [1097] |
|  | Anna University, Bharathidasan Institute of Technology (BIT) Campus | [639] |
|  | Council of Scientific and Industrial Research (CSIR) | [267], [935] |
|  | Dinabandhu Andrews College | [73] |
|  | Era University | [480] |
|  | Guru Nanak Institute of Pharmaceutical Science and Technology | [898] |
|  | Indian Council of Medical Research (ICMR) | [254], [465] |
|  | Indian Institute of Science Education and Research  (IISER-TVM) | [1205] |
|  | Integral University | [1258] |
|  | Mangalayatan University | [1103] |
|  | Manipal University | [547] |
|  | National Institute of Technology | [266] |
|  | Presidency University | [460] |
|  | Sree Buddha College of Engineering | [705] |
|  | Tezpur University | [602] |
| **United Arab Emirates** | Abu Dhabi Department of Health | [1016], [1056] |
|  | Abu Dhabi Food Control Authority | [374], [420], [946] |
|  | Central Veterinary Research Laboratory | [69], [95] |
|  | College of Medicine and Health Sciences | [208], [482] |
|  | Department of the President's Affairs | [167], [874] |
|  | Health Authority Abu Dhabi | [1010] |
|  | Mafraq Hospital | [784] |
| **Iran** | Babol University of Medical Sciences | [138] |
|  | Baqiyatallah University of Medical Sciences | [355] |
|  | Kerman University of Medical Sciences | [417] |
|  | Shahid Beheshti University of Medical Sciences | [515], [1193] |
|  | Tabriz University of Medical Sciences | [343] |
|  | Tehran University of Medical Sciences | [630], [231] |
|  | University of Tehran | [837] |
|  | Urmia University of Medical Sciences | [14], [309], [1094], [1170], [1197] |
| **Spain** | Campus de la Universitat Autònoma de Barcelona (UAB) | [235], [438], [1231] |
|  | Campus Universidad Autónoma de Madrid | [407], [745], [788] |
|  | Centre de Recerca en Sanitat Animal (CReSA, IRTA-UAB) | [172], [413], [692], [787] |
|  | Centro Nacional de Biotecnología (CNB-CSIC) | [506], [677], [722], [821] |
|  | University of Barcelona | [315] |
|  | University of Las Palmas de Gran Canaria | [941] |
| **Canada** | Public Health Agency of Canada | [741], [985] |
|  | Université de Sherbrooke | [191] |
|  | University of Alberta | [1000], [1153] |
|  | University of British Columbia | [394] |
|  | University of Guelph | [41], [484] |
|  | University of Manitoba | [303] |
|  | University of Saskatchewan | [494], [641], [1046] |
|  | University of Toronto | [922], [926] |
|  | Wilfrid Laurier University | [844] |
| **Qatar** | Hamad Medical Corporation | [392], [801] |
|  | Ministry of Public Health | [543], [881], [1131] |
|  | Qatar University | [251], [590], [1081] |
|  | Supreme Council of Health | [528] |
| **Switzerland** | Humabs BioMed SA | [982] |
|  | Institute of Virology and Immunology (IVI) | [53], [428] |
|  | SPIEZ LABORATORY | [606] |
|  | University of Bern | [861] |
|  | University of Zurich | [756], [1254] |
| **Kenya** | Jomo Kenyatta University of Agriculture and Technology | [504] |
|  | Food and Agriculture Organization of the United Nations (FAO), Kenya | [707], [1021] |
|  | International Livestock Research Institute, Old Naivasha Road | [775] |
|  | Ministry of Agriculture, Livestock and Fisheries | [670] |
|  | Washington State University Global Health Program | [525] |
|  | US Centers for Disease Control and Prevention | [699], [867] |
| **Pakistan** | Government College University | [269], [987] |
|  | Islamabad Diagnostic Center | [440] |
|  | National Institute of Health | [762] |
|  | University of Agriculture | [762] |
|  | University of Engineering and Technology (UET) | [264] |
|  | University of Lahore | [112] |
| **Singapore** | Agency for Science, Technology and Research | [894] |
|  | Gerard Kian-Meng Goh, Goh's BioComputing | [934] |
|  | National University of Singapore | [101], [918] |
|  | Nanyang Technological University | [133], [461], [712] |
| **Italy** | Istituto Superiore di Sanità | [181] |
|  | Istituto Zooprofilattico Sperimentale Lombardia ed Emilia Romagna | [332] |
|  | Molecular Horizon srl | [292] |
|  | Scientific Institute IRCCS E. Medea | [466], [1171] |
|  | University of Rome "Tor Vergata" | [1148] |
| **Jordan** | Jordan Ministry of Health | [538] |
|  | Jordan University of Science and Technology | [995] |
|  | National Center for Immunization and Respiratory Diseases | [1096] |
|  | University of Jordan | [268], [806] |
| **Greece** | Hellenic Center for Disease Control and Prevention | [38] |
|  | Hellenic Pasteur Institute | [674] |
|  | Medical School, 75 Mikras Asias Street | [1252] |
|  | University of Patras | [864] |
| **Turkey** | Canik Community Health Center | [672] |
|  | Gulhane Medical Academy | [624] |
|  | Hitit University | [653] |
|  | Turkish-German University | [1120] |
| **Bangladesh** | Disease Control & Research (IEDCR) | [448] |
|  | Jahangirnagar University | [591] |
|  | University of Chittagong | [324] |
|  | University of Rajshahi | [329] |
| **Thailand** | Chulalongkorn University | [854] |
|  | Kasetsart University | [1111] |
|  | Ministry of Public Health | [598], [684] |
| **Kuwait** | Kuwait University | [31], [905] |
|  | Kuwait Institute for Scientific Research | [124] |
| **Lebanon** | American University of Beirut Medical Center | [156], [1244] |
| **Mali** | University of Sciences, Techniques and Technologies of Bamako | [727] |
| **Austria** | University of Veterinary Medicine Vienna | [637], [731], [768] |
| **Belgium** | Ghent University Hospital | [378] |
|  | Laboratory of Virology and Chemotherapy | [1200] |
|  | University Hospitals Leuven | [833] |
| **Brazil** | Federal University of Rio de Janeiro | [334], [1149] |
| **Cambodia** | Institut Pasteur du Cambodge | [501] |
| **Ghana** | Kwame Nkrumah University of Science and Technology | [546], [928] |
| **Indonesia** | Indonesia International Institute for Life Sciences | [589] |
|  | Prof. Dr. Sulianti Saroso Infectious Disease Hospital | [339] |
| **Iraq** | University of Diyala | [297] |
|  | University of  Basrah | [1050] |
| **Israel** | Kimron Veterinary Institute | [816] |
|  | Tel Aviv University | [489] |
| **Kazakhstan** | Institute of veterinary medicine and animal husbandry | [1192] |
|  | Scientific Production Center of Microbiology and Virology | [1062] |
|  | Zhambylskaya Oblast | [1061] |
| **Malysia** | Institute for Medical Research | [876] |
| **Morocco** | Institut Pasteur Morocco | [385] |
|  | Université Hassan II, Casablanca | [771] |
| **Nigeria** | Salem University | [927] |
| **Sweden** | Umeå University | [940] |
| **Sudan** | Sudan Diabetic Childhood Center | [48] |
| **South Africa** | University of Pretoria | [59] |
|  | University of the Western Cape | [1168] |
| **Russia** | Lomonosov Moscow State University | [1256] |
|  | Siberian Federal University, Krasnoyarsk | [241] |
|  | State Research Center of Virology and Biotechnology "Vector" | [1132] |
| **Poland** | Jagiellonian University | [544] |
|  | University of Warsaw | [897] |
| **Oman** | Ministry of Health | [896] |
| **Sri Lanka** | International Organization for Migration (IOM), Colombo, Sri Lanka | [377] |
| **Ireland** | Trinity College Dublin | [634] |
| **Denmark** | World Health Organization (WHO) Regional Office for Europe | [673] |
| **Finland** | National Institute for Health and Welfare (THL) | [776] |
| **Argentina** | Universidad de Buenos Aires | [841] |
| **Vietnam** | Phan Chau Trinh University of Medicine | [978] |
| **Colombia** | National University of Colombia | [1026] |
| **New Zealand** | University of Auckland | [1198] |
| **Estonia** | University of Tartu | [1201] |
| **Portugal** | Instituto Nacional de Saúde Doutor Ricardo Jorge | [1240] |
| **Philippines** | Department of Health, Sta Cruz, Manila, Philippines | [274] |

List of funding agencies that have supported MERS-related research globally (n=337)

| **Country** | **Funding agency** | **Appendix S1: Technical Appendix Reference** |
| --- | --- | --- |
| **Multinational Agencies** | CGIAR Research Program on Agriculture for Nutrition and Health | [1060] |
|  | Coalition for Epidemic Preparedness Innovations (CEPI) | [199] |
|  | Global Challenges Research Fund (GCRF) | [1017] |
|  | European Center for Disease Prevention and Control (ECDC) | [479] |
|  | European Union projects | [95], [100], [116], [127], [128], [135], [154], [169], [172], [202], [209], [219], [220], [235], [303], [304], [336], [361], [407], [413], [428], [438], [454], [488], [509], [520], [549], [550], [626], [638], [643], [651], [655], [681], [692], [713], [715], [718], [722], [745], [751], [757], [774], [775], [779], [787], [788], [792], [805], [817], [821], [842], [864], [899], [940], [942], [964], [973], [1004], [1020], [1030], [1047], [1051], [1052], [1054], [1055], [1057], [1075], [1084], [1123], [1165], [1194], [1195], [1200], [1213], [1219], [1226], [1231], [1238], [1241], [1250], [1264] |
|  | World Health Organization (WHO) | [456] |
| **United States of America** | Aethlon Medical Inc. | [686] |
|  | Alberta Ministry of Economic Development, Trade, and Tourism Major Innovation | [1000], [1153] |
|  | American Leprosy Missions, USA | [931] |
|  | Barnes Jewish Hospital Foundation | [6], [7] |
|  | Baxter | [147], [514] |
|  | Bill and Melinda Gates Foundation | [277], [431], [823], [1118], [1204], [1241] |
|  | BioFire Diagnostics | [849] |
|  | Biomedical Advanced Research and Development Authority (BARDA) | [257], [1034] |
|  | Burroughs Wellcome Fund | [230], [856], [1118], [1121], [1238] |
|  | Campaign Urging Research for Eosinophilic Diseases Foundation (M.E.R.) | [6], [7] |
|  | Center for Biodefense Emerging Infectious Diseases (CBEID) | [49], [901] |
|  | Center of Excellence for Influenza Research and Response (CEIRS) | [100], [154] |
|  | Cerus Corporation | [107] |
|  | Colorado State University | [380], [623], [969], [1001], [1002], [1029] |
|  | Commonwealth of Pennsylvania | [570] |
|  | Cystic Fibrosis Foundation | [962] |
|  | Emory University | [527] |
|  | Florida International University | [158] |
|  | Galveston National Laboratory (GNL) | [901] |
|  | GeneOne Life Science | [1033] |
|  | George Mason University (C.S.C.) | [6], [7] |
|  | Georgia State University | [702] |
|  | Harvard University | [12], [1228] |
|  | Hauptman-Woodward Medical Research Institute | [1119] |
|  | Johns Hopkins University | [187] |
|  | Merck | [147], [514] |
|  | MSD Oncology | [892] |
|  | National Center for Functional Glycomics | [1037] |
|  | National Institute of Health (NIH) | [83], [88], [98], [99], [106], [109], [126], [129], [134], [141], [144], [163], [164], [171], [175], [180], [185], [188], [230], [240], [242], [248], [252], [253], [258], [260], [272], [279], [281], [285], [294], [305], [311], [314], [326], [349], [363], [370], [373], [375], [379], [382], [387], [397], [399], [402], [407], [408], [409], [410], [421], [431], [439], [446], [452], [481], [490], [496], [499], [512], [513], [526], [527], [539], [541], [553], [559], [564], [567], [570], [575], [578], [583], [585], [596], [610], [616], [622], [633], [635], [636], [642], [644], [645], [647], [648], [649], [663], [681], [687], [688], [693], [696], [701], [708], [713], [717], [718], [722], [723], [724], [728], [744], [745], [747], [748], [751], [758], [765], [766], [771], [782], [783], [788], [790], [797], [798], [806], [807], [808], [819], [823], [830], [839], [834], [846], [847], [848], [855], [856], [857], [858], [862], [870], [873], [882], [900], [901], [902], [903], [906], [911], [932], [954], [955], [956], [957], [958], [959], [962], [971], [983], [988], [990], [991], [993], [994], [1001], [1002], [1011], [1015], [1017], [1025], [1027], [1029], [1034], [1036], [1037], [1038], [1044], [1051], [1052], [1053], [1071], [1072], [1073], [1090], [1092], [1093], [1100], [1107], [1109], [1112], [1118], [1119], [1121], [1126], [1139], [1146], 1156], 1157], [1164], [1172], [1177], [1181], [1183], [1187], [1199], [1203], 1208], [1217], [1224], [1226], [1227], [1238], [1239], [1241], [1259], [1261] |
|  | New York Blood center | [49], [77], [84], [563], [564], [647], [649], [885], [1072] |
|  | North Carolina Coronavirus Relief Fund | [862] |
|  | Novavax Inc. | [749] |
|  | Pfizer | [147], 514] |
|  | Purdue Center for Cancer Research | [185] |
|  | Regenerative Medicine Minnesota | [858] |
|  | Regeneron Pharmaceuticals, Inc. | [551], [929] |
|  | Romark | [147], [514] |
|  | Saint Louis University | [1239] |
|  | Seed University | [1028] |
|  | U.S. Centers for Disease Control and Prevention | [32], [149], [190], [213], [229], [299], [383], [525], [625], [670, [699], [711], [795], [820], [867], [1018], [1019], [1056], [1016], [1085], [1220] |
|  | U.S. Department of Defense's Defense Threat Reduction Agency | [33], [331], [567], [790], [867], [1011], [1025], [1061], [1093] |
|  | U.S. Department of Energy by Lawrence Livermore National Laboratory | [803], [1119], [1238] |
|  | United States Agency for International Development (USAID) | [298], [484], [490], [499], [501], [527], [707], [790], [915], [1021], [1227] |
|  | United States Army Medical Research Institute of Infectious Diseases | [1077] |
|  | United States Department of Agriculture | [1109] |
|  | United States Department of the Army | [1033] |
|  | University of Kansas | [728] |
|  | University of Michigan Medical School | [884] |
|  | University of North Dakota | [884] |
|  | University of Pennsylvania | [728] |
|  | University of Pittsburgh's National Institutes of Health Cancer Center | [586] |
|  | University of Texas | [171], [496], [808], [906], [1164] |
|  | University of Washington | [230], [1118], [1238] |
|  | University Seed | [208] |
|  | US Civilian Research & Development Foundation (CRDF Global) | [268] |
|  | US Department of Health and Human Service | [186], [1037], [1137] |
|  | US Food and Drug Administration | [108] |
|  | US Global Disease Detection Operations Center Outbreak Response | [516], [538], 1096] |
|  | US National Science Foundation | [380] |
|  | Walther Cancer Foundation | [185], [687], [1146] |
|  | Washington National Primate Research Center | [1224] |
|  | Washington University | [6], [7] |
|  | Welch Foundation | [990] |
| **Saudi Arabia** | Imam Abdulrahman Bin Faisal University (IAU) | [698] |
|  | King Abdulaziz City for Science and Technology (KACST) | [56], [125], [142], [145], [184], [265], [279], [295], [327], [330, [341], [346], [441], [442], [462], [505], [508], [510], [529], [576], [588], [592], [605], [678], [696], [880], [1039], [1063], [1078], [1080], [1178], [1180] |
|  | King Abdulaziz University | [233], [340], [502], [587], [866] |
|  | King Abdullah International Medical Research Center (KAIMRC) | [122], [151], [193], [451], [478], [556], [640], [661], [665, [908], [1066], [1222], [1237] |
|  | King Abdullah University of Science and Technology (KAUST) | [21], [986], [1215] |
|  | King Fahad Armed Forces Hospital | [579] |
|  | King Fahad Medical City | [108], [401], [607], [739], [840] |
|  | King Faisal Specialist Hospital and Research Centre | [105], [763], [1080] |
|  | King Faisal University | [70], [118], [247], [328], [486], [838], [1134], [1226] |
|  | King Saud bin Abdulaziz University for Health Sciences | [1237] |
|  | king Saud University | [228], [327], [422], [433], [466], [603], [709], [773], [786], [796], [814], [815], [828], [888], [907], [944], [1041], [1048], [1171] |
|  | Ministry of Education | [566], [736] |
|  | Ministry of Health | [32], [149], [423], [625], [752], [795], [820], [942], [945], [1008], [1018], [1019], [1079], [1088], [1218] |
|  | Municipality of the Holy City of Makkah | [651] |
|  | Prince Sultan University | [269], [943] |
|  | Princess Nourah bint Abdulrahman University | [287] |
|  | Qassim University | [318] |
|  | Qassim University | [690] |
|  | Saudi Ministry of Environment, Water and Agriculture (MEWA) | [193] |
|  | Sulaiman Al Rajhi University | [225] |
|  | Taibah University | [738] |
|  | Taif University | [46], [1095], [1189] |
|  | Umm Al-Qura University | [173], [976] |
|  | University of Tabuk | [1065] |
| **Republic of Korea** | Korea University | [654], [1089] |
|  | Chungcheongbuk-do Value Creation Project (Plexense) | [1045] |
|  | Chungnam National University | [524] |
|  | Gyeonggi Infectious Disease Control Center | [949] |
|  | Gyeonggi provincial government | [981] |
|  | Konkuk University | [384] |
|  | Korea Centers for Disease Control and Prevention | [18], [104], [119], [227], [383], [437], [469], [498], [910], [981], [1064], [1129], [1194], [1246] |
|  | Korea Health Promotion Institute | [119] |
|  | Korea National Institute of Health | [18], [201], [1202], [1246], [750], [1045] |
|  | Korean Centers for Disease Control and Prevention (KCDC) | [437] |
|  | Korean Government | [432], [467], [507], [1253] |
|  | KRICT - Korea Research Institute of Chemical Technology | [1035] |
|  | Kyung Hee University | [618] |
|  | Ministry of Education, Science, and Technology | [50], [115], [137], [150], [198], [270], [380], [414], [500], [829], [1110], [1163], [1174], [1175] |
|  | Ministry of Food and Drug Safety | [750] |
|  | Ministry of Health & Welfare | [120], [139], [148], [150], [153], [195], [204], [211], [288], [293], [321], [357], [386], [388], [450], [467], [471], [497], [522], [548], [554], [582], [618], [694], [695], [750], [824], [825], [829], [965], [969], [998], [1005], [1049], [1070], [1087], [1125], [1133], [1251], [1260] |
|  | Ministry of Science, ICT, and Future Planning (MSIP) | [8], [42], [43], [54], [70], [103], [137], [174], [246], [288], 325], [335], [345], [357], [347], [367], [455], [485], [493], 522], [555], [560], [646], [700], [725], [952], [984], [1045], [1070], [1076], [1135], [1145], [1175] |
|  | Ministry of Trade, Industry & Energy | [685] |
|  | National Cancer Center in South Korea | [1162], [1174] |
|  | National Medical Center Research Institute | [60], [150], [1249] |
|  | National Research Foundation of Korea (NRF) | [195], [380], [495], [500], [694], [969], [981], [1049], [1133], [1174], [1163**]** |
|  | Oklahoma State University | [106] |
|  | Pusan National University Hospital | [1122] |
|  | Research Program of Rural Development Administration | [396] |
|  | Rural Development Administration | [883] |
|  | Samsung Biomedical Research Institute (SBRI) | [157], [194], [275], [393], [542], [936], [1123], |
|  | Samsung Science & Technology Foundation | [391] |
|  | Seoul Metropolitan Government | [1064**]** |
|  | Seoul National University | [362], [395], [666], [827] |
|  | Sungkyunkwan University | [1173] |
|  | Yongin Mental Hospital, 245 beon-gil, Jangan-gu, Suwon, Gyeonggi Province South Korea | [1138] |
|  | Yonsei University College of Medicine | [90], [115], [459] |
|  | Zhihui Zhengzhou - 1125 Talent Gathering Plan | [930] |
| **China** | Beijing Natural Science Foundation | [27] |
|  | Central South University | [540], [1233] |
|  | China Mega-Project for Infectious Diseases Control and Prevention | [9], [49], [57], [75], [110], [170], [203], [237], [262], [409], [412], [421], [562], [649], [735], [742], [865], [885], [961], [994], [996], [997], [1106], [1140], [1141], [1150], [1196], [1225], [1234] |
|  | Chinese Academy of Medical Sciences | [406], [558], [573] |
|  | Chinese Academy of Science | [165], [348], [562], [301], **[**972], [980], [1071] |
|  | Food and Health Bureau of Hong Kong SAR | [67], [550], [992], [1003], [1015], [1226] |
|  | Government of the Hong Kong Special Administrative Region | [4], [69], [79], [82], [91], [95], [177], [183], [260], [279], [283, [387], [545], [561], [657], [726], [780], [785], [804], [813], [920], [979], [1090], [1142], [1225], [1247], [1255] |
|  | Guangdong Laboratory for Lingnan Modern Agricultural Science and Technology | [45] |
|  | Guangdong Provincial Science and Technology | [121], [615], [1230] |
|  | Hong Kong Health and Medical Research Fund (HMRF) | [183], [232] |
|  | Hong Kong Polytechnic University | [1247] |
|  | Hong Kong Research Grants Council | [58] |
|  | Hubei University of Chinese Medicine | [652] |
|  | Hunan Government | [540], [842], [1233] |
|  | Ministry of Education | [183], [207], [261], [634], [780], [781], [1003], [1104] |
|  | Ministry of Environmental Protection | [382] |
|  | Ministry of Health | [110], [597], [812], [1230] |
|  | Ministry of Science and Technology | [20], [58], [86], [177], [221], [303], [313], [354], [368], [427, [504], [526], [532], [571], [663], [693], [743], [809], [812, [872], [877], [886], [900], [923], [961], [1059], [1102], [1113], [1115], [1124], [1147], [1176], [1182] |
|  | National 973 Program of China | [49], [79], [84], [563], [647], [959], [996], [1140], [1141] |
|  | National Cancer Center | [530] |
|  | National Health and Family Planning Commission | [250], [427], [886] |
|  | National Key Research and Development Program of China | [9], [11], [65], [66], [76], [170], [203], [237], [262], [284], [301], [302], [306], [382], [412], [426], [526], [540], [558], [735], [836], [875], [895], [961], [974], [1071], [1108], [1151], [1182, [1196], [1233], [1234] |
|  | National Natural Science Foundation of China (NSFC) | [9], [11], [19], [27], [35], [45], [58], [64], [86], [121], [165], [170], [206], [230], [237], [262], [284], [302], [313], [366], [382], [406], [409], [421], [436], [449], [511], [526], [530], [540], [550], [552], [558], [568], [570], [581], [609], [652], [662], [735], [755], [780], [842], [852], [860], [875], [884], [886], [895], [959], [961], [963], [972], [979], [996], [1059], [1098], [1101], [1104], [1106], [1108], [1114], [1116], [1124], [1147], [1151], [1182], [1204], [1230], [1233], [1245] |
|  | Peking Union Medical College | [573] |
|  | Program of Shanghai Academic/Technology Research Leader | [1151] |
|  | Providence Foundation Limited | [953] |
|  | Science and Technology Project of Zhejiang Province | [63] |
|  | Shanghai Public Health Clinical Center | [843] |
|  | State Key Laboratory of Pathogen and Biosecurity | [66], [735] |
|  | State Key Laboratory of Respiratory Disease | [375], [753], [1147] |
|  | Tam Wah Ching Research Endowment | [682] |
|  | University of Hong Kong | [4], [69], [79], [91], [92], [95], [131], [316], [344], [534], [545], [660], [680], [714], [781], [785], [804], [813], [920], [979], [992], [1003], [1090] |
|  | Wuhan University | [230] |
|  | Zhejiang University | [540], [1233] |
| **Germany** | Cardio-Pulmonary Institute (CPI) | [310] |
|  | Deutsche Forschungsgemeinschaft (the German Research Council) | [5], [6], [7], [55], [95], [116], [128], [176], [220], [546], [550], [611], [626], [651], [689], [715], [757], [779], [805], 867], [928], [942], [960], [964], [1030], [1055], [1075], [1211], [1216], [1219], [1250] |
|  | Deutsche Gesellschaft für Internationale Zusammenarbeit | [947] |
|  | Dutch Organization for Health Research and Innovation | [573] |
|  | Federal Ministry of Education and Research (Bundesministerium für Bildung und Forschung) | [5], [116], [128], [189], [220], [259], [415], [428], [488], [487], [577], [632], [651], [779], [919], [942], [1012], [1075], [1086], [1219] |
|  | Galveston National Laboratory (C.K.T) | [649] |
|  | German Center for Lung Research (DZL) | [310] |
|  | German Centre for Infection Research (DZIF) | [5], [55], [116], [163], [176], [202], [220], [304], [310], [651], [655], [691], [817], [909], [913], [942], [960], [964], [1004], [1032], [1264] |
|  | Hartmut Hoffmann-Berling International Graduate School of Molecular & Cellular Biology (HBIGS), Heidelberg, Germany | [186] |
|  | Institute for Infectious Diseases and Zoonoses | [371] |
|  | Leibniz graduate school Emerging Infectious Diseases (EIDIS) | [577] |
|  | LOEWE Research Center DRUID | [176] |
|  | Marc Strobel, CVC Capital Partners | [601], [924] |
|  | Ministry of Science and Culture of Lower Saxony, Germany | [72] |
|  | Projekt DEAL | [25] |
|  | University of Göttingen | [241] |
|  | University of Zurich | [756], [1254] |
|  | Universitätsmedizin Berlin | [6], [7] |
|  | Volkswagen Foundation | [1075] |
| **Netherlands** | Erasmus Graduate Program Infection & Immunity | [317], 851] |
|  | Leiden University Medical Center | [1043] |
|  | Nederlandse Organisatie voor Wetenschappelijk Onderzoek (Dutch Research Council) | [879] |
|  | Ministry of Education, Culture and Science | [1] |
|  | Netherlands Organization for Health Research and Development | [317], [999], [1210] |
|  | Netherlands Organization for Scientific Research | [85], [249], [303], [378], [464], [721], [759], [922], [1043], [1165], [1238], [1252] |
|  | Netherlands’ Ministry of Health,  Welfare, and Sport | [483] |
| **Japan** | Astellas Foundation for Research on Metabolic Disorders | [799] |
|  | Grants-in-Aid for Scientific Research (KAKENHI) | [939] |
|  | Japan Agency for Medical Research and Development | [97], [132], [144], [205], [271], [351], [356], [358], [430], [569], [716], [800], [868], [988], [1212], [1235**]** |
|  | Japan Agency for Medical Research and Development (AMED) | [238] |
|  | Japan Science and Technology Agency (JST) CREST program | [144], [430], [575], [950], [988] |
|  | Japan Society for the Promotion of Science | [144], [205], [333], [356], [430], [791], [811], [818], [932], [950], [988], [1235] |
|  | Japanese Respiratory Foundation | [557] |
|  | Ministry of Education, Culture, Sports, Science and Technology of Japan | [97], [132], [358], [365], [445], [569], [631], [799], [1076], [1212] |
|  | Ministry of Health, Labor, and Welfare | [97], [337], [351], [608], [631], [799], [800], [868], [1212] |
|  | Naito Foundation | [557] |
|  | National Center for Global Health and Medicine | [97], [1212] |
|  | National Institute of Infectious Diseases | [608] |
|  | Nihon University School of Dentistry | [1105] |
|  | Research Foundation for Opto-Science and Technology | [557] |
|  | RISTEX program for Science of Science, Technology and Innovation Policy | [144], [430], [988] |
|  | Takeda Science Foundation | [716] |
|  | Tokyo Society of Medical Sciences | [988] |
|  | University of Tokyo | [333] |
| **United Kingdom** | Antibiotic Research UK | [931] |
|  | AstraZeneca | [12] |
|  | Department of Health and Social Care (DHSC) | [80] |
|  | GSK | [147], [514] |
|  | Imperial College London | [599] |
|  | Pirbright Institute | [71] |
|  | Public Health England | [429] |
|  | UK Department for Environment, Food and Rural Affairs (Defra) | [135] |
|  | UK National Institute for Health Research | [130], [1031], [1218], [1219] |
|  | United Kingdom Biotechnology and Biological Sciences Research Council | [144], [931], [988] |
|  | University College London Hospitals Biomedical Research Centre | [116], [1218], [1219] |
|  | University of Oxford | [599] |
|  | VIRGO consortium | [792] |
|  | Wellcome Trust | [280], [931], [1060], [1218] |
|  | Welsh Government Office for Science | [1144] |
| **Egypt** | Egyptian National Research Centre | [1226] |
|  | Egyptian Academy of Scientific Research and Technology (ASRT) | [327], [1027] |
|  | Science and Technology Development Fund (STDF) | [74], [163] |
| **France** | Agence Nationale de la Recherche | [169], [643], [853] |
|  | Centre National de la Recherche Scientifique (F.A.R.) | [1238] |
|  | Fondation pour la Recherche Médicale (FRM - Fondation pour la Recherche Médicale) | [169] |
|  | French Institute for Public Health Surveillance | [219] |
|  | Hauts-de-France | [1158] |
|  | Institut de Recherche pour le Développement (IRD) | [853] |
|  | La fondation Infectiopôle Sud | [169] |
|  | LabEx Integrative Biology of Emerging Infectious Diseases (F.A.R.) | [242], [1015], [1143], [1241], [1238] |
|  | Montpellier University of Excellence (MUSE) | [853] |
|  | Sanofi Pasteur | [12], [147], [514] |
|  | SPILF Émergences | [1169] |
|  | University of Lille and CNRS | [10], [369] |
| **Australia** | Alliance project | [938] |
|  | Australian Government | [199] |
|  | Australian National Health & Medical Research Council (NHMRC) | [71], [136], [424], [1023], [1186], [1243] |
|  | CSIRO appropriation fund | [89], [453] |
|  | CSL | [147], [514] |
| **Taiwan** | Academia Sinica | [574] |
|  | Kaohsiung Medical University Hospital | [492] |
|  | Ministry of Education | [921], [1111] |
|  | Ministry of Science and Technology (MOST) | [79], [289], [307], [372], [403], [492], [703], [863], [921], [1117] |
|  | National Cheng Kung University Hospital | [492] |
|  | National Chung-Hsing University | [2] |
|  | National Science Council | [1099] |
|  | National Science Council of Taiwan | [2] |
|  | National Taiwan University | [403], [703], [863] |
|  | National Yang-Ming University | [1207] |
|  | Taipei Medical University | [492] |
|  | Taiwan Science Foundation | [244] |
| **India** | Council of Scientific and Industrial Research | [935] |
|  | Government of India | [73], [267], [897] |
|  | Indian Council of Medical Research | [465] |
|  | Indian Institute of Science Education and Research (IISERB) | [254], [1097], [1205] |
|  | Jawaharlal Nehru University | [1103] |
|  | Presidency University | [460] |
| **United Arab Emirates** | Abu Dhabi Department of Health | [1016] |
|  | Health Authority of Abu Dhabi | [1220] |
|  | Mohammed Bin Rashid University of Medicine and Health Sciences | [731] |
| **Iran** | Babol University of Medical Sciences | [138] |
|  | Baqiyatallah University of Medical Sciences | [355] |
|  | Shahid Beheshti University of Medical Sciences | [1193] |
|  | Tabriz University of Medical Sciences | [343] |
|  | Tehran University of Medical Sciences | [231] |
|  | Urmia University of Medical Sciences | [14], [1197] |
| **Spain** | Catalan Ministry of Health | [73] |
|  | Government of Spain | [506], [722], [745], [788], [821] |
|  | Ministry of Science and Innovation of Spain (MCINN) | [407] |
| **Canada** | An Ontario Veterinary College (OVC) Fellowship | [41] |
|  | Canadian Institutes of Health Research | [922], [926], [1000] |
|  | Canada Research Chairs Program (ALG) | [41] |
|  | Natural Science and Engineering Research Council of Canada | [41], [303], [494], [641], [1046] |
|  | Public Health Agency of Canada | [844], [985] |
|  | University of Saskatchewan | [394], [494], [641] |
| **Qatar** | Hamad Medical Corporation | [801], [861] |
|  | Ministry of Public Health | [543], [659], [881], [1131] |
|  | Qatar National Research Fund | [590] |
|  | Qatar University | [251], [859] |
| **Switzerland** | A. Vogel AG | [606] |
|  | Federal Ministry of Education and Research | [53] |
|  | Novartis | [147], [514] |
|  | Roche | [147], [514] |
|  | Swiss National Science Foundation | [428], [861], [1121] |
|  | Union Bank of Switzerland (UBS) Optimus Foundation | [806] |
| **Kenya** | Sino-Africa Joint Research Center (SAJOREC) | [504] |
| **Pakistan** | China-Pakistan Economic Corridor | [286] |
|  | External Cooperation Program of CAS | [286] |
|  | International Cooperation on Key Technologies of Biosafety | [286] |
|  | University of Engineering and Technology (UET) Lahore | [264] |
| **Singapore** | Institute of Molecular and Cell Biology | [894] |
|  | Ministry of Education | [461], [918] |
|  | Ministry of Health | [918], [1061] |
|  | Nanyang Technological University Singapore | [133] |
|  | National University of Singapore | [101], [894], [918] |
|  | Singapore National Research Foundation | [453], [712] |
| **Italy** | Italian Ministry of Health | [332] |
|  | Regione Umbria | [292] |
| **Jordan** | British Embassy in Amman | [1017] |
|  | Jordan University of Science and Technology | [527], [995] |
| **Turkey** | Turkish Academic Network and Information Center | [1120] |
| **Thailand** | Chulalongkorn University | [598] |
|  | Ministry of Public Health | [684] |
|  | National Science and Technology Development Agency (NSTDA) | [598] |
|  | Thailand Graduate Institute of Science and Technology | [854] |
|  | Thailand Research Fund | [854] |
| **Kuwait** | Kuwait University Research Administration | [905] |
|  | Kuwait Foundation for the Advancement of Sciences | [124] |
| **Austria** | Austrian Science Fund (FWF) | [637] |
|  | Mundipharma Research GmbH & Co. KG (MRG) | [604], [977] |
|  | University of Veterinary Medicine Vienn | [768] |
| **Belgium** | Ghent University | [378] |
| **Brazil** | Conselho Nacional de Desenvolvimento Científico e Tecnológico (CNPq) | [334], [1149] |
| **Indonesia** | Eijkman Institute for Molecular Biology | [339] |
|  | Ministry of Education, Culture, Research and Technology | [589] |
|  | Prof. Dr. Sulianti Saroso IDH | [339] |
| **Iraq** | Higher Committee for Education Development in Iraq (HCED) | [52] |
| **Israel** | Israel Science Foundation | [489] |
|  | Ministry of Health | [816] |
| **Kazakhstan** | Ministry of Education and Science of the Republic of Kazakhstan | [1062], 1129] |
| **South Africa** | The National Research Foundation (NRF) of South Africa | [59], [1030], [1168] |
| **Russia** | Russian Foundation for Basic Research (RFBR) | [1256] |
| **Poland** | Polish Ministry of Science and Higher Education | [544] |
| **Malaysia** | Institute for Medical Research Operational | [876] |
|  | Malaysian Ministry of Education | [350] |
|  | Universiti Sains Malaysia | [350] |
| **Ireland** | Science Foundation Ireland | [634] |
| **Finland** | Finnish Cultural Foundation | [776] |
|  | Jenny and Antti Wihuri Foundation | [776] |
|  | Medical Research Council of the Academy of Finland | [776] |
|  | Sigrid Juselius Stiftelse Sr | [776] |
| **Argentina** | Sistema Nacional de Computación de Alto Desempeño | [841] |
| **Colombia** | National University of Colombia | [1026] |
| **New Zealand** | New Zealand Ministry of Education | [406] |
| **Portugal** | Ministry of Education | [1240] |
| **Ethiopia** | Addis Ababa University | [509] |
| **Tunisia** | Tunisian Ministry of Higher Education and Research | [509] |
| **Estonia** | Institute of Technology | [1201] |

Mapping of MERS-CoV-related research themes, 1 January 2012 to 24 January 2023

| **Themes** | **Appendix S1: Technical Appendix Reference** |
| --- | --- |
| **Molecular genetics** | [1], [2], [9], [17], [20], [21], [22], [23], [24], [25], [35], [37], [44], [45], [49], [52], [53], [57], [59], [67], [69], [87], [99], [100], [108], [110], [115], [118], [120], [121], [126], [150], [151], [158], [165], [167], [169], [171], [180], [181], [183], [185], [186], [188], [191], [200], [202], [207], [209], [230], [232], [233], [235], [237], [239], [240], [241], [247], [248], [250], [261], [262], [264], [265], [266], [270], [271], [280], [281], [282], [283], [284], [285], [289], [292], [300], [301], [302], [303], [304], [305], [306], [307], [310], [311], [312], [314], [317], [324], [332], [333], [334], [335], [336], [349], [360], [361], [363], [364], [365], [366], [367], [368], [370], [373], [374], [384], [385], [397], [401], [402], [403], [413], [415], [425], [426], [427], [428], [438], [446], [449], [450], [451], [453], [454], [460], [461], [464], [465], [466], [469], [486], [487], [488], [489], [490], [491], [492], [493], [494], [495], [496], [497], [498], [499], [500], [501], [502], [503], [504], [507], [508], [513], [532], [534], [539], [540], [541], [549], [554], [555], [557], [558], [559], [561], [563], [565], [568], [570], [571], [572], [573], [574], [577], [578], [585], [592], [596], [608], [615], [616], [622], [626], [631], [632], [633], [634], [635], [637], [638], [644], [648], [657], [658], [659], [660], [662], [674], [678], [681], [686], [687], [688], [689], [692], [697], [698], [701], [702], [703], [708], [712], [716], [717], [718], [722], [723], [724], [725], [728], [735], [738], [739], [742], [743], [744], [745], [747], [753], [755], [756], [757], [759], [763], [765], [768], [776], [779], [780], [783], [785], [788], [783], [785], [788], [790], [791], [792], [803], [804], [807], [808], [809], [810], [811], [812], [813], [814], [818], [819], [821], [824], [825], [829], [834], [836], [837], [838], [840], [841], [842], [843], [844], [847], [848], [852], [853], [854], [855], [857], [858], [863], [864], [868], [873], [874], [876], [877], [879], [880], [886], [894], [895], [897], [902], [903], [905], [911], [915], [916], [917], [919], [920], [921], [922], [923], [924], [929], [930], [931], [933], [939], [953], [962], [963], [964], [972], [974], [983], [984], [987], [992], [993], [998], [1001], [1003], [1004], [1005], [1008], [1011], [1026], [1029], [1030], [1036], [1038], [1039], [1046], [1048], [1049], [1053], [1059], [1075], [1077], [1084], [1085, [1086], [1087], [1088], [1091], [1097], [1098], [1099], [1100], [1105], [1106], [1107], [1108], [1109], [1110], [1112], [1113], [1114], [1116], [1117], [1118], [1119], [1120], [1121], [1124], [1126], [1134], [1143], [1144], [1145], 1146], [1149], [1150], [1152], [1156], [1157], [1158], [1164], [1165], [1168], [1171], [1172], [1176], [1177], [1182], [1183], [1186], [1194], [1195], [1200], [1201], [1203], [1205], [1210], [1211], [1212], [1216], [1218], [1226], [1227], [1230], [1232], [1234, [1236], [1238], [1239], [1240], [1246], [1254], [1256], [1259], [1261], [1263] |
| **Therapeutics (drug-related or others)** | [1], [2], [3], [4], [5], [6], [7], [8], [9], [10], [17], [33], [34], [42], [44], [45], [53], [57], [66], [67], [70], [84], [88], [90], [105], [106], [107], [132], [133], [134], [141], [150], [158], [167], [169], [170], [173], [174], [175], [176], [177], [183], [188], [194], [198], [201], [207], [209], [236], [237], [253], [254], [261], [266], [267], [268], [269], [272], [281], [304], [310], [313], [316], [324], [325], [326], [327], [328], [330], [366], [367], [369], [372], [387], [389], [399], [403], [405], [406], [409], [446], [452], [470], [477], [478], [480], [481], [487], [492], [493], [495], [511], [532], [534], [540], [544], [545], [551], [552], [555], [559], [560], [562], [566], [567], [569], [573], [574], [577], [601], [603], [604], [605], [606], [613], [629], [631], [632], [633], [634], [635], [639], [640], [641], [642], [650], [686], [687], [700], [703], [704], [712], [721], [723], [724], [725], [742], [743], [748], [759], [783], [798], [799], [832], [874], [884], [895], [898], [901], [905], [906], [914], [924], [935], [952], [954], [955], [956], [957], [958], [959], [981], [982], [1000], [1013], [1014], [1027], 1031], 1032], [1033], [1034], [1035], [1041], [1043], [1054], [1072], [1076], [1090], [1095], [1100], [1101], [1102], [1104], [1111], [1113], [1115], [1116], [1117], [1119], [1121], [1124], [1135], [1136], [1142], 1143], [1144], [1145], [1146], [1150], [1151], [1153], [1171], [1175], [1181], [1205], [1207], 1208], 1211], [1212], [1222], [1223], [1224], [1225], [1227], [1233], [1234], [1236], [1238], [1240], [1255], [1261], [1264] |
| **Clinical features & complications** | [12], [14], [15], [18], [39], [46], [47], [61], [68], [81], [91], [92], [94], [96], [97], [98], [109], [145], [149], [157], [164], [170], [178], [184], [188], [196], [197], [200], [207], [208], [211], [212], [213], [214], [216], [218], [219], [221], [222], [223], [224], [225], [226], [227], [229], [231], [234], [235], [236], [242], [243], [244], [245], [249], [250], [257], [258], [262], [283], [284], [285], [290], [291], [308], [315], [316], [320], [323], [342], [349], [353], [360], [363], [369], [378], [381], [401], [402], [417], [418], [419], [421], [422], [424], [435], [454], [472], [476], [482], [483], [491], [496], [515], [521], [526], [531], [533], [535], [538], [542], [550], [554], [572], [578], [579], [580], [581], [582], [595], [597], [598], [607], [615], [616], [622], [625], [627], [655], [664], [665], [693], [694], [695], [697], [705], [714], [719], [722], [723], [726], [727], [728], [732], [734], [735], [738], [739], [745], [747], [752], [760], [763], [765], [769], [776], [779], [783], [785], [788], [789], [795], [801], [809], [812], [822], [832], [845], [846], [869], [891], [892], [904], [936], [937], [943], [944], [1025], [1028], [1037], [1057], [1068], [1069], [1077], [1078], [1112], [1122], [1126], [1147], [1155], [1164], [1165], [1168], [1172], [1176], [1177], [1182], [1183], [1203], [1209], [1211], [1212], [1220, [1223], [1231], [1236], [1242], [1252] |
| **Vaccinology & Immunization** | [9], [43], [48], [49], [55], [56], [58], [64], [67], [71], [72], [74], [75], [76], [77], [80], [83], [84], [86], [121], [129], [130], [137], [138], [163], [172], [189], [192], [195], [199], [201], [206], [255], [270], [288], [293], [305], [314], [318], [329], [351], [352], [371], [375], [384], [397], [406], [407], [408], [412], [425], [426], [436], [452], [489], [506], [508], [522], [548], [551], [553], [563], [564], [556], [567], [583], [584], [586], [587], [588], [591], [592], [596], [602], [609], [626], [638], [645], [646], [647], [649], [663], [690], [691], [693], [694], [695], [696], [715], [723], [724], [726], [736], [749], [750], [753], [758], [817], [821], [856], [861], [862], [872], [882], [885], [899], [900], [913], [960], [961], [971], [987], [994], [995], [996], [997], [1044], [1045], [1049], [1054], [1071], [1073], [1092], [1102], [1103], [1125], [1133], [1140], [1141], [1150], [1152], [1187], [1195], [1196], [1202], [1209], [1213], [1215], [1238] |
| **Seroprevalence** | [26], [89], [95], [116, [127], [128], [129, [155], [193], [232], [233], [234], [243], [251], [259], [277], [279], [280], [286], [294], [295], [296], [297], [298], [299], [319], [322], [341], [374], [379], [415], [420], [423, [435], [443], [445], [448], [483], [484], [499], [504], [509], [516], [518], [525], [526], [527], [529], [538], [546], [547], [607], [624], [629], [630], [651], [653], [666], [675], [677], [679], [680], [682], [683], [684], [693, [694], [695], [696], [699], [713], [715], [731], [733], [754], [760], [764], [766], [767], [768], [771], [775], [781], [782], [791], [792], [796], [797], [800], [802], [805], [806], [815], [816], [830], [842], [851], [857], [867], [878], [912], [928], [941], [942], [946], [947], [1015], [1016], [1017], [1018], [1021], [1042, [1050], [1051], [1052], [1055, [1056], [1057], [1058], [1059], [1060], [1061], [1062], [1063], [1064], [1065], [1066], [1079], [1127], [1128], [1129], [1139], [1189], [1190], [1191], [1192], [1217], [1219] |
| **Transmissibility** | [13], [19], [31], [40], [50], [60], [65], [73], [78], [82], [103], [104], [119], [122], [125], [146], [172], [191], [211], [239], [258], [388], [391], [394], [398], [413], [414], [415] , [429], [432], [451], [455], [462], [463], [467], [485], [514], [516], [518], [530], [536], [575], [599], [623], [643, [655], [681], [683], [707], [720], [729], [731], [746], [766], [777], [787], [810], [818], [823], [824], [825], [830], [831], [834], [837], [840], [850], [859], [865], [866], [870], [871], [876], [887], [889], [917], [918], [926], [928], [934], [991], [1002], [1005], [1006], [1008], [1016], [1017], [1019], [1020], [1021], [1022], [1023, [1024], [1137], [1154], [1155], [1186], [1199], [1204], [1220], [1237], [1241], [1247], [1250], [1251] |
| **Outbreak investigation** | [13], [32], [38], [39], [51], [68], [69], [93], [95], [116], [156], [157], [184], [197], [213], [214], [216], [218], [219], [220], [221], [222], [223], [225], [231], [233], [239], [273], [274], [275], [276], [315], [322], [339], [411], [416], [419], [421], [422], [423], [443], [468], [483], [510], [515], [518], [524], [528], [529], [535], [536], [538], [543], [549], 597], [598], [666], [676], [683], [720], [730], [746], [762], [770], [778], [793], [820], [839], [850], [851], [887], [888], [889], [890], [893], [896], [904], [951], [1009], [1010], [1024], [1040], [1056], [1155], [1214], [1217], [1219], [1221], [1242], [1252] |
| **Laboratory diagnostics** | [11], [54], [59], [63], [77], [79], [108], [114], [117], [123], [131], [139], [141], [142], [189], [203], [204], [205], [228], [238], [246], [251], [259], [260], [271], [278], [331], [336], [337], [344], [345], [346], [347], [348], [350], [354], [355], [356], [357], [358], [437], [441], [442], [443], [445], [447], [479], [494], [512], [518], [528], [600], [610], [614], [636], [673], [674], [833], [849], [854], [875], [883], [909], [910], [974], [978], [979], [980], [984], [985], [986], [989], [990], [999], [1012], [1028], [1047], [1051], [1054], [1070], [1089], [1093], [1123], [1130], [1206], [1232], [1235], [1244], [1248], [1249] |
| **Anthropology and Social behavior** | [28], [29], [62], [101], [111], [113], [140], [147], [152], [160], [161], [162], [166], [168], [256], [263], [343], [362], [390], [396], [439], [440], [444], [457], [458], [474], [475], [537], [543], [628], [656], [661], [667], [668], [669], [670], [671], [672], [685], [706], [709], [710], [740], [761], [786], [826], [827], [828], [831], [907], [908], [948], [949], [966], [968], [970], [975], [976], [1160], [1161], [1162], [1163], [1167], [1174], [1185], [1193], [1198], [1228], [1229], [1257], [1260], [1262] |
| **Spillover** | [41], [99], [239], [332], [333], [334], [340], [368], [370], [374], [376], [382], [397], [420], [449], [466], [484], [485], [488], [490], [504], [509], [517], [525], [526], [528], [541], [549], [659], [660], [680], [682], [689], [699], [717], [718], [751], [755], [756], [771], [774], [780], [790], [791], [792], [814], [991], [992], [1002], [1019], [1020], [1021], [1079], [1080], [1086], [1114], [1178], [1180], [1204], [1226], [1241], [1254], [1263] |
| **Outbreak preparedness and response** | [16], [30], [31], [73], [102], [103], [124], [135], [136], [144], [159], [182], [190], [210], [278], [353], [377], [380], [381], [386], [388], [391], [392], [396], [400], [429], [430], [431], [432], [434], [436], [463], [469], [479], [517], [543], [598], [643], [654], [673], [676], [707], [729], [741], [890], [896], [908], [926], [940], [945], [969], [1010], [1083], [1127], [1128], [1130], [1148], [1159], [1169], [1243] |
| **Infection prevention and Control (IPC)** | [18], [60], [62], [111], [113], [152], [159], [160], [161], [191], [210], [362], [393], [398], [414], [456], [520], [604], [610], [611], [612], [619], [620], [621], [671], [672], [684], [706], [736], [777], [820], [870], [871], [889], [890], [893], [938], [948], [975], [977], [1018], [1064], [1091], [1126], [1132], [1179], [1200], [1247], [1251], [1253] |
| **Impact on health system\BOD & economic impact** | [14], [15], [94], [96], [179], [196], [214], [223], [226], [242], [290], [291], [309], [320], [322], [342], [383], [395], [422], [424], [430], [431], [433], [435], [472], [510], [524], [536], [538], [579], [595], [627], [752], [772], [778], [793], [801], [1007], [1067], [1082], [1094], [1170], [1173], [1184], [1197], [1258] |
| **Mathematical models for MERS** | [12], [16], [19], [27], [31], [36], [40], [50], [65], [73], [78], [135], [136], [144], [146], [382], [394], [429], [430], [431], [432], [434], [436], [455], [461], [463], [485], [530], [599], [654], [835], [932], [950], [973], [988], [1083], [1137], [1154], [1204], [1243] |
| **Comorbidities** | [94], [96], [196], [217], [234], [240], [242], [243], [245], [252], [290], [291], [342], [353], [378], [388], [524], [542], [576], [579], [593], [617], [618], [644], [705], [719], [727], [732], [734], [752], [760], [772], [789], [795], [1067], [1081], [1197], [1223], [1252], [1258] |
| **Surveillance systems** | [30], [93], [273], [319], [322], [380], [392], [411], [448], [468], [514], [523], [859], [823], [871], [918], [945], [1042], [1074], [1127], [1128], [1130] |
| **MERS-CoV & SARS-CoV-2 cross-reactive immunity** | [105], [112], [187], [277], [280], [292], [294], [335], [506], [589], [590], [853], [856], [925], [1053], [1066], [1067], [1092], [1097], [1245] |
| **Silent/asymptomatic** | [51], [145], [154], [155], [410], [418], [424], [526], [726], [734], [823], [1057], [1080], [1204], [1231] |
| **One Health approach** | [41], [286], [397], [484], [504], [517], [881], [1131], [374] |
| **MERS & mental health** | [143], [148], [153], [168], [321], [359], [457], [459], [471], [473], [519], [618], [661], [709], [710], [711], [773], [786], [828], [907], [908], [965], [967], [1138], [1167], [1260] |
| **Case management** | [94], [182], [224], [291], [322], [392], [476], [477], [478], [665], [666], [822], [832], [845], [869], [891], [892], [937], [1025], [1122], [1155], [1179] |
| **MERS & COVID-19** | [215], [217], [248], [249], [252], [254], [256], [264], [287], [440], [732], [1188] |
| **Traditional medicine** | [652], [682], [775], [860], [927], [1147] |
| **MERS & pregnancy** | [404], [594], [737], [784], [794], [1096] |
| **Trade and animal food production** | [338], [774], [789], [1080], [1221] |
| **Reinfection** | [410], [528], [529], [623], [696] |

Mapping of MERS-CoV-related research themes across years, 1 January 2012 to 24 January 2023 (**Appendix S1: Technical Appendix Reference)**

|  | **2012** | **2013** | **2014** | **2015** | **2016** | **2017** | **2018** | **2019** | **2020** | **2021** | **2022** | **2023** |
| --- | --- | --- | --- | --- | --- | --- | --- | --- | --- | --- | --- | --- |
| Molecular genetics | Non | [20], [305], [563], [631], [765], [779], [790], [1011], [1029], [1113], [1177], [1195], [1218] | [44], [49], [100], [110], [165], [171], [185], [186], [202], [239], [303], [304], [311], [446], [539], [541], [549], [571], [577], [622], [659], [718], [747], [768], [792], [814], [847], [902], [953], [963], [983], [993],  [1003], [1004], [1008], [1009], [1030], [1088], [1099], [1116], [1150], [1183], [1226], [1259] | [69], [200], [271], [289], [307], [324], [426], [496], [513], [608], [626], [632], [635], [648], [674], [687], [712], [808], [812], [834], [852], [876], [879], [886], [929], [933], [1146], [1171] | [2], [35], [37], [67], [120], [151], [158], [183], [188], [233], [317], [361], [363], [365], [449], [453], [466], [469], [494], [573], [574], [657], [689], [703], [708], [757], [759], [776], [783], [785], [803], [804], [807], [810], [863], [868], [920], [962], [972], [1001], [1087], [1194], [1246] | [9], [22], [169], [247], [250], [301], [302], [332], [374], [427], [464], [490], [501], [565], [572], [585], [692], [716], [723], [753], [763], [838], [848], [854], [911], [922], [1085], [1109], [1110], [1112], [1114], [1134], [1203], [1210], [1216], [1230], [1261] | [21], [57], [59], [99], [121], [209], [281], [282], [366], [368], [454], [487], [504], [555], [570], [596], [633], [686], [701], [717], [722], [724], [738], [780], [818], [858], [877], [894], [903], [905], [921], [984], [992], [998], [1003], [1005], [1098], [1108], [1119], [1156], [1164], [1172], [1211], [1234], [1263] | [52], [87], [126], [207], [235], [237], [240], [262], [336], [373], [384], [397], [402], [415], [425], [450], [493], [495], [497], [498], [554], [559], [561], [578], [616], [658], [681], [791], [824], [825], [857], [917], [923], [939], [1049], [1059], [1075], [1084], [1086], [1106], [1107], [1121], [1158], [1212], [1238] | [23], [85], [115], [118], [180], [181], [248], [261], [270], [306], [310], [314], [349], [370], [451], [486], [491], [502], [508], [568], [615], [702], [739], [809], [813], [829], [836], [840], [853], [855], [919], [1046], [1053], [1077], [1091], [1100], [1117], [1118], [1165], [1186], [1201], [1227], [1240] | [1], [25], [45], [53], [108], [232], [264], [266], [280], [285], [312], [335], [364], [367], [385], [401], [403], [460], [460], [461], [465], [488], [492], [503], [505], [558], [592], [637], [638], [660], [662], [678], [688], [697], [725], [735], [755], [756], [788], [819], [821], [841], [864], [873], [880], [895], [915], [916], [931], [974], [987], [1037], [1039], [1048], [1105], [1120], [1126], [1143], [1144], [1149], [1152], [1157], [1176], [1200], [1205], [1236], [1239], [1256] | [17], [24], [150], [167], [191], [230], [241], [283], [284], [292], [300], [333], [334], [360], [413], [428], [438], [489], [500], [507], [532], [534], [540], [557], [634], [644], [698], [728], [742], [743], [744], [745], [811], [837], [842], [843], [844], [897], [924], [930], [964], [1026], [1036], [1038], [1097], [1124], [1145], [1168], [1182], [1232], [1254] | [265], [499], [874] |
| Therapeutics (drug-related or others) | Non | [84], [141], [177], [316], [631], [721], [799], [1113], [1135], [1224] | [44], [304], [369], [446], [452], [567], [577], [642], [832], [1013], [1043], [1116], [1150] | [134], [236], [324], [326], [632], [635], [687], [712], [958], [959], [1102], [1146], [1171], [1207], [1225] | [2], [33], [67], [88], [158], [183], [188], [477], [478], [511], [569], [573], [574], [703], [759], [783], [798], [901], [957], [982], [1072], [1223] | [5], [9], [169], [194], [268], [389], [399], [552], [562], [629], [723], [1261] | [57], [66], [170], [176], [209], [281], [313], [366], [372], [387], [470], [487], [555], [603], [604], [633], [686], [724], [748], [905], [952], [954], [1034], [1035], [1119], [1211], [1234] | [3], [90], [107], [175], [198], [207], [237], [272], [406], [409], [493], [495], [559], [639], [641], [704], [906], [956], [1014], [1033], [1090], [1121], [1212], [1238] | [34], [42], [70], [132], [201], [253], [261], [310], [328], [405], [481], [545], [560], [606], [640], [955], [1000], [1031], [1054], [1076], [1100], [1111], [1117], [1153], [1175], [1208], [1222], [1227], [1240], [1255], [1264] | [1], [6], [8], [45], [53], [105], [133], [174], [254], [266], [267], [269], [325], [330], [367], [403], [492], [544], [605], [700], [725], [884], [895], [914], [935], [1032], [1095], [1143], [1144], [1205], [1236] | [4], [7], [10], [17], [106], [150], [167], [173], [327], [480], [532], [534], [540], [551], [566], [601], [613], [634], [650], [742], [743], [898], [924], [981], [1027], [1041], [1101], [1104], [1115], [1124], [1142], [1145], [1151], [1181], [1233] | [874], [1136] |
| Clinical features & complications | Non | [15], [219], [316], [476], [535], [655], [765], [779], [1177] | [68], [92], [213], [214], [218], [369], [435], [538], [622], [664], [747], [769], [822], [832], [1183], [1252] | [96], [149], [200], [221], [231], [236], [308], [323], [419], [483], [496], [515], [531], [579], [597], [719], [812], [846], [1068] | [12], [39], [47], [61], [94], [109], [178], [188], [197], [216], [223], [227], [229], [243], [244], [258], [363], [714], [776], [783], [785], [904], [936], [937], [943], [1025], [1069], [1122], [1209], [1220], [1223] | [157], [184], [211], [245], [250], [417], [482], [521], [550], [572], [598], [723], [726], [763], [789], [1057], [1112], [1155], [1203] | [14], [81], [98], [170], [222], [224], [290], [315], [353], [378], [381], [454], [472], [526], [533], [542], [625], [722], [738], [752], [845], [944], [1164], [1172], [1211], [1242] | [97], [164], [196], [207], [212], [225], [226], [235], [242], [262], [402], [422], [554], [578], [582], [595], [616], [705], [795], [869], [1212] | [91], [145], [234], [249], [320], [349], [424], [491], [580], [581], [615], [734], [739], [809], [892], [1077], [1078], [1165] | [18], [208], [285], [291], [342], [401], [421], [607], [627], [665], [693], [697], [727], [732], [735], [760], [788], [801], [891], [1028], [1037], [1126], [1147], [1176], [1231], [1236] | [46], [257], [283], [284], [360], [418], [694], [695], [728], [745], [1168], [1182] | Non |
| Vaccinology & Immunization | Non | [77], [84], [305], [407], [563], [817], [1195] | [49], [452], [567], [586], [647], [715], [971], [1044], [1141], [1150] | [55], [58], [83], [426], [626], [663], [885], [960], [996], [1102], [1140] | [67], [72], [75], [129], [553], [564], [583], [649], [900], [997], [1209] | [9], [192], [375], [587], [696], [723], [726], [749], [753], [872], [882], [961], [994], [1196] | [76], [121], [189], [195], [371], [408], [412], [522], [596], [691], [724], [1103] | [56], [64], [74], [130], [163], [172], [288], [351], [384], [397], [406], [425], [436], [556], [995], [1045], [1049], [1071], [1125], [1213], [1238] | [48], [80], [86], [201], [206], [270], [293], [314], [508], [750], [758], [899], [1054], [1073], [1187] | [329], [506], [548], [592], [602], [638], [645], [693], [821], [861], [862], [987], [1092], [1133], [1152], [1215] | [43], [71], [137], [138], [199], [255], [318], [352], [489], [551], [584], [588], [591], [609], [646], [690], [694], [695], [856], [913], [1202] | [736] |
| Seroprevalence | Non | [679], [805], [1051] | [127], [128], [435], [509], [516], [538], [546], [651], [677], [713], [715], [768], [792], [796], [806], [1042], [1052], [1219] | [26], [89], [95], [116], [443], [483], [666], [675], [680], [683], [733], [767], [800], [878], [941], [942], [946], [1060], [1079] | [129], [233], [243], [322], [420], [423], [547], [684], [912], [1018], [1129] | [279], [298], [299], [374], [379], [527], [624], [629], [696], [867], [1015], [1055], [1057], [1058], [1127], [1139] | [155], [286], [296], [448], [504], [518], [526], [630], [682], [782], [816], [851], [1050], [1065], [1190], [1191] | [251], [259], [415], [445], [484], [754], [766], [771], [791], [797], [802], [815], [857], [928], [1016], [1056], [1059], [1064], [1217] | [193], [234], [295], [319], [341], [525], [529], [764], [775], [781], [1021], [1063], [1128] | [232], [280], [294], [607], [653], [693], [699], [760], [830], [947], [1017], [1066], [1189] | [277], [297], [694], [695], [731], [842], [1061], [1062], [1192] | [499] |
| Transmissibility | Non | [643], [655], [926], [934] | [78], [146], [239], [514], [516], [777], [823], [991], [1002], [1008], [1137] | [19], [683], [720], [729], [834], [876], [918], [1199], [1251] | [13], [50], [258], [398], [414], [463], [467], [575], [623], [746], [810], [831], [850], [887], [1023], [1154], [1220], [1241], [1250] | [65], [211], [429], [432], [536], [787], [865], [870], [889], [1020], [1022], [1155], [1247] | [31], [60], [82], [103], [104], [518], [530], [707], [818], [859], [866], [1005], [1006], [1237] | [40], [172], [415], [599], [681], [766], [824], [825], [917], [928], [1016], [1019], [1024] | [73], [122], [388], [451], [455], [840], [871], [1021], [1186] | [391], [394], [462], [830], [1017] | [119], [125], [191], [413], [485], [731], [837], [1204] | Non |
| Outbreak investigation | Non | [51], [219], [220], [535] | [38], [68], [213], [214], [218], [239], [411], [538], [549], [676], [770], [896], [1219], [1221], [1252] | [32], [69], [95], [116], [221], [231], [274], [416], [419], [443], [483], [515], [528], [597], [666], [683], [720], [778], [839], [951] | [13], [39], [93], [197], [216], [223], [233], [273], [275], [322], [423], [468], [730], [746], [820], [850], [887], [890], [904], [1010], [1214] | [157], [184], [276], [524], [536], [543], [598], [762], [888], [889], [893], [1155] | [156], [222], [315], [510], [518], [851], [1242] | [225], [339], [422], [793], [1024], [1040], [1056], [1217] | [529] | [421] | [1009] | Non |
| Laboratory diagnostics | Non | [77], [141], [1012], [1051] | [337], [673], [909], [989], [999] | [79], [117], [260], [271], [331], [344], [345], [443], [479], [512], [528], [610], [674], [990] | [123], [203], [246], [355], [358], [494], [985], [1093], [1130], [1244], [1249] | [63], [142], [228], [437], [854], [883], [910], [1070], [1123], [1206] | [11], [59], [139], [189], [205], [347], [356], [447], [518], [614], [636], [849], [979], [984] | [114], [251], [259], [336], [346], [350], [357], [445], [833], [1047], [1248] | [131], [204], [278], [354], [441], [442], [978], [1054], [1235] | [54], [108], [238], [348], [600], [875], [974], [986], [1028], [1089] | [277], [1232] | Non |
| Anthropology and Social behavior | Non | Non | Non | [113], [162], [439], [672] | [147], [475], [656], [667], [671], [826], [831], [949], [966], [1162], [1198] | [28], [111], [140], [396], [457], [543], [865], [706], [709], [827], [907], [970], [975], [976], [1174], [1228], [1260], [1262] | [62], [101], [362], [444], [458], [474], [661], [668], [710], [740], [1161], [1167], [1229] | [152], [390], [669], [670], [786], [908], [948], [1160], [1185], [1257] | [29], [160], [161], [168], [256], [263], [343], [628], [828], [968], [1163] | [166], [440], [761], [1193] | [537] | Non |
| Spillover | Non | [790] | [239], [340], [509], [541], [549], [659], [718], [774], [792], [814], [991], [1002], [1226] | [528], [680], [1079] | [420], [449], [466], [689], [1241] | [332], [374], [376], [490], [1020], [1114] | [99], [368], [504], [526], [682], [717], [751], [780], [992], [1263] | [41], [397], [484], [517], [771], [791], [1086], [1019] | [370], [525], [1021], [1080], [1180] | [488], [660], [696], [755], [756], [1178] | [333], [334], [382], [485], [1204], [1254] | Non |
| Outbreak preparedness and response | Non | [377], [643], [926] | [673], [676], [741], [896], [945] | [30], [144], [190], [392], [400], [430], [479], [729] | [16], [159], [431], [434], [463], [469], [890], [1010], [1130] | [102], [135], [396], [429], [432], [543], [598], [969], [1127] | [31], [103], [353], [380], [381], [654], [707], [940], [1169] | [436], [517], [908], [1083] | [73], [136], [182], [210], [278], [386], [388], [1128], [1243] | [124], [391], [1148], [1159] | Non | Non |
| Infection prevention and Control (IPC) | Non | Non | [520], [777] | [113], [610], [672], [977], [1251] | [159], [398], [414], [612], [619], [671], [684], [820], [890], [1018] | [111], [706], [870], [889], [893], [975], [1247], [1253] | [60], [62], [362], [604], [611], [1132] | [152], [393], [620], [948], [1064] | [160], [161], [210], [621], [871], [1091], [1179] | [18], [938], [1126], [1200] | [191], [456] | [736] |
| Impact on health system\BOD & economic impact | Non | [15] | [214], [435], [538], [1184] | [430], [579], [778], [96] | [94], [223], [322], [431], [1007], [1173] | [395], [524], [536], [772] | [14], [290], [472], [510], [752] | [196], [226], [242], [309], [383], [422], [433], [595], [793], [1094], [1170] | [179], [320], [424], [1082], [1197], [1258] | [291], [342], [627], [801], [1067] | Non | Non |
| Mathematical models for MERS | Non | Non | [78], [146], [950], [1137] | [19], [144], [430], [973], [988] | [12], [16], [50], [431], [434], [463], [932], [1154] | [65], [135], [429], [432] | [31], [530], [654], [835] | [40], [436], [599], [1083] | [36], [73], [136], [455], [1243] | [394], [461] | [27], [382], [485], [1204] | Non |
| MERS & mental health | Non | Non | Non | Non | [473], [519], [711], [773], [1138] | [457], [709], [907], [1260] | [661], [710], [965], [967], [1167] | [321], [618], [786], [908] | [143], [168], [459], [471], [828] | [148], [359] | [153] | Non |
| Surveillance systems | Non | Non | [411], [514], [823], [945], [1042] | [30], [392], [918] | [93], [273], [322], [468], [523], [1130] | [1127] | [380], [448], [859] | Non | [319], [871], [1074], [1128] | Non | Non | Non |
| Case management | Non | [476] | [822], [832] | [392], [666] | [94], [322], [477], [478], [937], [1025], [1122] | [1155] | [224], [845] | [869] | [182], [892], [1179] | [291], [665], [891] | Non | Non |
| MERS-CoV & SARS-CoV-2 cross-reactive immunity | Non | Non | Non | Non | Non | Non | Non | Non | [112], [853], [1053], [1245] | [105], [280], [294], [335], [506], [925], [1066], [1067], [1092] | [187], [277], [292], [589], [590], [856], [1097] | Non |
| Silent/asymptomatic | Non | [51] | [823] | [154] | Non | [410], [726], [1057] | [155], [526] | Non | [145], [424], [1734], [1080] | [1231] | [418], [1204] | Non |
| Comorbidities | Non | Non | [1252] | [96], [579], [719] | [94], [243], [1223] | [245], [524], [593], [772], [789], [1081] | [290], [353], [378], [542], [752], | [196], [240], [242], [617], [618], [705], [795] | [234], [388], [576], [734], [1197], [1258] | [217], [252], [291], [342], [727], [732], [760], [1067] | [644] | Non |
| MERS & COVID-19 | Non | Non | Non | Non | Non | Non | Non | Non | [215], [248], [249], [256], [1188] | [217], [252], [254], [264], [287], [440], [732] | Non | Non |
| One Health approach | Non | Non | Non | Non | Non | [574] | [286], [504] | [41],  [397], [484], [517], [881], [1131] | Non | Non | Non | Non |
| Traditional medicine | Non | Non | Non | Non | Non | Non | [682] | Non | [775], [860], [927] | [652], [1147] | Non | Non |
| MERS & pregnancy | Non | Non | [1096] | Non | [404], [594], [784], [794] | [737] | Non | Non | Non | Non | Non | Non |
| Trade and animal food production | Non | Non | [774], [1221] | Non | Non | [789] | Non | Non | [1080] | Non | [338] | Non |
| Reinfection | Non | Non | Non | [528] | [623] | [410], [696] | Non | Non | [529] | Non | Non | Non |

Mapping of MERS-CoV-related research themes across geographic distribution, 1 January 2012 to 24 January 2023 (**Appendix S1: Technical Appendix Reference)**

|  | **Non-EMR** | | | **EMR** | | |
| --- | --- | --- | --- | --- | --- | --- |
|  | **United States of America** | **China** | **Republic of Korea** | **Saudi Arabia** | **UAE** | **Egypt** |
| Molecular genetics | [23], [24], [37], [44], [49], [87], [99], [126], [158], [171], [180], [185], [186], [188], [200], [240], [248], [281], [285], [305], [311], [314], [349], [363], [364], [370], [397], [446], [490], [496], [513], [539], [541], [559], [563], [565], [578], [585], [596], [616], [622**]**, [633], [635], [644], [648], [658], [662], [687], [688], [701], [702], [708], [723], [728], [747], [765], [783], [803], [807], [808], [814], [819], [834], [847], [848], [858, [873], [902], [903], [911], [916], [929], [962], [983], [993], [1001], [1011], [1036], [1037], [1053], [1077], [1085**]**, [1100], [1107], [1109], [1112], [1118], [1119], [1121], [1126], [1143], [1146], [1156], [1157], [1164], [1172], [1177], [1183], [1203], [1227], [1238], [1239], [1259], [1261], [1263] | [9], [20], [22], [35], [45], [57], [67], [110], [121], [165], [183], [207], [230], [232], [237], [250], [261], [262], [283], [284], [300], [301], [302], [306], [366], [368], [373], [425], [426], [427], [449], [499], [503], [532], [534], [540], [558], [561], [568], [570], [571], [573], [615], [657], [660], [717], [718], [735], [742], [743], [753], [755], [780], [785], [804], [812], [813], [836], [842], [843], [852], [886], [895], [915], [920], [923], [930], [933], [953], [963], [972], [974], [992], [1003], [1029], [1038], [1059], [1098], [1106], [1108], [1113], [1114], [1116], [1124], [1150], [1176], [1182], [1226], [1230], [1234] | [54], [115], [120], [150], [270], [335], [384], [450], [469], [491], [493], [495], [497], [498], [500], [507], [554], [555], [725], [757], [824], [825], [829], [984], [998], [1005], [1049], [1087], [1110], [1145], [1194], [1246] | [17], [21], [108], [118], [151], [233], [239], [247], [265], [360], [367], [401], [402], [415], [451], [486], [502], [505], [508], [549], [592], [678], [698], [738], [739], [763], [790], [763], [790], [810], [838], [840], [880], [917], [1008], [1039], [1048], [1134] | [69], [167], [374], [874] | [1091], [1232] |
| Therapeutics (drug-related or others) | [3], [6], [7], [33], [34], [44], [84], [88], [106], [134], [141], [158], [175], [188], [253], [272], [281], [326], [399], [446], [452], [481], [551], [559], [567], [633], [635], [642], [687], [723], [724], [748], [783], [798], [884], [901], [906], [954], [955], [956], [957], [958], [959], [1033], [1034], [1100], [1119], [1121], [1143], [1146], [1181], [1208], [1224], [1227], [1238], [1261] | [4], [9], [45], [57], [66], [67], [170], [177], [183], [207], [237], [261], [313], [316], [366], [387], [406], [409], [511], [532], [534], [540], [545], [552], [562], [573], [742], [743], [895], [1072], [1090], [1101], [1102], [1104], [1113], [1115], [1116], [1124], [1142], [1150], [1151], [1225], [1233], [1234], [1255] | [8], [42], [54], [90], [150], [174], [194], [198], [201], [236], [325], [493], [495], [555], [560], [700], [725], [952], [981], [1035], [1135], [1145], [1175] | [17], [70], [105], [107], [132], [327], [330], [367], [470], [477], [478], [566], [603], [605], [629], [640], [704], [832], [914], [1013], [1014], [1041], [1076], [1222], [1223] | [167], [874] | [173], [328], [613], [650], [1027], [1095], [1136] |
| Pathogenesis | [61], [81], [98], [109], [149], [164], [188], [200], [213], [229], [285], [349], [363], [496], [578], [616], [622], [723], [728], [747], [765], [783], [846], [1025], [1037], [1077], [1112], [1126], [1164], [1172], [1177], [1183], [1203], [1220] | [91], [92], [170], [207], [221], [244], [250], [262], [283], [284], [316], [421], [550], [581], [597], [615], [714], [735], [785], [812], [1069], [1147], [1176], [1182] | [12], [18], [39], [157], [211], [212], [222], [223], [227], [236], [243], [419], [491], [542], [554], [582], [595], [694], [695], [726], [936], [1057], [1122], [1155] | [15], [46], [47], [94], [96], [145], [178], [184], [196], [197], [214], [218], [225], [226], [234], [290], [291], [308], [320], [323], [342], [353], [360], [381], [401], [402], [418], [422], [435], [472], [476], [521], [526], [533], [535],[579], [580], [607], [625], [627], [665], [693], [719], [732], [738], [739], [752], [760], [763], [769], [789], [795], [822], [832], [845], [869], [891], [892], [904], [937], [943], [944], [1068], [1078], [1223], [1242] | [208], [482], [1028] | [216], [734] |
| Vaccinology & Immunization | [49], [80], [83], [84], [305], [314], [397], [408], [436], [452], [551], [553], [563], [564], [567], [583], [586], [596], [645], [649], [723], [724], [749], [758], [856], [862], [882], [971], [1044], [1073], [1092], [1187], [1238] | [9], [58], [64], [67], [75], [76], [77], [86], [121], [206], [375], [406], [412], [425], [426], [609], [647], [663], [753], [872], [885], [900], [961], [994], [996], [997], [1071], [1102], [1140], [1141], [1150], [1196] | [43], [137], [195], [201], [255], [270], [288], [293], [352], [384], [522], [548], [584], [646], [694], [695], [726], [750], [1045], [1049], [1125], [1133], [1202] | [56], [129], [318], [508], [556], [587], [588], [592], [690], [693], [696], [736], [1215] | Non | [74], [163] |
| Seroprevalence | [277], [294], [379], [443], [516], [527], [675], [683], [767], [782], [815], [830], [851], [912], [1060], [1190] | [26], [232], [286], [499], [682], [781], [842], [1051], [1059] | [155], [243], [666], [694], [695], [1057], [1064], [1129] | [129], [193], [233], [234], [279], [295], [296], [319], [322], [341], [415], [423], [435], [518], [526], [529], [607], [629], [651], [680], [693], [696], [713], [733], [754], [760], [796], [802], [1018], [1042], [1052], [1058], [1063], [1065], [1066], [1079], [1127], [1128], [1189], [1191] | [95], [374], [420], [946], [1016], [1056] | [298], [299], [766], [797], [1139] |
| Transmissibility | [40], [122], [398], [455], [463], [516], [623], [683], [729], [830], [834], [870], [887], [991], [1002], [1137], [1220] | [19], [65], [82], [865], [1204], [1247] | [13], [50], [60], [103], [104], [119], [211], [388], [391], [414], [432], [467], [530], [536], [720], [746], [824], [825], [831], [1005], [1022], [1024], [1154], [1155], [1251] | [125], [239], [415], [451], [462], [518], [777], [810], [840], [850], [866], [871], [889], [917], [1008], [1019], [1237] | [1016] | [485], [766] |
| Outbreak investigation | [32], [213], [276], [443], [683], [820], [851], [887], [1214] | [221], [273], [421], [468], [597], [951] | [13], [39], [157], [222], [223], [275], [416], [419], [524], [536], [666], [720], [746], [890], [1024], [1155] | [51], [184], [197], [214], [218], [225], [233], [239], [322], [422], [423], [510], [518], [529], [535], [549], [730], [778], [793], [839], [850], [888], [889], [893], [904], [1040], [1242] | [69], [95], [1010], [1056] | [216] |
| Laboratory diagnostics | [141], [331], [441], [443], [512], [610], [614], [636], [849], [989], [990], [1093], [1206], [1235] | [11], [63], [77], [79], [131], [203], [344], [348], [354], [875], [974], [979], [980], [1051] | [123], [139], [204], [246], [260], [345], [347], [357], [437], [883], [910], [984], [1070], [1089], [1123], [1130], [1249] | [108], [114], [142], [228], [346], [442], [447], [518], [600], [986] | [1028] | [1012], [1232] |
| Anthropology and Social behavior | [28], [166], [439], [1160], [1228], [1229] | Non | [29], [152], [160], [168], [256], [263], [362], [396], [457], [458], [474], [628], [685], [710], [826], [827], [831], [948], [949], [1161], [1162], [1163], [1167], [1174], [1257], [1260] | [62], [111], [113], [140], [161], [162], [444], [537], [656], [661], [667], [668], [669], [706], [709], [740], [761], [786], [828], [907], [908], [966], [968], [975], [976], [1185], [1262] | Non | Non |
| Spillover | [99], [370], [376], [397], [490], [541], [751], [814], [991], [1002], [1263] | [368], [382], [449], [660], [682], [717], [718], [755], [780], [992], [1114], [1204], [1226] | Non | [239], [340], [517], [526], [549], [680], [790], [1019], [1079], [1080], [1178], [1180] | [374], [420] | [485] |
| Outbreak preparedness and response | [16], [102], [182], [190], [380], [431], [434], [436], [463], [729], [969], [1159] | [400] | [30], [103], [159], [386], [388], [391], [396], [432], [469], [654], [890], [1130] | [210], [353], [381], [517], [908], [945], [1083], [1127], [1128] | [1010] | Non |
| Infection prevention and Control (IPC) | [398], [456], [610], [612], [820], [870], [1126] | [1247] | [18], [60], [152], [159], [160], [362], [393], [414], [621], [890], [948], [1064], [1251] | [62], [111], [113], [161], [210], [619], [620], [706], [736], [777], [871], [889], [893], [975], [1018], [1179] | Non | [938], [1091] |
| Impact on health system\BOD & economic impact | [383], [431] | Non | [223], [395], [524], [536], [595], [772], [1173] | [15], [94], [96], [179], [196], [214], [226], [290], [291], [320], [322], [342], [422], [433], [435], [472], [510], [579], [627], [752], [778], [793], [1067], [1082], [1184] | Non | Non |
| Mathematical models for MERS | [16], [36], [40], [431], [434], [436], [455], [463], [1137] | [19], [27], [65], [382], [835], [1204] | [12], [50], [432], [530], [654], [1154] | [1083] | Non | [485] |
| MERS & mental health | Non | Non | [143], [148], [153], [168], [321], [359], [457], [459], [471], [473], [618], [710], [711], [965], [967], [1138], [1167], [1260] | [519], [661], [709], [773], [786], [828], [907], [908] | Non | Non |
| Surveillance systems | [380] | [273], [468] | [30], [523], [1130] | [319], [322], [871], [945], [1042], [1127], [1128] | Non | Non |
| Case management | [182], [1025] | Non | [665], [1122], [1155] | [94], [291], [322], [476], [477], [478], [665], [822], [832], [845], [869], [891], [892], [837], [1179] | Non | Non |
| MERS-CoV & SARS-CoV-2 cross-reactive immunity | [187], [277], [294], [856], [1053], [1092] | [1245] | [335] | [105], [925], [1066], [1067] | Non | Non |
| Silent/asymptomatic | [410] | [1204] | [155], [726], [1057] | [51], [145], [418], [526], [1080] | Non | [734] |
| Comorbidities | [240], [252], [644] | Non | [243], [388], [524], [542], [618], [772] | [94], [96], [196], [217], [234], [290], [291], [342], [353], [576], [579], [719], [732], [752], [760], [789], [795], [1067], [1223] | Non | [734] |
| MERS & COVID-19 | [248], [252] | [1188] | [256] | [215], [217], [287], [732] | Non | Non |
| One Health approach | [397] | [286] | Non | [517] | [374] | Non |
| Traditional medicine | Non | [652], [682], [860], [1147] | Non | Non | Non | Non |
| MERS & pregnancy | Non | Non | [404], [737] | [594], [794] | [784] | Non |
| Trade and animal food production | Non | Non | Non | [338], [789], [1080] | Non | Non |
| Reinfection | [410], [623] | Non | Non | [529], [696] | Non | Non |

Mapping of MERS-CoV-related research themes by scope level, 1 January 2012 to 24 January 2023 (**Appendix S1: Technical Appendix Reference)**

|  | **Animal** | **Human** | **Animal-human interface** | **Environment** |
| --- | --- | --- | --- | --- |
| Molecular genetics | [24], [35], [37], [52], [59], [118], [165], [167], [181], [202], [230], [232], [235], [373], [428], [438], [454], [494], [499], [500], [507], [692] | [1], [2], [9], [17], [20], [21], [23], [25], [44], [45], [49], [53], [57], [67], [69], [87], [100], [108], [110], [115], [118], [120], [121], [126], [150], [151], [158], [169], [171], [180], [183], [185], [188], [191], [200], [207], [237], [240], [241], [248], [250], [261], [262], [264], [265], [266], [270], [271], [280], [281], [282], [283], [284], [285], [289], [292], [300], [301], [302], [303], [304], [305], [306], [307], [310], [311], [312], 314], [317], [324], [335], [336], [349], [360], [363], 364], [365], [366], [367], [384], [385], [401], [402], [403], [413], [415], [425], [ 426], [427], [446], [451], [460], [461], [464], [465], [469], [486], [487], [489], [491], [492], [493], [495], [496], [497], [498], [501], [513], [532], [534], [539], [540], [554], [555], [557], [558], [559], [561], [563], [565], [568], [570], [571], [572], [573], [574], [577], [578], [585], [592], [596], [608], [615], [616], [622], [626], [631], [632], [633], [634], [635], [637], [638], [644], [648], [657], [658], [662], [674], [678], [681], [686], [687], [688], [697], [698], [702], [703], [708], [712], [716], [722], [723], [724], [725], [728], [735], [738], [739], [742], [743], [744], [745], [747], [753], [757], [759], [763], [765], [768], [776], [779], [783], [785], [788], [783], [785], [788], [804], [807], [808], [809], [810], [811], [812], [813], [818], [821], [824], [825], [829], [834], [836], [837], [838], [840], [841, [842], [843], [844], [847], [848, [852], [853], [854], [855], [857], [858], [863], [864, [868], [873], [874], [876], [877], [879], [880], [886], [894], [895], [897], [902], [903], [905], [911], [916], [917], [919], [920], [921], [922], [923], [924], [929], [930], [931], [933], [939], [953], [962], [963], [964], [972], [974], [983], [984], [987], [998], [1001], [1003], [1004], [1005], [1008], [1011], [1026], [1029], [1030], [1036], [1038], [1039], [1048], [1049], [1053], [1059], [1075], [1077], [1085], [1087], [1088], [1091], [1097], [1098], [1099], [1100], [1105], [1106], [1107], [1108], [1110], [1112], [1113, [1116], [1117], [1118], [1119], [1120], [1121], [1124], [1126], [1143], [1144], [1145], [1149], [1150], [1152], [1156], [1157], [1158], [1164], [1165], [1168], [1172], [1176], [1177], [1182], [1186], [1194], [1195], [1200], [1203], [1205], [1210], [1211], [1212], [1216], [1218], [1230], [1234], [1236], [1238], [1239], [1240], [1246], [1256], [1259], [1261] | [22], [99], [186], [209], [233], [239], [247], [332], [333], [334], [361], [368], [370], [374], [397], [449], [450], [453], [466], [488], [490], [502], [503], [504], [508], [541, [549], [659], [660], [689], [701], [717], [718], [755], [756], [780], [790], [791], [792], [803], [814], [819], [915], [992], [993], [1046], [1084], [1086], [1109], [1114], [1134], [1146], [1171], [1183], [1226], [1227], [1232], [1254], [1263] | [1201] |
| Therapeutics (drug-related or others) | [167], [641], [874] | [1], [2], [3], [4], [5], [6], [7], [8], [9], [10], [17], [33], [34], [42], [44], [45], [53], [57], [66], [67], [70], [84], [88], [90], [105], [106], [107], [132], [133], [134], [141], [150], [158], [169], [170], [173], [174], [175], [176], [177], [183], [188], [194], [198], [201], [207], [236], [237], [253], [254], [261], [266], [267], [268], [269], [272], [281], [304], [310], [313], [316], [324], [325], [326], [327], [328], [330], [366], [367], [369], [372], [387], [389], [399], [403], [405], [406], [409], [446], [452], [470], [477], [478], [480], [481], [487], [492], [493], 495], [511], [532], [534], [540], [544], [545], [551], [552], [555], [559], [560], [562], [566], [567], [569], [573], [574], [577], [601], [603], [604], [605], [606], [613], [629], [631], [632], [633], [634], [635], [639], [640], [642], [650], [686], [687], [700], [703], [704], [712], [721], [723], [724], [725], [742], [743], [748], [759], [783], [798], [799], [832], [884], [895], [898], [901], [905], [906], [914], [924], [935], [952], [954], [955], [956], [957], [958], [959], [981], [982], [1000], [1013], [1014], [1027], [1031], [1032], [1033], [1034], [1035], [1041], [1043], [1054], [1072], [1076], [1090], [1095], [1100], [1101], [1102], [1104], [1111], [1113], [1115], [1116], [1117], [1119], [1121], [1124], [1135], [1136], [1142], [1143], [1144], [1145], [1150], [1151], [1153], [1175], [1181], [1205], [1207], [1208], [1211], [1212], [1222], [1223], [1224], [1225], [1233], [1234], [1236], [1238], [1240], [1255], [1261], [1264] | [209], [1146], [1171], [1227] | Non |
| Pathogenesis | [164], [235], [257], [454], [1078] | [12], [14], [15], [18], [39], [46], [47], [61], [68], [81], [91], [92], [94], [96], [97], [98], [109], [145], [149], [157], [170], [178], [184], [188], [196], [197], [200], [207], [208], [211], [212], [213], [214], [216], [218], [219], [221], [222], [223], [224], [225], [226], [227], [229], [231], [234], 236], [242], [243], [244], [245], [249], [250], [258], [262], [283], [284], [285], [290], [291], [308], [315], [316], [320], [323], [342], [349], [353], [360], [363], [369], [378], [381], [401], [402], [417], [418], [419], [421], [422], [424], [435], [472], [476], [482], [483], [491], [496], [515], [521], [531], [533], [535], [538], [542], [550], [554], [572], [578], [579], [580], [581], [582], [595], [597], [598], [607], [615], [616], [622], [625], [627], [655], [664], [665], [693], [694], [695], [697], [705], [714], [719], [722], [723], [726], [727], [728], [732], [734], [735], [738], [739], [745], [747], [760], [763], [765], [769], [776], [779], [783], [785], [788], [795], [801], [809], [812], [822], [832], [845], [846], [869, [891], [892], [904], [936], [937], [943], [944], [1025], [1028], [1037], [1057], [1068], [1069], 1077], [1112], [1122], [1126], [1155], [1164], [1165], [1168], [1172], [1176], [1177], [1182], [1203], [1209], [1211], [1212], [1220], [1223], [1231], [1236], [1242], [1252] | [526], [752], [789], [1147], [1183] | Non |
| Vaccinology & Immunization | [172], [192], [556], [586], [587], [588], [609], [696], [715], [882], [997], [1209], [1215] | [9], [43],[48], [49], [55], [56], [58], [64], [67], [71], [72], [74], [75], [76], [77], [80], [83], [84], [86], [121], [129], [130], [137], [138], [163], [189], [192], [195], [199], [201], [206], [255], [270], [288], [293], [305], [314], [318], [329], [351], [352], [371], [375], [384], [406], [407], [408], [412], [425], [426], [436], [452], [489], [506], [522], [548], [551], [553], [563], [564], [567], [583], [584], [587], [591], [592], [596], [602, [626], [638], [645], [646], [647], [649], [663], [690], [691], [693], [694], [695], [723], [724], [726], [736], [749], [750], [753], [758], [817], [821], [856], [861], [862], [872], [882], [885], [899], [900], [913], [960], [961], [971], [987], [994], [995], [996], [997], [1044], [1045], [1049], [1054], [1071], [1073], [1092], [1102], [1103], [1125], [1133], [1140], [1141], [1150], [1152], [1187], [1195], [1196], [1202], [1213], [1238] | [397], [508] | Non |
| Seroprevalence | [26], [89], [95], [127], [128], [193], [232], [279], [295], [298], [341], [379], [415], [445], [499], [527], [529], [696], [713], [715], [731], [733], [766], [767], [768], [781], [782], 796], 797], [800], [805], [816], [830], [928], [941], [946], [947], [1015], [1017], [1050], [1051], [1052], [1055], [1058], [1059], [1060], [1061], [1062], [1065], [1139], [1190], [1191], [1192] | [116], [129], [155], [234], [243, [251], [259], [277], [280], [294], [299], [319], [322], [423], [435], [443], [448], [483], [516], [518], [538], [546], [547], [607], [624], [629], [630], [653], [666], [675], [677], [679], [683], [684], [693], [694], [695], [754], [760], [764], [802], [806], [842], [851], [857], [912], [942], [1018], [1042], [1056], [1057], [1063], [1064], [1066], [1127], [1128], [1129], [1189], [1217], [1219] | [233], [286], [296], [297], [374], [420], [484], [504], [509], [525], [526], [651], [680], [682], [699], [771], [775], [791], [792], [815], [867], [878], [1016], [1021], [1079] | Non |
| Transmissibility | [125], [172], [415], [462], [623], [681], [707], [731], [766], [787], [830], [834], [837], [917], [928], [934] | [13], [19], [31], [40], [50], [60], [65], [73], [78], [82], [103], [104], [119], [122], [146], [191], [258], [388], [391], [394], [398], [414], [429], [432], [451], [455], [463], [467], [514], [516], [518], [530], [536], [575], [599], [643], [655], [683], [720], [746], [777], [810], [818], [823], [824], [825], [831], [840], [850], [859], [865], [866], [870], [871], [876], [887], [889], [918], [926], [1005], [1006], [1008], [1016], [1022], [1024], [1137], [1154], [1155], [1186], [1199], [1220], [1237], [1250] | [239], [485], [991], [1002], [1017], [1019], [1020], [1021], [1204], [1241] | [146], [729], [1023], [1247], [926], [1251] |
| Outbreak investigation | [69], [529] | [13], [32], [38], [39], [51], [68], [69], [93], [95], [116], [156], [157], [184], [197], [213], [214], [216], [218], [219], [220], [221], [222], [223, [225], [231], [273], [274], [275], [276], [315], [322], [339], [411], [416], [419], [421], [422], [423], [443], [468], [483], [510], [515], [518], [524], [529], [535], [536], [538], [543], [597], [598], [666], [676], [683], [720], [730], [746], [762], [770], [778], [793], [820] [839], [850], [851], [887], [888], [889], [890], [893], [896], [904], [951], [1009], [1010], [1024], [1040], [1056], [1155], [1214], [1217], [1219], [1221], [1242], [1252] | [233], [239], [528], [549], [1221] | [1221] |
| Laboratory diagnostics | [79], [441], [442], [445], [1070] | [11], [54], [59], [63], [77], [79], [108], [114], [117], [123], [131], [139], [141], [142], [189], [203], [204], [205], [228], [238], [246], [251], [259], [260], [271], [278], [331], [336], [337], [344], [345], [346], [347], [348], [350], [354], [355], [356], [357], [358], [437], [441], [442], [443], [447], [479], [494], [512], [518], [600], [610], [614], [636], [673], [833], [849], [854], [875], [883], [909], [910], [974], [978], [979], [980], [984] [985], [986], [989], [990], [1012], [1028], [1047], [1051], [1070], [1089], [1093], [1123], [1130], [1206], [1235], [1244], [1248], [1249] | [528], [674], [999], [1054], [1232] | Non |
| Anthropology and Social behavior | Non | [28], [29], [62], [101], [111], [113], [140], [147], [152], [160], [161], [162], [166], [168], [256], [263], [343], [362], [390], [396], [439], [440], [444], [457], [458], [474], [475], [537], [543], [628], [656], [661], [667], [668], [669], [671], [672], [685], [706], [709], [710], [740], [761], [786], [826], [827], [828], [831], [907], [908], [948], [949], [966], [968], [970], [975], [976], [1160], [1161], [1162], [1163], [1167], [1174], [1185], [1193], [1198], [1228], [1229], [1257], [1260], [1262] | [670] | Non |
| Spillover | Non | Non | [41], [99], [239], [332], [333], [334], [340], [368], [370], [374], [376], [382], [397], [420], [449], [466], [484], [485], [488], [490], [504], [509], [517], [525], [526], [528], [541], [549], [659], [660], [680, [689], [699], [717], [718], [751], [755], [756], [771], [774], [780, [790], [791], [792], [814], [991], [992], [1002], [1019], [1020], [1021], [1079], [1080], [1086], [1114], [1178], [1180], [1204], [1226], [1241], [1254], [1263] | [41], [682] |
| Outbreak preparedness and response | Non | [16], [30], [31], [73], [102], [103], [124], [136], [144], [159], [182], [190], [278], [353], [377], [380], [381], [386], [388], [391], [392], [396], [400], [429], [430], [431], [432], [434], [436], [463], [469], [479], [543], [598], [643], [654], [673], [676], [707], [741], [890], [896], [908], [926], [940], [945], [969], [1010], [1083], [1127], [1128], [1130], [1148], [1159], [1169], [1243] | [135], [517] | [210], [729], [926] |
| Infection prevention and Control (IPC) | Non | [18], [60], [62], [111], [113], [152], [159], [160], [161], [191], [362], [393], [398], [414], [456], [520], [604], [611], [612], [619], [620], [621], [671], [672], [684], [706], [736], [777], [820], [870], [871], [889], [890], [893], [938], [948], [975], [977], [1018], [1064], [1091], [1126], [1132], [1179], [1200], [1247], [1251], [1253] | [610] | [210], [820] |
| Impact on health system\BOD & economic impact | Non | [14], [15], [94], [96], [179], [196], [214], [223], [226], [242], [290], [291], [309], [320], [322], [342], [383], [395], [422], [424], [430], [431], [433], [435], [472], [510], [524], [536], [538], [579], [595], [627], [772], [778], [793], [801], [1007], [1067], [1082], [1094], [1170], [1173], [1184], [1197], [1258] | [752], | Non |
| Mathematical models for MERS | Non | [12], [16], [19], [27], [31], [36], [40], [50], [65], [73], [78], [136], [144], [146], [394], [429], [430], [431], [432], [434], [436], [455], [461], [463], [530], [599], [654], [932], [950], [988], [1083], [1137], [1154], [1243] | [135], [382], [485], [835], [973], [1204] | [146], [932] |
| Comorbidities | Non | [94], [96], [196], [217], [234], [240], [242], [243], [245], [252], [290], [291], [342], [353], [378], [388], [524], [542], [576], [579], [593], [617], [618], [644], [705], [719], [727], [732], [734], [760], [772], [789], [795], [1067], [1081], [1197], [1223], [1252], [1258] | [752], [789] | Non |
| Surveillance systems | Non | [30], [93], [273], [319], [322], [380], [392], [411], [448], [468], [514], [523], [859], [823], [871], [918], [945], [1042], [1074], [1127], [1128], [1130] | Non | Non |
| MERS-CoV & SARS-CoV-2 cross-reactive immunity | Non | [105], [112], [187], [277], [280], [292], [294], [335], [506], [589], [590], [853], [856], [925], [1053], [1066], [1067], [1092], [1097], [1245] | Non | Non |
| Silent/asymptomatic | [154], [1080], [1231] | [51], [145], [155], [410], [418], [424], [726], [734], [823], [1057], [1080] | [526], [1080], [1204] | Non |
| One Health approach | Non | Non | [41], [286], [374], [397], [484], [504], [517], [881], [1131] | [41], [1131] |
| MERS & mental health | Non | [143], [148], [153], [168], [321], [359], [457], [459], [471], [473], [519], [618], [661], [709], [710], [711], [773], [786], [828], [907], [908], [965], [967], [1138], [1167, [1260] | Non | Non |
| Case management | Non | [94], [182], [224], [291], [322], [392], [476], [477], [478], [665], [666], [822], [832], [845], [869], [891], [892], [937], [1025], [1122], [1155], [1179] | Non | Non |
| MERS & COVID-19 | Non | [215], [217], [248], [249], [252], [254], [256], [264], [287], [440], [732], [1188] | Non | Non |
| Traditional medicine | Non | [652], [860], [927] | [775], [1147] | [682] |
| MERS & pregnancy | Non | [404], [594], [737], [784], [794], [1096] | Non | Non |
| Trade and animal food production | [774], [1080] | [789], [1221] | [338], [774], [789], [1080], [1221] | [1221] |
| Reinfection | [529], [623], [696] | [410] | [528] | Non |
