## Supplementary material for "Mapping the Middle East Respiratory Syndrome (MERS) related Research – A Scoping Review (2012-2023)": Technical appendix

List of the 1,264 articles characterized in the scoping review

169. Binding of the methyl donor S-adenosyl-Lmethionine to Middle East respiratory syndrome coronavirus 2’-Omethyltransferase nsp16 promotes recruitment of the allosteric activator nsp10.

1000. Gordon CJ, Tchesnokov EP, Woolner E, Perry JK, Feng JY, Porter DP, et al. Remdesivir is a direct-acting antiviral that inhibits RNA-dependent RNA polymerase from severe acute respiratory syndrome coronavirus 2 with high potency. J Biol Chem [Internet]. 2020;295(20):6785–97. Available from: http://dx.doi.org/10.1074/jbc.RA120.013679

1001. Munster VJ, Adney DR, van Doremalen N, Brown VR, Miazgowicz KL, Milne-Price S, et al. Replication and shedding of MERS-CoV in Jamaican fruit bats (Artibeus jamaicensis). Sci Rep [Internet]. 2016;6(1):21878. Available from: http://dx.doi.org/10.1038/srep21878

1002. Adney DR, van Doremalen N, Brown VR, Bushmaker T, Scott D, de Wit E, et al. Replication and shedding of MERS-CoV in upper respiratory tract of inoculated dromedary camels. Emerg Infect Dis [Internet]. 2014;20(12):1999–2005. Available from: http://dx.doi.org/10.3201/eid2012.141280

1003. Lau SKP, Fan RYY, Luk HKH, Zhu L, Fung J, Li KSM, et al. Replication of MERS and SARS coronaviruses in bat cells offers insights to their ancestral origins. Emerg Microbes Infect [Internet]. 2018;7(1):209. Available from: http://dx.doi.org/10.1038/s41426-018-0208-9

1004. Eckerle I, Corman VM, Müller MA, Lenk M, Ulrich RG, Drosten C. Replicative capacity of MERS Coronavirus in livestock cell lines. Emerg Infect Dis [Internet]. 2014;20(2):276–9. Available from: http://dx.doi.org/10.3201/eid2002.131182

1005. Park WB, Poon LLM, Choi S-J, Choe PG, Song K-H, Bang JH, et al. Replicative virus shedding in the respiratory tract of patients with Middle East respiratory syndrome coronavirus infection. Int J Infect Dis [Internet]. 2018;72:8–10. Available from: http://dx.doi.org/10.1016/j.ijid.2018.05.003

1006. Conzade R, Grant R, Malik MR, Elkholy A, Elhakim M, Samhouri D, et al. Reported direct and indirect contact with dromedary camels among laboratory-confirmed MERS-CoV cases. Viruses [Internet]. 2018;10(8). Available from: http://dx.doi.org/10.3390/v10080425

1007. Veater J, Wong N, Stephenson I, Kirk-Granger H, Baxter LF, Cannon R, et al. Resource impact of managing suspected Middle East respiratory syndrome patients in a UK teaching hospital. J Hosp Infect [Internet]. 2017;95(3):280–5. Available from: http://dx.doi.org/10.1016/j.jhin.2016.12.010

1008. Memish ZA, Al-Tawfiq JA, Makhdoom HQ, Assiri A, Alhakeem RF, Albarrak A, et al. Respiratory tract samples, viral load, and genome fraction yield in patients with Middle East respiratory syndrome. J Infect Dis [Internet]. 2014;210(10):1590–4. Available from: http://dx.doi.org/10.1093/infdis/jiu292

1009. Mercier A, Méheut A, Alidjinou EK, Lazrek M, Faure K, Hober D, et al. Respiratory virus detection in returning travelers and pilgrims from the Middle East. Travel Med Infect Dis [Internet]. 2023;51(102482):102482. Available from: http://dx.doi.org/10.1016/j.tmaid.2022.102482

1010. Al Hosani FI, Pringle K, Al Mulla M, Kim L, Pham H, Alami NN, et al. Response to emergence of middle east respiratory syndrome Coronavirus, Abu Dhabi, United Arab Emirates, 2013–2014. Emerg Infect Dis [Internet]. 2016;22(7):1162–8. Available from: http://dx.doi.org/10.3201/eid2207.160040

1011. Scobey T, Yount BL, Sims AC, Donaldson EF, Agnihothram SS, Menachery VD, et al. Reverse genetics with a full-length infectious cDNA of the Middle East respiratory syndrome coronavirus. Proc Natl Acad Sci U S A [Internet]. 2013;110(40):16157–62. Available from: http://dx.doi.org/10.1073/pnas.1311542110

1012. Abd El Wahed A, Patel P, Heidenreich D, Hufert FT, Weidmann M. Reverse transcription recombinase polymerase amplification assay for the detection of middle East respiratory syndrome coronavirus. PLoS Curr [Internet]. 2013;5. Available from: http://dx.doi.org/10.1371/currents.outbreaks.62df1c7c75ffc96cd59034531e2e8364

1013. Omrani AS, Saad MM, Baig K, Bahloul A, Abdul-Matin M, Alaidaroos AY, et al. Ribavirin and interferon alfa-2a for severe Middle East respiratory syndrome coronavirus infection: a retrospective cohort study. Lancet Infect Dis [Internet]. 2014;14(11):1090–5. Available from: http://dx.doi.org/10.1016/S1473-3099(14)70920-X

1014. Arabi YM, Shalhoub S, Mandourah Y, Al-Hameed F, Al-Omari A, Al Qasim E, et al. Ribavirin and interferon therapy for critically ill patients with Middle East respiratory syndrome: A multicenter observational study. Clin Infect Dis [Internet]. 2020;70(9):1837–44. Available from: http://dx.doi.org/10.1093/cid/ciz544

1015. Miguel E, Chevalier V, Ayelet G, Ben Bencheikh MN, Boussini H, Chu DK, et al. Risk factors for MERS coronavirus infection in dromedary camels in Burkina Faso, Ethiopia, and Morocco, 2015. Euro Surveill [Internet]. 2017;22(13). Available from: http://dx.doi.org/10.2807/1560-7917.ES.2017.22.13.30498

1016. Khudhair A, Killerby ME, Al Mulla M, Abou Elkheir K, Ternanni W, Bandar Z, et al. Risk factors for MERS-CoV seropositivity among animal market and slaughterhouse workers, Abu Dhabi, United Arab Emirates, 2014-2017. Emerg Infect Dis [Internet]. 2019;25(5):927–35. Available from: http://dx.doi.org/10.3201/eid2505.181728

1017. Holloway P, Gibson M, van Doremalen N, Nash S, Holloway T, Letko M, et al. Risk factors for Middle East respiratory syndrome Coronavirus infection among camel populations, southern Jordan, 2014-2018. Emerg Infect Dis [Internet]. 2021;27(9):2301–11. Available from: http://dx.doi.org/10.3201/eid2709.203508

1018. Alraddadi BM, Al-Salmi HS, Jacobs-Slifka K, Slayton RB, Estivariz CF, Geller AI, et al. Risk factors for Middle East respiratory syndrome Coronavirus infection among healthcare personnel. Emerg Infect Dis [Internet]. 2016;22(11):1915–20. Available from: http://dx.doi.org/10.3201/eid2211.160920

1019. Alraddadi BM, Watson JT, Almarashi A, Abedi GR, Turkistani A, Sadran M, et al. Risk factors for primary Middle East respiratory syndrome Coronavirus illness in humans, Saudi Arabia, 2014. Emerg Infect Dis [Internet]. 2016;22(1):49–55. Available from: http://dx.doi.org/10.3201/eid2201.151340

1020. Sikkema RS, Farag EABA, Himatt S, Ibrahim AK, Al-Romaihi H, Al-Marri SA, et al. Risk factors for primary Middle East respiratory syndrome Coronavirus infection in camel workers in Qatar during 2013-2014: A case-control study. J Infect Dis [Internet]. 2017;215(11):1702–5. Available from: http://dx.doi.org/10.1093/infdis/jix174

1021. Sitawa R, Folorunso F, Obonyo M, Apamaku M, Kiambi S, Gikonyo S, et al. Risk factors for serological evidence of MERS-CoV in camels, Kenya, 2016-2017. Prev Vet Med [Internet]. 2020;185(105197):105197. Available from: http://dx.doi.org/10.1016/j.prevetmed.2020.105197

1022. Kim SW, Park JW, Jung H-D, Yang J-S, Park Y-S, Lee C, et al. Risk factors for transmission of Middle East respiratory syndrome coronavirus infection during the 2015 outbreak in South Korea. Clin Infect Dis [Internet]. 2017;64(5):551–7. Available from: http://dx.doi.org/10.1093/cid/ciw768

1023. Gardner LM, Chughtai AA, MacIntyre CR. Risk of global spread of Middle East respiratory syndrome coronavirus (MERS-CoV) via the air transport network. J Travel Med [Internet]. 2016;23(6):taw063. Available from: http://dx.doi.org/10.1093/jtm/taw063

1024. Ki HK, Han SK, Son JS, Park SO. Risk of transmission via medical employees and importance of routine infection-prevention policy in a nosocomial outbreak of Middle East respiratory syndrome (MERS): a descriptive analysis from a tertiary care hospital in South Korea. BMC Pulm Med [Internet]. 2019;19(1):190. Available from: http://dx.doi.org/10.1186/s12890-019-0940-5

1025. Rivers CM, Majumder MS, Lofgren ET. Risks of death and severe disease in patients with middle east respiratory syndrome Coronavirus, 2012–2015. Am J Epidemiol [Internet]. 2016;184(6):460–4. Available from: http://dx.doi.org/10.1093/aje/kww013

1026. Rojas-Cruz AF, Gallego-Gómez JC, Bermúdez-Santana CI. RNA structure-altering mutations underlying positive selection on Spike protein reveal novel putative signatures to trace crossing host-species barriers in Betacoronavirus. RNA Biol [Internet]. 2022;19(1):1019–44. Available from: http://dx.doi.org/10.1080/15476286.2022.2115750

1027. Kutkat O, Moatasim Y, Al-Karmalawy AA, Abulkhair HS, Gomaa MR, El-Taweel AN, et al. Robust antiviral activity of commonly prescribed antidepressants against emerging coronaviruses: in vitro and in silico drug repurposing studies. Sci Rep [Internet]. 2022;12(1):12920. Available from: http://dx.doi.org/10.1038/s41598-022-17082-6

1028. Das KM, Alkoteesh JA, Sheek-Hussein M, Alzadjali SA, Alafeefi MT, Singh R, et al. Role of chest radiograph in MERS-Cov pneumonia: a single tertiary referral center experience in the United Arab Emirates. Egypt J Radiol Nucl Med [Internet]. 2021;52(1). Available from: http://dx.doi.org/10.1186/s43055-021-00517-x

1029. Qian Z, Dominguez SR, Holmes KV. Role of the spike glycoprotein of human Middle East respiratory syndrome coronavirus (MERS-CoV) in virus entry and syncytia formation. PLoS One [Internet]. 2013;8(10):e76469. Available from: http://dx.doi.org/10.1371/journal.pone.0076469

1030. Corman VM, Ithete NL, Richards LR, Schoeman MC, Preiser W, Drosten C, et al. Rooting the phylogenetic tree of middle East respiratory syndrome coronavirus by characterization of a conspecific virus from an African bat. J Virol [Internet]. 2014;88(19):11297–303. Available from: http://dx.doi.org/10.1128/JVI.01498-14

1031. Folegatti PM, Bittaye M, Flaxman A, Lopez FR, Bellamy D, Kupke A, et al. Safety and immunogenicity of a candidate Middle East respiratory syndrome coronavirus viral-vectored vaccine: a dose-escalation, open-label, non-randomised, uncontrolled, phase 1 trial. Lancet Infect Dis [Internet]. 2020;20(7):816–26. Available from: http://dx.doi.org/10.1016/S1473-3099(20)30160-2

1032. Koch T, Dahlke C, Fathi A, Kupke A, Krähling V, Okba NMA, et al. Safety and immunogenicity of a modified vaccinia virus Ankara vector vaccine candidate for Middle East respiratory syndrome: an open-label, phase 1 trial. Lancet Infect Dis [Internet]. 2020;20(7):827–38. Available from: http://dx.doi.org/10.1016/S1473-3099(20)30248-6

1033. Modjarrad K, Roberts CC, Mills KT, Castellano AR, Paolino K, Muthumani K, et al. Safety and immunogenicity of an anti-Middle East respiratory syndrome coronavirus DNA vaccine: a phase 1, open-label, single-arm, dose-escalation trial. Lancet Infect Dis [Internet]. 2019;19(9):1013–22. Available from: http://dx.doi.org/10.1016/S1473-3099(19)30266-X

1034. Beigel JH, Voell J, Kumar P, Raviprakash K, Wu H, Jiao J-A, et al. Safety and tolerability of a novel, polyclonal human anti-MERS coronavirus antibody produced from transchromosomic cattle: a phase 1 randomised, double-blind, single-dose-escalation study. Lancet Infect Dis [Internet]. 2018;18(4):410–8. Available from: http://dx.doi.org/10.1016/s1473-3099(18)30002-1

1035. Shin J, Jung E, Kim M, Baric R, Go Y. Saracatinib inhibits middle east respiratory syndrome-Coronavirus replication in vitro. Viruses [Internet]. 2018;10(6):283. Available from: http://dx.doi.org/10.3390/v10060283

1036. Oh L, Varki A, Chen X, Wang L-P. SARS-CoV-2 and MERS-CoV spike protein binding studies support stable mimic of bound 9-O-acetylated sialic acids. Molecules [Internet]. 2022;27(16):5322. Available from: http://dx.doi.org/10.3390/molecules27165322

1037. Byrd-Leotis L, Lasanajak Y, Bowen T, Baker K, Song X, Suthar MS, et al. SARS-CoV-2 and other coronaviruses bind to phosphorylated glycans from the human lung. Virology [Internet]. 2021;562:142–8. Available from: http://dx.doi.org/10.1016/j.virol.2021.07.012

1038. Yang S, Zhou H, Liu M, Jaijyan D, Cruz-Cosme R, Ramasamy S, et al. SARS-CoV-2, SARS-CoV, and MERS-CoV encode circular RNAs of spliceosome-independent origin. J Med Virol [Internet]. 2022;94(7):3203–22. Available from: http://dx.doi.org/10.1002/jmv.27734

1039. Alnaeem A, Kasem S, Qasim I, Refaat M, Alhufufi AN, Al-Doweriej A, et al. Scanning electron microscopic findings on respiratory organs of some naturally infected dromedary camels with the lineage-B of the middle east respiratory syndrome Coronavirus (MERS-CoV) in Saudi Arabia-2018. Pathogens [Internet]. 2021;10(4):420. Available from: http://dx.doi.org/10.3390/pathogens10040420

1040. Alanazi KH, Killerby ME, Biggs HM, Abedi GR, Jokhdar H, Alsharef AA, et al. Scope and extent of healthcare-associated Middle East respiratory syndrome coronavirus transmission during two contemporaneous outbreaks in Riyadh, Saudi Arabia, 2017. Infect Control Hosp Epidemiol [Internet]. 2019;40(1):79–88. Available from: http://dx.doi.org/10.1017/ice.2018.290

1041. Ali Dahhas M, M Alkahtani H, Malik A, Almehizia AA, Bakheit AH, Akber Ansar S, et al. Screening and identification of potential MERS-CoV papain-like protease (PLpro) inhibitors; Steady-state kinetic and Molecular dynamic studies. Saudi Pharm J [Internet]. 2023;31(2):228–44. Available from: http://dx.doi.org/10.1016/j.jsps.2022.12.007

1042. Memish ZA, Al-Tawfiq JA, Makhdoom HQ, Al-Rabeeah AA, Assiri A, Alhakeem RF, et al. Screening for Middle East respiratory syndrome coronavirus infection in hospital patients and their healthcare worker and family contacts: a prospective descriptive study. Clin Microbiol Infect [Internet]. 2014;20(5):469–74. Available from: http://dx.doi.org/10.1111/1469-0691.12562

1043. de Wilde AH, Jochmans D, Posthuma CC, Zevenhoven-Dobbe JC, van Nieuwkoop S, Bestebroer TM, et al. Screening of an FDA-approved compound library identifies four small-molecule inhibitors of Middle East respiratory syndrome coronavirus replication in cell culture. Antimicrob Agents Chemother [Internet]. 2014;58(8):4875–84. Available from: http://dx.doi.org/10.1128/AAC.03011-14

1044. Ma C, Wang L, Tao X, Zhang N, Yang Y, Tseng C-TK, et al. Searching for an ideal vaccine candidate among different MERS coronavirus receptor-binding fragments—The importance of immunofocusing in subunit vaccine design. Vaccine [Internet]. 2014;32(46):6170–6. Available from: http://dx.doi.org/10.1016/j.vaccine.2014.08.086

1045. Kim Y, Lee H, Park K, Park S, Lim J-H, So MK, et al. Selection and characterization of monoclonal antibodies targeting Middle East respiratory syndrome Coronavirus through a human synthetic Fab phage display library panning. Antibodies (Basel) [Internet]. 2019;8(3):42. Available from: http://dx.doi.org/10.3390/antib8030042

1046. Banerjee A, Subudhi S, Rapin N, Lew J, Jain R, Falzarano D, et al. Selection of viral variants during persistent infection of insectivorous bat cells with Middle East respiratory syndrome coronavirus. Sci Rep [Internet]. 2020;10(1):7257. Available from: http://dx.doi.org/10.1038/s41598-020-64264-1

1047. Okba NMA, Raj VS, Widjaja I, GeurtsvanKessel CH, de Bruin E, Chandler FD, et al. Sensitive and specific detection of low-level antibody responses in mild middle East respiratory syndrome Coronavirus infections. Emerg Infect Dis [Internet]. 2019;25(10):1868–77. Available from: http://dx.doi.org/10.3201/eid2510.190051

1048. Farrag MA, Amer HM, Bhat R, Almajhdi FN. Sequence and phylogentic analysis of MERS-CoV in Saudi Arabia, 2012-2019. Virol J [Internet]. 2021;18(1):90. Available from: http://dx.doi.org/10.1186/s12985-021-01563-7

1049. Kim Y-S, Aigerim A, Park U, Kim Y, Rhee J-Y, Choi J-P, et al. Sequential emergence and wide spread of neutralization escape Middle East respiratory syndrome Coronavirus mutants, South Korea, 2015. Emerg Infect Dis [Internet]. 2019;25(6):1161–8. Available from: http://dx.doi.org/10.3201/eid2506.181722

1050. Thwiny HT, Department of Veterinary Microbiology and Parasitology, College of Veterinary Medicine, University of Basrah, Basrah, Iraq, Al Hamed TA, Nazzal AR. Seroepidemiological study of Middle East Respiratory Syndrome (MERS) virus infection in Iraqi dromedary camels. Vet Arh [Internet]. 2018;88(2):191–200. Available from: http://dx.doi.org/10.24099/vet.arhiv.161224

1051. Perera RA, Wang P, Gomaa MR, El-Shesheny R, Kandeil A, Bagato O, et al. Seroepidemiology for MERS coronavirus using microneutralisation and pseudoparticle virus neutralisation assays reveal a high prevalence of antibody in dromedary camels in Egypt, June 2013. Euro Surveill [Internet]. 2013;18(36):ii=20574. Available from: http://dx.doi.org/10.2807/1560-7917.es2013.18.36.20574

1052. Hemida MG, Perera RA, Al Jassim RA, Kayali G, Siu LY, Wang P, et al. Seroepidemiology of Middle East respiratory syndrome (MERS) coronavirus in Saudi Arabia (1993) and Australia (2014) and characterisation of assay specificity. Euro Surveill [Internet]. 2014;19(23). Available from: http://dx.doi.org/10.2807/1560-7917.es2014.19.23.20828

1053. Hicks J, Klumpp-Thomas C, Kalish H, Shunmugavel A, Mehalko J, Denson J-P, et al. Serologic cross-reactivity of SARS-CoV-2 with endemic and seasonal betacoronaviruses. J Clin Immunol [Internet]. 2021;41(5):906–13. Available from: http://dx.doi.org/10.1007/s10875-021-00997-6

1054. Okba NMA, Widjaja I, Li W, GeurtsvanKessel CH, Farag EABA, Al-Hajri M, et al. Serologic detection of Middle East respiratory syndrome Coronavirus functional antibodies. Emerg Infect Dis [Internet]. 2020;26(5):1024–7. Available from: http://dx.doi.org/10.3201/eid2605.190921

1055. Saqib M, Sieberg A, Hussain MH, Mansoor MK, Zohaib A, Lattwein E, et al. Serologic evidence for MERS-CoV infection in dromedary camels, Punjab, Pakistan, 2012–2015. Emerg Infect Dis [Internet]. 2017;23(3):550–1. Available from: http://dx.doi.org/10.3201/eid2303.161285

1056. Al Hosani FI, Kim L, Khudhair A, Pham H, Al Mulla M, Al Bandar Z, et al. Serologic follow-up of Middle East respiratory syndrome Coronavirus cases and contacts-Abu Dhabi, United Arab Emirates. Clin Infect Dis [Internet]. 2019;68(3):409–18. Available from: http://dx.doi.org/10.1093/cid/ciy503

1057. Ko J-H, Müller MA, Seok H, Park GE, Lee JY, Cho SY, et al. Serologic responses of 42 MERS-coronavirus-infected patients according to the disease severity. Diagn Microbiol Infect Dis [Internet]. 2017;89(2):106–11. Available from: http://dx.doi.org/10.1016/j.diagmicrobio.2017.07.006

1058. Olarinmoye AO, Olugasa BO, Niphuis H, Herwijnen RV, Verschoor E, Boug A, et al. Serological evidence of coronavirus infections in native hamadryas baboons (Papio hamadryas hamadryas) of the Kingdom of Saudi Arabia. Epidemiol Infect [Internet]. 2017;145(10):2030–7. Available from: http://dx.doi.org/10.1017/S0950268817000905

1059. Zhang W, Zheng X-S, Agwanda B, Ommeh S, Zhao K, Lichoti J, et al. Serological evidence of MERS-CoV and HKU8-related CoV co-infection in Kenyan camels. Emerg Microbes Infect [Internet]. 2019;8(1):1528–34. Available from: http://dx.doi.org/10.1080/22221751.2019.1679610

1060. Deem SL, Fèvre EM, Kinnaird M, Browne AS, Muloi D, Godeke G-J, et al. Serological evidence of MERS-CoV antibodies in dromedary camels (Camelus dromedaries) in Laikipia County, Kenya. PLoS One [Internet]. 2015;10(10):e0140125. Available from: http://dx.doi.org/10.1371/journal.pone.0140125

1061. Orynbayev MB, Hitch AT, Kerimbayev AA, Nissanova RK, Sultankulova KT, Rystayeva RA, et al. Serological exposure in Bactrian and dromedary camels in Kazakhstan to a MERS or MERS-like coronavirus. Transbound Emerg Dis [Internet]. 2022;69(5):e1374–81. Available from: http://dx.doi.org/10.1111/tbed.14468

1062. Karamendin K, Seidalina A, Sabyrzhan T, Nuralibekov S, Kasymbekov Y, Suleimenova S, et al. Serological screening for Middle East Respiratory Syndrome Coronavirus and hepatitis E virus in camels in Kazakhstan. Pathogens [Internet]. 2022;11(11):1224. Available from: http://dx.doi.org/10.3390/pathogens11111224

1063. Degnah AA, Al-Amri SS, Hassan AM, Almasoud AS, Mousa M, Almahboub SA, et al. Seroprevalence of MERS-CoV in healthy adults in western Saudi Arabia, 2011-2016. J Infect Public Health [Internet]. 2020;13(5):697–703. Available from: http://dx.doi.org/10.1016/j.jiph.2020.01.001

1064. Ryu B, Cho S-I, Oh M-D, Lee J-K, Lee J, Hwang Y-O, et al. Seroprevalence of Middle East respiratory syndrome coronavirus (MERS-CoV) in public health workers responding to a MERS outbreak in Seoul, Republic of Korea, in 2015. Western Pac Surveill Response J [Internet]. 2019;10(2):46–8. Available from: http://dx.doi.org/10.5365/wpsar.2018.9.3.002

1065. Harrath R, Abu Duhier FM. Sero-prevalence of Middle East respiratory syndrome coronavirus (MERS-CoV) specific antibodies in dromedary camels in Tabuk, Saudi Arabia. J Med Virol [Internet]. 2018;90(8):1285–9. Available from: http://dx.doi.org/10.1002/jmv.25186

1066. Alroqi F, Masuadi E, Alabdan L, Nogoud M, Aljedaie M, Abu-Jaffal AS, et al. Seroprevalence of SARS-CoV-2 among high-risk healthcare workers in a MERS-CoV endemic area. J Infect Public Health [Internet]. 2021;14(9):1268–73. Available from: http://dx.doi.org/10.1016/j.jiph.2021.08.029

1067. Elhazmi A, Al-Tawfiq JA, Sallam H, Al-Omari A, Alhumaid S, Mady A, et al. Severe acute respiratory syndrome coronavirus 2 (SARS-CoV-2) and Middle East Respiratory Syndrome Coronavirus (MERS-CoV) coinfection: A unique case series. Travel Med Infect Dis [Internet]. 2021;41(102026):102026. Available from: http://dx.doi.org/10.1016/j.tmaid.2021.102026

1068. Arabi YM, Harthi A, Hussein J, Bouchama A, Johani S, Hajeer AH, et al. Severe neurologic syndrome associated with Middle East respiratory syndrome corona virus (MERS-CoV). Infection [Internet]. 2015;43(4):495–501. Available from: http://dx.doi.org/10.1007/s15010-015-0720-y

1069. Tan M, Wang J, Hu P, Wang B, Xu W, Chen J. Severe pneumonia due to infection with Candida krusei in a case of suspected Middle East respiratory syndrome: A case report and literature review. Exp Ther Med [Internet]. 2016;12(6):4085–8. Available from: http://dx.doi.org/10.3892/etm.2016.3892

1070. Noh JY, Yoon S-W, Kim D-J, Lee M-S, Kim J-H, Na W, et al. Correction to: Simultaneous detection of severe acute respiratory syndrome, Middle East respiratory syndrome, and related bat coronaviruses by real-time reverse transcription PCR. Arch Virol [Internet]. 2018;163(3):819. Available from: http://dx.doi.org/10.1007/s00705-017-3677-6

1071. Jia W, Channappanavar R, Zhang C, Li M, Zhou H, Zhang S, et al. Single intranasal immunization with chimpanzee adenovirus-based vaccine induces sustained and protective immunity against MERS-CoV infection. Emerg Microbes Infect [Internet]. 2019;8(1):760–72. Available from: http://dx.doi.org/10.1080/22221751.2019.1620083

1072. Qiu H, Sun S, Xiao H, Feng J, Guo Y, Tai W, et al. Single-dose treatment with a humanized neutralizing antibody affords full protection of a human transgenic mouse model from lethal Middle East respiratory syndrome (MERS)-coronavirus infection. Antiviral Res [Internet]. 2016;132:141–8. Available from: http://dx.doi.org/10.1016/j.antiviral.2016.06.003

1073. Li K, Li Z, Wohlford-Lenane C, Meyerholz DK, Channappanavar R, An D, et al. Single-dose, intranasal immunization with recombinant Parainfluenza virus 5 expressing Middle East respiratory syndrome Coronavirus (MERS-CoV) spike protein protects mice from fatal MERS-CoV infection. MBio [Internet]. 2020;11(2). Available from: http://dx.doi.org/10.1128/mBio.00554-20

1074. Zhao H, ParryFord F, Dabrera G, Sinnathamby M, Ellis J, Dunning J, et al. Six-year experience of detection and investigation of possible Middle East Respiratory Syndrome coronavirus cases, England, 2012-2018. Public Health [Internet]. 2020;189:141–3. Available from: http://dx.doi.org/10.1016/j.puhe.2020.10.007

1075. Gassen NC, Niemeyer D, Muth D, Corman VM, Martinelli S, Gassen A, et al. SKP2 attenuates autophagy through Beclin1-ubiquitination and its inhibition reduces MERS-Coronavirus infection. Nat Commun [Internet]. 2019;10(1):5770. Available from: http://dx.doi.org/10.1038/s41467-019-13659-4

1076. Kandeel M, Yamamoto M, Al-Taher A, Watanabe A, Oh-Hashi K, Park BK, et al. Small molecule inhibitors of middle East Respiratory Syndrome Coronavirus fusion by targeting cavities on heptad repeat trimers. Biomol Ther (Seoul) [Internet]. 2020;28(4):311–9. Available from: http://dx.doi.org/10.4062/biomolther.2019.202

1077. Totura A, Livingston V, Frick O, Dyer D, Nichols D, Nalca A. Small particle aerosol exposure of African green monkeys to MERS-CoV as a model for highly pathogenic Coronavirus infection. Emerg Infect Dis [Internet]. 2020;26(12):2835–43. Available from: http://dx.doi.org/10.3201/eid2612.201664

1078. Alnaeem A, Kasem S, Qasim I, Al-Doweriej A, Al-Houfufi A, Alwazan A, et al. Some pathological observations on the naturally infected dromedary camels (Camelus dromedarius) with the Middle East respiratory syndrome coronavirus (MERS-CoV) in Saudi Arabia 2018-2019. Vet Q [Internet]. 2020;40(1):190–7. Available from: http://dx.doi.org/10.1080/01652176.2020.1781350

1079. Memish ZA, Alsahly A, Masri MA, Heil GL, Anderson BD, Peiris M, et al. Sparse evidence of MERS-CoV infection among animal workers living in Southern Saudi Arabia during 2012. Influenza Other Respi Viruses [Internet]. 2015;9(2):64–7. Available from: http://dx.doi.org/10.1111/irv.12287

1080. Al-Ahmadi K, Alahmadi M, Al-Zahrani A. Spatial association between primary Middle East respiratory syndrome coronavirus infection and exposure to dromedary camels in Saudi Arabia. Zoonoses Public Health [Internet]. 2020;67(4):382–90. Available from: http://dx.doi.org/10.1111/zph.12697

1081. Adegboye OA, Gayawan E, Hanna F. Spatial modelling of contribution of individual level risk factors for mortality from Middle East respiratory syndrome coronavirus in the Arabian Peninsula. PLoS One [Internet]. 2017;12(7):e0181215. Available from: http://dx.doi.org/10.1371/journal.pone.0181215

1082. Al-Ahmadi KH, Alahmadi MH, Al-Zahrani AS, Hemida MG. Spatial variability of Middle East respiratory syndrome coronavirus survival rates and mortality hazard in Saudi Arabia, 2012–2019. PeerJ [Internet]. 2020;8(e9783):e9783. Available from: http://dx.doi.org/10.7717/peerj.9783

1083. Al-Ahmadi K, Alahmadi S, Al-Zahrani A. Spatiotemporal clustering of middle East respiratory syndrome Coronavirus (MERS-CoV) incidence in Saudi Arabia, 2012-2019. Int J Environ Res Public Health [Internet]. 2019;16(14):2520. Available from: http://dx.doi.org/10.3390/ijerph16142520

1084. Widagdo W, Okba NMA, Li W, de Jong A, de Swart RL, Begeman L, et al. Species-specific colocalization of middle east respiratory syndrome Coronavirus attachment and entry receptors. J Virol [Internet]. 2019;93(16). Available from: http://dx.doi.org/10.1128/jvi.00107-19

1085. Lu X, Rowe LA, Frace M, Stevens J, Abedi GR, Elnile O, et al. Spike gene deletion quasispecies in serum of patient with acute MERS-CoV infection: MERS-CoV Spike Gene Deletion. J Med Virol [Internet]. 2017;89(3):542–5. Available from: http://dx.doi.org/10.1002/jmv.24652

1086. Kleine-Weber H, Pöhlmann S, Hoffmann M. Spike proteins of novel MERS-coronavirus isolates from North- and West-African dromedary camels mediate robust viral entry into human target cells. Virology [Internet]. 2019;535:261–5. Available from: http://dx.doi.org/10.1016/j.virol.2019.07.016

1087. Kim Y, Cheon S, Min C-K, Sohn KM, Kang YJ, Cha Y-J, et al. Spread of mutant Middle East respiratory syndrome Coronavirus with reduced affinity to human CD26 during the South Korean outbreak. MBio [Internet]. 2016;7(2):e00019. Available from: http://dx.doi.org/10.1128/mBio.00019-16

1088. Cotten M, Watson SJ, Zumla AI, Makhdoom HQ, Palser AL, Ong SH, et al. Spread, circulation, and evolution of the Middle East respiratory syndrome coronavirus. MBio [Internet]. 2014;5(1). Available from: http://dx.doi.org/10.1128/mBio.01062-13

1089. Kang A, Yeom M, Kim H, Yoon S-W, Jeong D-G, Moon H-J, et al. Sputum processing method for lateral flow immunochromatographic assays to detect coronaviruses. Immune Netw [Internet]. 2021;21(1):e11. Available from: http://dx.doi.org/10.4110/in.2021.21.e11

1090. Yuan S, Chu H, Chan JF-W, Ye Z-W, Wen L, Yan B, et al. SREBP-dependent lipidomic reprogramming as a broad-spectrum antiviral target. Nat Commun [Internet]. 2019;10(1):120. Available from: http://dx.doi.org/10.1038/s41467-018-08015-x

1091. Abdallah NMA, Zaki AM, Abdel-Salam SA. Stability of MERS-CoV RNA on spin columns of RNA extraction kit at room temperature. Diagn Microbiol Infect Dis [Internet]. 2020;98(4):115182. Available from: http://dx.doi.org/10.1016/j.diagmicrobio.2020.115182

1092. Hsieh C-L, Werner AP, Leist SR, Stevens LJ, Falconer E, Goldsmith JA, et al. Stabilized coronavirus spike stem elicits a broadly protective antibody. Cell Rep [Internet]. 2021;37(5):109929. Available from: http://dx.doi.org/10.1016/j.celrep.2021.109929

1093. Yaren O, Glushakova LG, Bradley KM, Hoshika S, Benner SA. Standard and AEGIS nicking molecular beacons detect amplicons from the Middle East respiratory syndrome coronavirus. J Virol Methods [Internet]. 2016;236:54–61. Available from: http://dx.doi.org/10.1016/j.jviromet.2016.07.008

1094. Mobaraki K, Salamatbakhsh M, Ahmadzadeh J. Standard expected years of life lost as a neglected index for calculating the burden of premature mortality due to middle east respiratory syndrome. Health Secur [Internet]. 2019;17(5):407–9. Available from: http://dx.doi.org/10.1089/hs.2019.0074

1095. Zakaria MY, Fayad E, Althobaiti F, Zaki I, Abu Almaaty AH. Statistical optimization of bile salt deployed nanovesicles as a potential platform for oral delivery of piperine: accentuated antiviral and anti-inflammatory activity in MERS-CoV challenged mice. Drug Deliv [Internet]. 2021;28(1):1150–65. Available from: http://dx.doi.org/10.1080/10717544.2021.1934190

1096. Payne DC, Iblan I, Alqasrawi S, Al Nsour M, Rha B, Tohme RA, et al. Stillbirth during infection with middle east respiratory syndrome Coronavirus. J Infect Dis [Internet]. 2014;209(12):1870–2. Available from: http://dx.doi.org/10.1093/infdis/jiu068

1097. Barman N, De A, Paul J, Haldar S, Bhattacharya A, Pal K. Strategy to Configure Multi-epitope Recombinant Immunogens with Weightage on Proinflamatory Response using SARS-CoV-2 Spike Glycoprotein (S-protein) and RNA-dependent RNA Polymerase (RdRp) as Model Targets. J Pure Appl Microbiol [Internet]. 2022;16(1):281–95. Available from: http://dx.doi.org/10.22207/jpam.16.1.17

1098. Zhang L, Li L, Yan L, Ming Z, Jia Z, Lou Z, et al. Structural and biochemical characterization of endoribonuclease Nsp15 encoded by Middle East respiratory syndrome Coronavirus. J Virol [Internet]. 2018;92(22). Available from: http://dx.doi.org/10.1128/JVI.00893-18

1099. Lin M-H, Chuang S-J, Chen C-C, Cheng S-C, Cheng K-W, Lin C-H, et al. Structural and functional characterization of MERS coronavirus papain-like protease. J Biomed Sci [Internet]. 2014;21(1):54. Available from: http://dx.doi.org/10.1186/1423-0127-21-54

1100. Wrapp D, De Vlieger D, Corbett KS, Torres GM, Wang N, Van Breedam W, et al. Structural basis for potent neutralization of betacoronaviruses by single-domain camelid antibodies. Cell [Internet]. 2020;181(6):1436–41. Available from: http://dx.doi.org/10.1016/j.cell.2020.05.047

1101. Hu X, Lin C, Xu Q, Zhou X, Zeng P, McCormick PJ, et al. Structural basis for the inhibition of coronaviral main proteases by a benzothiazole-based inhibitor. Viruses [Internet]. 2022;14(9). Available from: http://dx.doi.org/10.3390/v14092075

1102. Yu X, Zhang S, Jiang L, Cui Y, Li D, Wang D, et al. Structural basis for the neutralization of MERS-CoV by a human monoclonal antibody MERS-27. Sci Rep [Internet]. 2015;5(1):13133. Available from: http://dx.doi.org/10.1038/srep13133

1103. Srivastava S, Kamthania M, Singh S, Saxena AK, Sharma N. Structural basis of development of multi-epitope vaccine against Middle East respiratory syndrome using in silico approach. Infect Drug Resist [Internet]. 2018;11:2377–91. Available from: http://dx.doi.org/10.2147/IDR.S175114

1104. Li J, Lin C, Zhou X, Zhong F, Zeng P, Yang Y, et al. Structural basis of the main proteases of Coronavirus bound to drug candidate PF-07321332. J Virol [Internet]. 2022;96(8):e0201321. Available from: http://dx.doi.org/10.1128/jvi.02013-21

1105. Cueno ME, Imai K. Structural comparison of the SARS CoV 2 spike protein relative to other human-infecting coronaviruses. Front Med (Lausanne) [Internet]. 2020;7:594439. Available from: http://dx.doi.org/10.3389/fmed.2020.594439

1106. Zhou H, Chen Y, Zhang S, Niu P, Qin K, Jia W, et al. Structural definition of a neutralization epitope on the N-terminal domain of MERS-CoV spike glycoprotein. Nat Commun [Internet]. 2019;10(1):3068. Available from: http://dx.doi.org/10.1038/s41467-019-10897-4

1107. Wang N, Rosen O, Wang L, Turner HL, Stevens LJ, Corbett KS, et al. Structural definition of a neutralization-sensitive Epitope on the MERS-CoV S1-NTD. Cell Rep [Internet]. 2019;28(13):3395-3405.e6. Available from: http://dx.doi.org/10.1016/j.celrep.2019.08.052

1108. Zhang S, Zhou P, Wang P, Li Y, Jiang L, Jia W, et al. Structural definition of a unique neutralization Epitope on the receptor-binding domain of MERS-CoV spike glycoprotein. Cell Rep [Internet]. 2018;24(2):441–52. Available from: http://dx.doi.org/10.1016/j.celrep.2018.06.041

1109. Daczkowski CM, Dzimianski JV, Clasman JR, Goodwin O, Mesecar AD, Pegan SD. Structural insights into the interaction of Coronavirus papain-like proteases and interferon-stimulated gene product 15 from different species. J Mol Biol [Internet]. 2017;429(11):1661–83. Available from: http://dx.doi.org/10.1016/j.jmb.2017.04.011

1110. Batool M, Shah M, Patra MC, Yesudhas D, Choi S. Structural insights into the Middle East respiratory syndrome coronavirus 4a protein and its dsRNA binding mechanism. Sci Rep [Internet]. 2017;7(1):11362. Available from: http://dx.doi.org/10.1038/s41598-017-11736-6

1111. Theerawatanasirikul S, Kuo CJ, Phecharat N, Chootip J, Lekcharoensuk C, Lekcharoensuk P. Structural-based virtual screening and in vitro assays for small molecules inhibiting the feline coronavirus 3CL protease as a surrogate platform for coronaviruses. Antiviral Res [Internet]. 2020;182:104927. Available from: http://dx.doi.org/10.1016/j.antiviral.2020.104927

1112. Daczkowski CM, Goodwin OY, Dzimianski JV, Farhat JJ, Pegan SD. Structurally guided removal of DeISGylase biochemical activity from papain-like protease originating from Middle East respiratory syndrome Coronavirus. J Virol [Internet]. 2017;91(23). Available from: http://dx.doi.org/10.1128/jvi.01067-17

1113. Gao J, Lu G, Qi J, Li Y, Wu Y, Deng Y, et al. Structure of the fusion core and inhibition of fusion by a heptad repeat peptide derived from the S protein of Middle East respiratory syndrome coronavirus. J Virol [Internet]. 2013;87(24):13134–40. Available from: http://dx.doi.org/10.1128/JVI.02433-13

1114. Han X, Qi J, Song H, Wang Q, Zhang Y, Wu Y, et al. Structure of the S1 subunit C-terminal domain from bat-derived coronavirus HKU5 spike protein. Virology [Internet]. 2017;507:101–9. Available from: http://dx.doi.org/10.1016/j.virol.2017.04.016

1115. Zhang Y, Gao H, Hu X, Wang Q, Zhong F, Zhou X, et al. Structure-based discovery and structural basis of a novel broad-spectrum natural product against the main protease of Coronavirus. J Virol [Internet]. 2022;96(1):e0125321. Available from: http://dx.doi.org/10.1128/JVI.01253-21

1116. Lu L, Liu Q, Zhu Y, Chan K-H, Qin L, Li Y, et al. Structure-based discovery of Middle East respiratory syndrome coronavirus fusion inhibitor. Nat Commun [Internet]. 2014;5(1):3067. Available from: http://dx.doi.org/10.1038/ncomms4067

1117. Lin S-M, Lin S-C, Hsu J-N, Chang C-K, Chien C-M, Wang Y-S, et al. Structure-based stabilization of non-native protein-protein interactions of Coronavirus nucleocapsid proteins in antiviral drug design. J Med Chem [Internet]. 2020;63(6):3131–41. Available from: http://dx.doi.org/10.1021/acs.jmedchem.9b01913

1118. McCallum M, Walls AC, Bowen JE, Corti D, Veesler D. Structure-guided covalent stabilization of coronavirus spike glycoprotein trimers in the closed conformation. Nat Struct Mol Biol [Internet]. 2020;27(10):942–9. Available from: http://dx.doi.org/10.1038/s41594-020-0483-8

1119. Galasiti Kankanamalage AC, Kim Y, Damalanka VC, Rathnayake AD, Fehr AR, Mehzabeen N, et al. Structure-guided design of potent and permeable inhibitors of MERS coronavirus 3CL protease that utilize a piperidine moiety as a novel design element. Eur J Med Chem [Internet]. 2018;150:334–46. Available from: http://dx.doi.org/10.1016/j.ejmech.2018.03.004

1120. Akbayrak IY, Caglayan SI, Durdagi S, Kurgan L, Uversky VN, Ulver B, et al. Structures of MERS-CoV macro domain in aqueous solution with dynamics: Impacts of parallel tempering simulation techniques and CHARMM36m and AMBER99SB force field parameters. Proteins [Internet]. 2021;89(10):1289–99. Available from: http://dx.doi.org/10.1002/prot.26150

1121. Park Y-J, Walls AC, Wang Z, Sauer MM, Li W, Tortorici MA, et al. Structures of MERS-CoV spike glycoprotein in complex with sialoside attachment receptors. Nat Struct Mol Biol [Internet]. 2019;26(12):1151–7. Available from: http://dx.doi.org/10.1038/s41594-019-0334-7

1122. Kim I, Lee JE, Kim K-H, Lee S, Lee K, Mok JH. Successful treatment of suspected organizing pneumonia in a patient with Middle East respiratory syndrome coronavirus infection: a case report. J Thorac Dis [Internet]. 2016;8(10):E1190–4. Available from: http://dx.doi.org/10.21037/jtd.2016.09.26

1123. Ko J-H, Müller MA, Seok H, Park GE, Lee JY, Cho SY, et al. Suggested new breakpoints of anti-MERS-CoV antibody ELISA titers: performance analysis of serologic tests. Eur J Clin Microbiol Infect Dis [Internet]. 2017;36(11):2179–86. Available from: http://dx.doi.org/10.1007/s10096-017-3043-3

1124. Wang C, Xia S, Wang X, Li Y, Wang H, Xiang R, et al. Supercoiling structure-based design of a trimeric coiled-coil peptide with high potency against HIV-1 and human β-Coronavirus infection. J Med Chem [Internet]. 2022;65(4):2809–19. Available from: http://dx.doi.org/10.1021/acs.jmedchem.1c00258

1125. Kim MH, Kim HJ, Chang J. Superior immune responses induced by intranasal immunization with recombinant adenovirus-based vaccine expressing full-length Spike protein of Middle East respiratory syndrome coronavirus. PLoS One [Internet]. 2019;14(7):e0220196. Available from: http://dx.doi.org/10.1371/journal.pone.0220196

1126. van Doremalen N, Letko M, Fischer RJ, Bushmaker T, Schulz J, Yinda CK, et al. Surface‒aerosol stability and pathogenicity of diverse Middle East respiratory syndrome Coronavirus strains, 2012‒2018. Emerg Infect Dis [Internet]. 2021;27(12):3052–62. Available from: http://dx.doi.org/10.3201/eid2712.210344

1127. Bin Saeed AA, Abedi GR, Alzahrani AG, Salameh I, Abdirizak F, Alhakeem R, et al. Surveillance and testing for middle east respiratory syndrome Coronavirus, Saudi Arabia, April 2015–February 2016. Emerg Infect Dis [Internet]. 2017;23(4):682–5. Available from: http://dx.doi.org/10.3201/eid2304.161793

1128. Alzahrani A, Kujawski SA, Abedi GR, Tunkar S, Biggs HM, Alghawi N, et al. Surveillance and testing for Middle East respiratory syndrome Coronavirus, Saudi Arabia, March 2016-March 2019. Emerg Infect Dis [Internet]. 2020;26(7):1571–4. Available from: http://dx.doi.org/10.3201/eid2607.200437

1129. Kim C-J, Choi WS, Jung Y, Kiem S, Seol HY, Woo HJ, et al. Surveillance of the Middle East respiratory syndrome (MERS) coronavirus (CoV) infection in healthcare workers after contact with confirmed MERS patients: incidence and risk factors of MERS-CoV seropositivity. Clin Microbiol Infect [Internet]. 2016;22(10):880–6. Available from: http://dx.doi.org/10.1016/j.cmi.2016.07.017

1130. Lee M-K, Kim S, Kim M-N, Kweon OJ, Lim YK, Ki C-S, et al. Survey of clinical laboratory practices for 2015 Middle East respiratory syndrome Coronavirus outbreak in the Republic of Korea. Ann Lab Med [Internet]. 2016;36(2):154–61. Available from: http://dx.doi.org/10.3343/alm.2016.36.2.154

1131. Farag EAB, Nour M, El Idrissi A, Berrada J, Moustafa A, Mehmood M, et al. Survey on implementation of One Health approach for MERS-CoV preparedness and control in Gulf Cooperation Council and Middle East countries. Emerg Infect Dis [Internet]. 2019;25(3). Available from: http://dx.doi.org/10.3201/eid2503.171702

1132. Pyankov OV, Bodnev SA, Pyankova OG, Agranovski IE. Survival of aerosolized coronavirus in the ambient air. J Aerosol Sci [Internet]. 2018;115:158–63. Available from: http://dx.doi.org/10.1016/j.jaerosci.2017.09.009

1133. Kim Y-S, Aigerim A, Park U, Kim Y, Park H, Rhee J-Y, et al. Sustained responses of neutralizing antibodies against Middle East respiratory syndrome Coronavirus (MERS-CoV) in recovered patients and their therapeutic applicability. Clin Infect Dis [Internet]. 2021;73(3):e550–8. Available from: http://dx.doi.org/10.1093/cid/ciaa1345

1134. Kandeel M, Altaher A. Synonymous and biased Codon usage by MERS CoV papain-like and 3CL-proteases. Biol Pharm Bull [Internet]. 2017;40(7):1086–91. Available from: http://dx.doi.org/10.1248/bpb.b17-00168

1135. Yoon JH, Lee JY, Lee J, Shin YS, Jeon S, Kim DE, et al. Synthesis and biological evaluation of 3-acyl-2-phenylamino-1,4-dihydroquinolin-4(1H)-one derivatives as potential MERS-CoV inhibitors. Bioorg Med Chem Lett [Internet]. 2019;29(23):126727. Available from: http://dx.doi.org/10.1016/j.bmcl.2019.126727

1136. Elnosary ME, Aboelmagd HA, Habaka MA, Salem SR, El-Naggar ME. Synthesis of bee venom loaded chitosan nanoparticles for anti-MERS-COV and multi-drug resistance bacteria. Int J Biol Macromol [Internet]. 2023;224:871–80. Available from: http://dx.doi.org/10.1016/j.ijbiomac.2022.10.173

1137. Chowell G, Blumberg S, Simonsen L, Miller MA, Viboud C. Synthesizing data and models for the spread of MERS-CoV, 2013: key role of index cases and hospital transmission. Epidemics [Internet]. 2014;9:40–51. Available from: http://dx.doi.org/10.1016/j.epidem.2014.09.011

1138. Yoon M-K, Kim S-Y, Ko H-S, Lee M-S. System effectiveness of detection, brief intervention and refer to treatment for the people with post-traumatic emotional distress by MERS: a case report of community-based proactive intervention in South Korea. Int J Ment Health Syst [Internet]. 2016;10(1):51. Available from: http://dx.doi.org/10.1186/s13033-016-0083-5

1139. Ali MA, Shehata MM, Gomaa MR, Kandeil A, El-Shesheny R, Kayed AS, et al. Systematic, active surveillance for Middle East respiratory syndrome coronavirus in camels in Egypt. Emerg Microbes Infect [Internet]. 2017;6(1):e1. Available from: http://dx.doi.org/10.1038/emi.2016.130

1140. Guo X, Deng Y, Chen H, Lan J, Wang W, Zou X, et al. Systemic and mucosal immunity in mice elicited by a single immunization with human adenovirus type 5 or 41 vector-based vaccines carrying the spike protein of Middle East respiratory syndrome coronavirus. Immunology [Internet]. 2015;145(4):476–84. Available from: http://dx.doi.org/10.1111/imm.12462

1141. Lan J, Deng Y, Chen H, Lu G, Wang W, Guo X, et al. Tailoring subunit vaccine immunity with adjuvant combinations and delivery routes using the Middle East respiratory coronavirus (MERS-CoV) receptor-binding domain as an antigen. PLoS One [Internet]. 2014;9(11):e112602. Available from: http://dx.doi.org/10.1371/journal.pone.0112602

1142. Yuan S, Gao X, Tang K, Cai J-P, Hu M, Luo P, et al. Targeting papain-like protease for broad-spectrum coronavirus inhibition. Protein Cell [Internet]. 2022;13(12):940–53. Available from: http://dx.doi.org/10.1007/s13238-022-00909-3

1143. Firpo MR, Mastrodomenico V, Hawkins GM, Prot M, Levillayer L, Gallagher T, et al. Targeting polyamines inhibits Coronavirus infection by reducing cellular attachment and entry. ACS Infect Dis [Internet]. 2021;7(6):1423–32. Available from: http://dx.doi.org/10.1021/acsinfecdis.0c00491

1144. Flude BM, Nannetti G, Mitchell P, Compton N, Richards C, Heurich M, et al. Targeting the complement Serine protease MASP-2 as a therapeutic strategy for Coronavirus infections. Viruses [Internet]. 2021;13(2):312. Available from: http://dx.doi.org/10.3390/v13020312

1145. Kim J, Kim M, Kim D, Park S, Kang M, Baek K, et al. Targeting the interaction between Spike protein and nucleocapsid protein for suppression and detection of human Coronavirus OC43. Front Immunol [Internet]. 2022;13:835333. Available from: http://dx.doi.org/10.3389/fimmu.2022.835333

1146. St John SE, Tomar S, Stauffer SR, Mesecar AD. Targeting zoonotic viruses: Structure-based inhibition of the 3C-like protease from bat coronavirus HKU4--The likely reservoir host to the human coronavirus that causes Middle East Respiratory Syndrome (MERS). Bioorg Med Chem [Internet]. 2015;23(17):6036–48. Available from: http://dx.doi.org/10.1016/j.bmc.2015.06.039

1147. Mok CKP, Zhu A, Zhao J, Lau EHY, Wang J, Chen Z, et al. T-cell responses to MERS coronavirus infection in people with occupational exposure to dromedary camels in Nigeria: an observational cohort study. Lancet Infect Dis [Internet]. 2021;21(3):385–95. Available from: http://dx.doi.org/10.1016/S1473-3099(20)30599-5

1148. Baldassi F, Cenciarelli O, Malizia A, Gaudio P. Testing the identification effectiveness of an unknown outbreak of the Infectious Diseases Seeker (IDS) using and comparing the novel coronavirus disease (COVID-19) outbreak with the past SARS and MERS epidemics. J Infect Public Health [Internet]. 2021;14(1):123–30. Available from: http://dx.doi.org/10.1016/j.jiph.2020.11.014

1149. de Araujo TS, Barbosa GM, Sanches K, Azevedo JM, Dos Santos Cabral KM, Almeida MS, et al. The 1H, 15N, and 13C resonance assignments of the N-terminal domain of the nucleocapsid protein from the Middle East respiratory syndrome coronavirus. Biomol NMR Assign [Internet]. 2021;15(2):341–5. Available from: http://dx.doi.org/10.1007/s12104-021-10027-6

1150. Yang Y, Deng Y, Wen B, Wang H, Meng X, Lan J, et al. The amino acids 736-761 of the MERS-CoV spike protein induce neutralizing antibodies: implications for the development of vaccines and antiviral agents. Viral Immunol [Internet]. 2014;27(10):543–50. Available from: http://dx.doi.org/10.1089/vim.2014.0080

1151. Pu J, He X, Xu W, Wang C, Lan Q, Hua C, et al. The analogs of furanyl methylidene rhodanine exhibit broad-spectrum inhibitory and inactivating activities against enveloped viruses, including SARS-CoV-2 and its variants. Viruses [Internet]. 2022;14(3):489. Available from: http://dx.doi.org/10.3390/v14030489

1152. Martínez YA, Guo X, Portales-Pérez DP, Rivera G, Castañeda-Delgado JE, García-Pérez CA, et al. The analysis on the human protein domain targets and host-like interacting motifs for the MERS-CoV and SARS-CoV/CoV-2 infers the molecular mimicry of coronavirus. PLoS One [Internet]. 2021;16(2):e0246901. Available from: http://dx.doi.org/10.1371/journal.pone.0246901

1153. Gordon CJ, Tchesnokov EP, Feng JY, Porter DP, Götte M. The antiviral compound remdesivir potently inhibits RNA-dependent RNA polymerase from Middle East respiratory syndrome coronavirus. J Biol Chem [Internet]. 2020;295(15):4773–9. Available from: http://dx.doi.org/10.1074/jbc.AC120.013056

1154. Kim Y, Lee S, Chu C, Choe S, Hong S, Shin Y. The characteristics of Middle Eastern respiratory syndrome Coronavirus transmission dynamics in South Korea. Osong Public Health Res Perspect [Internet]. 2016;7(1):49–55. Available from: http://dx.doi.org/10.1016/j.phrp.2016.01.001

1155. Lee JY, Kim Y-J, Chung EH, Kim D-W, Jeong I, Kim Y, et al. The clinical and virological features of the first imported case causing MERS-CoV outbreak in South Korea, 2015. BMC Infect Dis [Internet]. 2017;17(1). Available from: http://dx.doi.org/10.1186/s12879-017-2576-5

1156. Grunewald ME, Fehr AR, Athmer J, Perlman S. The coronavirus nucleocapsid protein is ADP-ribosylated. Virology [Internet]. 2018;517:62–8. Available from: http://dx.doi.org/10.1016/j.virol.2017.11.020

1157. Gribble J, Stevens LJ, Agostini ML, Anderson-Daniels J, Chappell JD, Lu X, et al. The coronavirus proofreading exoribonuclease mediates extensive viral recombination. PLoS Pathog [Internet]. 2021;17(1):e1009226. Available from: http://dx.doi.org/10.1371/journal.ppat.1009226

1158. Perrier A, Bonnin A, Desmarets L, Danneels A, Goffard A, Rouillé Y, et al. The C-terminal domain of the MERS coronavirus M protein contains a trans-Golgi network localization signal. J Biol Chem [Internet]. 2019;294(39):14406–21. Available from: http://dx.doi.org/10.1074/jbc.RA119.008964

1159. Ku M, Han A, Lee K-H. The dynamics of cross-sector collaboration in centralized disaster governance: A network study of interorganizational collaborations during the MERS epidemic in South Korea. Int J Environ Res Public Health [Internet]. 2021;19(1):18. Available from: http://dx.doi.org/10.3390/ijerph19010018

1160. Joo H, Henry RE, Lee Y-K, Berro AD, Maskery BA. The effects of past SARS experience and proximity on declines in numbers of travelers to the Republic of Korea during the 2015 MERS outbreak: A retrospective study. Travel Med Infect Dis [Internet]. 2019;30:54–66. Available from: http://dx.doi.org/10.1016/j.tmaid.2019.05.009

1161. Jang K, Park N. The effects of repetitive information communication through multiple channels on prevention behavior during the 2015 MERS outbreak in South Korea. J Health Commun [Internet]. 2018;23(7):670–8. Available from: http://dx.doi.org/10.1080/10810730.2018.1501440

1162. Yoo W, Choi D-H, Park K. The effects of SNS communication: How expressing and receiving information predict MERS-preventive behavioral intentions in South Korea. Comput Human Behav [Internet]. 2016;62:34–43. Available from: http://dx.doi.org/10.1016/j.chb.2016.03.058

1163. Lee M, Ju Y, You M. The effects of social determinants on public health emergency preparedness mediated by health communication: The 2015 MERS outbreak in South Korea. Health Commun [Internet]. 2020;35(11):1396–406. Available from: http://dx.doi.org/10.1080/10410236.2019.1636342

1164. Nakagawa K, Narayanan K, Wada M, Popov VL, Cajimat M, Baric RS, et al. The endonucleolytic RNA cleavage function of nsp1 of middle East respiratory syndrome Coronavirus promotes the production of infectious virus particles in specific human cell lines. J Virol [Internet]. 2018;92(21). Available from: http://dx.doi.org/10.1128/JVI.01157-18

1165. Ogando NS, Zevenhoven-Dobbe JC, van der Meer Y, Bredenbeek PJ, Posthuma CC, Snijder EJ. The enzymatic activity of the nsp14 exoribonuclease is critical for replication of MERS-CoV and SARS-CoV-2. J Virol [Internet]. 2020;94(23). Available from: http://dx.doi.org/10.1128/JVI.01246-20

1166. Alsahafi AJ, Cheng AC. The epidemiology of Middle East respiratory syndrome coronavirus in the Kingdom of Saudi Arabia, 2012-2015. Int J Infect Dis [Internet]. 2016;45:1–4. Available from: http://dx.doi.org/10.1016/j.ijid.2016.02.004

1167. Im S-B, Baumann SL, Ahn M, Kim H, Youn B-H, Park M, et al. The experience of Korean nurses during the Middle East Respiratory Syndrome outbreak. Nurs Sci Q [Internet]. 2018;31(1):72–6. Available from: http://dx.doi.org/10.1177/0894318417741119

1168. Schoeman D, Cloete R, Fielding BC. The flexible, extended coil of the PDZ-binding motif of the three deadly human Coronavirus E proteins plays a role in pathogenicity. Viruses [Internet]. 2022;14(8):1707. Available from: http://dx.doi.org/10.3390/v14081707

1169. Coignard-Biehler H, Rapp C, Chapplain JM, Hoen B, Che D, Berthelot P, et al. The French Infectious Diseases Society’s readiness and response to epidemic or biological risk–the current situation following the Middle East respiratory syndrome coronavirus and Ebola virus disease alerts. Med Mal Infect [Internet]. 2018;48(2):95–102. Available from: http://dx.doi.org/10.1016/j.medmal.2017.10.002

1170. Salamatbakhsh M, Mobaraki K, Sadeghimohammadi S, Ahmadzadeh J. The global burden of premature mortality due to the Middle East respiratory syndrome (MERS) using standard expected years of life lost, 2012 to 2019. BMC Public Health [Internet]. 2019;19(1):1523. Available from: http://dx.doi.org/10.1186/s12889-019-7899-2

1171. Forni D, Filippi G, Cagliani R, De Gioia L, Pozzoli U, Al-Daghri N, et al. The heptad repeat region is a major selection target in MERS-CoV and related coronaviruses. Sci Rep [Internet]. 2015;5(1):14480. Available from: http://dx.doi.org/10.1038/srep14480

1172. Chefer S, Seidel J, Cockrell AS, Yount B, Solomon J, Hagen KR, et al. The human sodium iodide symporter as a reporter gene for studying Middle East respiratory syndrome Coronavirus pathogenesis. mSphere [Internet]. 2018;3(6). Available from: http://dx.doi.org/10.1128/mSphere.00540-18

1173. Lee A, Cho J. The impact of epidemics on labor market: identifying victims of the Middle East Respiratory Syndrome in the Korean labor market. Int J Equity Health [Internet]. 2016;15(1). Available from: http://dx.doi.org/10.1186/s12939-016-0483-9

1174. Choi D-H, Yoo W, Noh G-Y, Park K. The impact of social media on risk perceptions during the MERS outbreak in South Korea. Comput Human Behav [Internet]. 2017;72:422–31. Available from: http://dx.doi.org/10.1016/j.chb.2017.03.004

1175. Jeon YJ, Gil CH, Jo A, Won J, Kim S, Kim HJ. The influence of interferon-lambda on restricting Middle East Respiratory Syndrome Coronavirus replication in the respiratory epithelium. Antiviral Res [Internet]. 2020;180(104860):104860. Available from: http://dx.doi.org/10.1016/j.antiviral.2020.104860

1176. Zhu L, Gao T, Fu Y, Han X, Yue J, Liu Y, et al. The MERS-CoV N protein regulates host cytokinesis and protein translation via interaction with EF1A. Front Microbiol [Internet]. 2021;12:551602. Available from: http://dx.doi.org/10.3389/fmicb.2021.551602

1177. de Wit E, Prescott J, Baseler L, Bushmaker T, Thomas T, Lackemeyer MG, et al. The Middle East respiratory syndrome coronavirus (MERS-CoV) does not replicate in Syrian hamsters. PLoS One [Internet]. 2013;8(7):e69127. Available from: http://dx.doi.org/10.1371/journal.pone.0069127

1178. Hemida MG, Ali AM, Alnaeem A. The Middle East respiratory syndrome coronavirus (MERS-CoV) nucleic acids detected in the saliva and conjunctiva of some naturally infected dromedary camels in Saudi Arabia -2019. Zoonoses Public Health [Internet]. 2021;68(4):353–7. Available from: http://dx.doi.org/10.1111/zph.12816

1179. Alabdali A, Almakhalas K, Alhusain F, Albaiz S, Almutairi K, Aljerian N. The middle East Respiratory Syndrome Coronavirus (MERS-CoV) outbreak at King Abdul-Aziz Medical City-Riyadh from Emergency Medical Services perspective. Prehosp Disaster Med [Internet]. 2020;35(4):457–61. Available from: http://dx.doi.org/10.1017/S1049023X20000709

1180. Hemida MG, Ali M, Alhammadi M, Alnaeem A. The Middle East respiratory syndrome coronavirus in the breath of some infected dromedary camels (Camelus dromedarius). Epidemiol Infect [Internet]. 2020;148(e247):e247. Available from: http://dx.doi.org/10.1017/S0950268820002459

1181. Rona G, Zeke A, Miwatani-Minter B, de Vries M, Kaur R, Schinlever A, et al. The NSP14/NSP10 RNA repair complex as a Pan-coronavirus therapeutic target. Cell Death Differ [Internet]. 2022;29(2):285–92. Available from: http://dx.doi.org/10.1038/s41418-021-00900-1

1182. Li Y, Jin Y, Kuang L, Luo Z, Li F, Sun J, et al. The N-terminal region of middle East respiratory syndrome Coronavirus accessory protein 8b is essential for enhanced virulence of an attenuated Murine Coronavirus. J Virol [Internet]. 2022;96(3):e0184221. Available from: http://dx.doi.org/10.1128/JVI.01842-21

1183. Matthews KL, Coleman CM, van der Meer Y, Snijder EJ, Frieman MB. The ORF4b-encoded accessory proteins of Middle East respiratory syndrome coronavirus and two related bat coronaviruses localize to the nucleus and inhibit innate immune signalling. J Gen Virol [Internet]. 2014;95(Pt 4):874–82. Available from: http://dx.doi.org/10.1099/vir.0.062059-0

1184. Alghamdi IG, Hussain II, Almalki SS, Alghamdi MS, Alghamdi MM, El-Sheemy MA. The pattern of Middle East respiratory syndrome coronavirus in Saudi Arabia: a descriptive epidemiological analysis of data from the Saudi Ministry of Health. Int J Gen Med [Internet]. 2014;7:417–23. Available from: http://dx.doi.org/10.2147/IJGM.S67061

1185. Aldohyan M, Al-Rawashdeh N, Sakr FM, Rahman S, Alfarhan AI, Salam M. The perceived effectiveness of MERS-CoV educational programs and knowledge transfer among primary healthcare workers: a cross-sectional survey. BMC Infect Dis [Internet]. 2019;19(1):273. Available from: http://dx.doi.org/10.1186/s12879-019-3898-2

1186. Chen X, Adam DC, Chughtai AA, Stelzer-Braid S, Scotch M, MacIntyre CR. The phylogeography of MERS-CoV in hospital outbreak-associated cases compared to sporadic cases in Saudi Arabia. Viruses [Internet]. 2020;12(5):540. Available from: http://dx.doi.org/10.3390/v12050540

1187. George PJ, Tai W, Du L, Lustigman S. The potency of an anti-MERS Coronavirus subunit vaccine depends on a unique combinatorial adjuvant formulation. Vaccines (Basel) [Internet]. 2020;8(2):251. Available from: http://dx.doi.org/10.3390/vaccines8020251

1188. Huang J, Teoh JY-C, Wong SH, Wong MCS. The potential impact of previous exposure to SARS or MERS on control of the COVID-19 pandemic. Eur J Epidemiol [Internet]. 2020;35(11):1099–103. Available from: http://dx.doi.org/10.1007/s10654-020-00674-9

1189. Alsharif KF, Alzahrani AB, Alharbi AO, Algregri TO, Almuafa BH, Alsulami MO, et al. The prevalence of MERS-CoV among military personnel and their families: A single-center study. J Med Virol [Internet]. 2021;93(5):2815–9. Available from: http://dx.doi.org/10.1002/jmv.26642

1190. Harcourt JL, Rudoler N, Tamin A, Leshem E, Rasis M, Giladi M, et al. The prevalence of Middle East respiratory syndrome coronavirus (MERS-CoV) antibodies in dromedary camels in Israel. Zoonoses Public Health [Internet]. 2018;65(6):749–54. Available from: http://dx.doi.org/10.1111/zph.12482

1191. Kasem S, Qasim I, Al-Doweriej A, Hashim O, Alkarar A, Abu-Obeida A, et al. The prevalence of Middle East respiratory Syndrome coronavirus (MERS-CoV) infection in livestock and temporal relation to locations and seasons. J Infect Public Health [Internet]. 2018;11(6):884–8. Available from: http://dx.doi.org/10.1016/j.jiph.2018.01.004

1192. Karagulov AI, Argimbayeva TU, Omarova ZD, Tulendibayev AB, Dushayeva LZ, Svotina MA, et al. The prevalence of viral pathogens among bats in Kazakhstan. Viruses [Internet]. 2022;14(12):2743. Available from: http://dx.doi.org/10.3390/v14122743

1193. Rahmatizadeh S, Valizadeh-Haghi S. The readability of online health information on Middle East respiratory syndrome Coronavirus disease. Journal of Cellular & Molecular Anesthesia [Internet]. 2021 [cited 2023 Mar 22];6(2):154–63. Available from: https://journals.sbmu.ac.ir/jcma/article/view/31749

1194. Kim JI, Kim Y-J, Lemey P, Lee I, Park S, Bae J-Y, et al. The recent ancestry of Middle East respiratory syndrome coronavirus in Korea has been shaped by recombination. Sci Rep [Internet]. 2016;6(1):18825. Available from: http://dx.doi.org/10.1038/srep18825

1195. Mou H, Raj VS, van Kuppeveld FJM, Rottier PJM, Haagmans BL, Bosch BJ. The receptor binding domain of the new Middle East respiratory syndrome coronavirus maps to a 231-residue region in the spike protein that efficiently elicits neutralizing antibodies. J Virol [Internet]. 2013;87(16):9379–83. Available from: http://dx.doi.org/10.1128/JVI.01277-13

1196. Jiaming L, Yanfeng Y, Yao D, Yawei H, Linlin B, Baoying H, et al. The recombinant N-terminal domain of spike proteins is a potential vaccine against Middle East respiratory syndrome coronavirus (MERS-CoV) infection. Vaccine [Internet]. 2017;35(1):10–8. Available from: http://dx.doi.org/10.1016/j.vaccine.2016.11.064

1197. Ahmadzadeh J, Mobaraki K, Mousavi SJ, Aghazadeh-Attari J, Mirza-Aghazadeh-Attari M, Mohebbi I. The risk factors associated with MERS-CoV patient fatality: A global survey. Diagn Microbiol Infect Dis [Internet]. 2020;96(3):114876. Available from: http://dx.doi.org/10.1016/j.diagmicrobio.2019.114876

1198. Park H-J, Lee BJ. The role of social work for foreign residents in an epidemic: The MERS crisis in the republic of Korea. Soc Work Public Health [Internet]. 2016;31(7):656–64. Available from: http://dx.doi.org/10.1080/19371918.2016.1160352

1199. Kucharski AJ, Althaus CL. The role of superspreading in Middle East respiratory syndrome coronavirus (MERS-CoV) transmission. Euro Surveill [Internet]. 2015;20(25):14–8. Available from: http://dx.doi.org/10.2807/1560-7917.es2015.20.25.21167

1200. Laporte M, Raeymaekers V, Van Berwaer R, Vandeput J, Marchand-Casas I, Thibaut H-J, et al. The SARS-CoV-2 and other human coronavirus spike proteins are fine-tuned towards temperature and proteases of the human airways. PLoS Pathog [Internet]. 2021;17(4):e1009500. Available from: http://dx.doi.org/10.1371/journal.ppat.1009500

1201. Örd M, Faustova I, Loog M. The sequence at Spike S1/S2 site enables cleavage by furin and phospho-regulation in SARS-CoV2 but not in SARS-CoV1 or MERS-CoV. Sci Rep [Internet]. 2020;10(1):16944. Available from: http://dx.doi.org/10.1038/s41598-020-74101-0

1202. Jang T-H, Park W-J, Lee H, Woo H-M, Lee S-Y, Kim K-C, et al. The structure of a novel antibody against the spike protein inhibits Middle East respiratory syndrome coronavirus infections. Sci Rep [Internet]. 2022;12(1):1260. Available from: http://dx.doi.org/10.1038/s41598-022-05318-4

1203. Earnest JT, Hantak MP, Li K, McCray PB Jr, Perlman S, Gallagher T. The tetraspanin CD9 facilitates MERS-coronavirus entry by scaffolding host cell receptors and proteases. PLoS Pathog [Internet]. 2017;13(7):e1006546. Available from: http://dx.doi.org/10.1371/journal.ppat.1006546

1204. Rui J, Wang Q, Lv J, Zhao B, Hu Q, Du H, et al. The transmission dynamics of Middle East Respiratory Syndrome coronavirus. Travel Med Infect Dis [Internet]. 2022;45(102243):102243. Available from: http://dx.doi.org/10.1016/j.tmaid.2021.102243

1205. Joseph J, Karthika T, Das VRA, Raj VS. The use of pseudotyped coronaviruses for the screening of entry inhibitors: Green tea extract inhibits the entry of SARS-CoV-1, MERSCoV, and SARS-CoV-2 by blocking receptor-spike interaction. Curr Pharm Biotechnol [Internet]. 2022;23(8):1118–29. Available from: http://dx.doi.org/10.2174/1389201022666210810111716

1206. Fauver JR, Gendernalik A, Weger-Lucarelli J, Grubaugh ND, Brackney DE, Foy BD, et al. The use of xenosurveillance to detect human bacteria, parasites, and viruses in mosquito bloodmeals. Am J Trop Med Hyg [Internet]. 2017;97(2):324–9. Available from: http://dx.doi.org/10.4269/ajtmh.17-0063

1207. Cheng K-W, Cheng S-C, Chen W-Y, Lin M-H, Chuang S-J, Cheng I-H, et al. Thiopurine analogs and mycophenolic acid synergistically inhibit the papain-like protease of Middle East respiratory syndrome coronavirus. Antiviral Res [Internet]. 2015;115:9–16. Available from: http://dx.doi.org/10.1016/j.antiviral.2014.12.011

1208. Ekins S, Madrid PB. Tilorone, a broad-spectrum antiviral for emerging viruses. Antimicrob Agents Chemother [Internet]. 2020;64(5). Available from: http://dx.doi.org/10.1128/AAC.00440-20

1209. Meyer B, Juhasz J, Barua R, Das Gupta A, Hakimuddin F, Corman VM, et al. Time course of MERS-CoV infection and immunity in dromedary camels. Emerg Infect Dis [Internet]. 2016;22(12):2171–3. Available from: http://dx.doi.org/10.3201/eid2212.160382

1210. Widagdo W, Begeman L, Schipper D, van Run PR, Cunningham AA, Kley N, et al. Tissue distribution of the MERS-Coronavirus receptor in bats. Sci Rep [Internet]. 2017;7(1). Available from: http://dx.doi.org/10.1038/s41598-017-01290-6

1211. Zmora P, Hoffmann M, Kollmus H, Moldenhauer A-S, Danov O, Braun A, et al. TMPRSS11A activates the influenza A virus hemagglutinin and the MERS coronavirus spike protein and is insensitive against blockade by HAI-1. J Biol Chem [Internet]. 2018;293(36):13863–73. Available from: http://dx.doi.org/10.1074/jbc.ra118.001273

1212. Iwata-Yoshikawa N, Okamura T, Shimizu Y, Hasegawa H, Takeda M, Nagata N. TMPRSS2 contributes to virus spread and immunopathology in the airways of Murine models after Coronavirus infection. J Virol [Internet]. 2019;93(6). Available from: http://dx.doi.org/10.1128/JVI.01815-18

1213. Widjaja I, Wang C, van Haperen R, Gutiérrez-Álvarez J, van Dieren B, Okba NMA, et al. Towards a solution to MERS: protective human monoclonal antibodies targeting different domains and functions of the MERS-coronavirus spike glycoprotein. Emerg Microbes Infect [Internet]. 2019;8(1):516–30. Available from: http://dx.doi.org/10.1080/22221751.2019.1597644

1214. Regan JJ, Jungerman MR, Lippold SA, Washburn F, Roland E, Objio T, et al. Tracing airline travelers for a public health investigation: Middle East Respiratory Syndrome Coronavirus (MERS-CoV) infection in the United States, 2014: Middle East Respiratory Syndrome Coronavirus (MERS-CoV) infection in the United States, 2014. Public Health Rep [Internet]. 2016;131(4):552–9. Available from: http://dx.doi.org/10.1177/0033354916662213

1215. Hala S, Ribeca P, Aljami HA, Alsagaby SA, Qasim I, Gilbert SC, et al. Transcriptomic profiling of dromedary camels immunised with a MERS vaccine candidate. Vet Sci [Internet]. 2021;8(8):156. Available from: http://dx.doi.org/10.3390/vetsci8080156

1216. Muth D, Meyer B, Niemeyer D, Schroeder S, Osterrieder N, Müller MA, et al. Transgene expression in the genome of Middle East respiratory syndrome coronavirus based on a novel reverse genetics system utilizing Red-mediated recombination cloning. J Gen Virol [Internet]. 2017;98(10):2461–9. Available from: http://dx.doi.org/10.1099/jgv.0.000919

1217. Van Kerkhove MD, Alaswad S, Assiri A, Perera RAPM, Peiris M, El Bushra HE, et al. Transmissibility of MERS-CoV infection in closed setting, Riyadh, Saudi Arabia, 2015. Emerg Infect Dis [Internet]. 2019;25(10):1802–9. Available from: http://dx.doi.org/10.3201/eid2510.190130

1218. Cotten M, Watson SJ, Kellam P, Al-Rabeeah AA, Makhdoom HQ, Assiri A, et al. Transmission and evolution of the Middle East respiratory syndrome coronavirus in Saudi Arabia: a descriptive genomic study. Lancet [Internet]. 2013;382(9909):1993–2002. Available from: http://dx.doi.org/10.1016/s0140-6736(13)61887-5

1219. Drosten C, Meyer B, Müller MA, Corman VM, Al-Masri M, Hossain R, et al. Transmission of MERS-coronavirus in household contacts. N Engl J Med [Internet]. 2014;371(9):828–35. Available from: http://dx.doi.org/10.1056/NEJMoa1405858

1220. Hunter JC, Nguyen D, Aden B, Al Bandar Z, Al Dhaheri W, Abu Elkheir K, et al. Transmission of middle East respiratory syndrome Coronavirus infections in healthcare settings, Abu Dhabi. Emerg Infect Dis [Internet]. 2016;22(4):647–56. Available from: http://dx.doi.org/10.3201/eid2204.151615

1221. Fanoy EB, van der Sande MA, Kraaij-Dirkzwager M, Dirksen K, Jonges M, van der Hoek W, et al. Travel-related MERS-CoV cases: an assessment of exposures and risk factors in a group of Dutch travellers returning from the Kingdom of Saudi Arabia, May 2014. Emerg Themes Epidemiol [Internet]. 2014;11(1):16. Available from: http://dx.doi.org/10.1186/1742-7622-11-16

1222. Arabi YM, Asiri AY, Assiri AM, Aziz Jokhdar HA, Alothman A, Balkhy HH, et al. Treatment of Middle East respiratory syndrome with a combination of lopinavir/ritonavir and interferon-β1b (MIRACLE trial): statistical analysis plan for a recursive two-stage group sequential randomized controlled trial. Trials [Internet]. 2020;21(1):8. Available from: http://dx.doi.org/10.1186/s13063-019-3846-x

1223. Al Ghamdi M, Alghamdi KM, Ghandoora Y, Alzahrani A, Salah F, Alsulami A, et al. Treatment outcomes for patients with Middle Eastern Respiratory Syndrome Coronavirus (MERS CoV) infection at a coronavirus referral center in the Kingdom of Saudi Arabia. BMC Infect Dis [Internet]. 2016;16(1):174. Available from: http://dx.doi.org/10.1186/s12879-016-1492-4

1224. Falzarano D, de Wit E, Rasmussen AL, Feldmann F, Okumura A, Scott DP, et al. Treatment with interferon-α2b and ribavirin improves outcome in MERS-CoV–infected rhesus macaques. Nat Med [Internet]. 2013;19(10):1313–7. Available from: http://dx.doi.org/10.1038/nm.3362

1225. Chan JF-W, Yao Y, Yeung M-L, Deng W, Bao L, Jia L, et al. Treatment with lopinavir/ritonavir or interferon-β1b improves outcome of MERS-CoV infection in a nonhuman primate model of common marmoset. J Infect Dis [Internet]. 2015;212(12):1904–13. Available from: http://dx.doi.org/10.1093/infdis/jiv392

1226. Chan RWY, Hemida MG, Kayali G, Chu DKW, Poon LLM, Alnaeem A, et al. Tropism and replication of Middle East respiratory syndrome coronavirus from dromedary camels in the human respiratory tract: an in-vitro and ex-vivo study. Lancet Respir Med [Internet]. 2014;2(10):813–22. Available from: http://dx.doi.org/10.1016/S2213-2600(14)70158-4

1227. Menachery VD, Dinnon KH 3rd, Yount BL Jr, McAnarney ET, Gralinski LE, Hale A, et al. Trypsin treatment unlocks barrier for zoonotic bat Coronavirus infection. J Virol [Internet]. 2020;94(5). Available from: http://dx.doi.org/10.1128/JVI.01774-19

1228. Lin L, McCloud RF, Bigman CA, Viswanath K. Tuning in and catching on? Examining the relationship between pandemic communication and awareness and knowledge of MERS in the USA. J Public Health (Oxf) [Internet]. 2016;fdw028. Available from: http://dx.doi.org/10.1093/pubmed/fdw028

1229. Fung IC-H, Zeng J, Chan C-H, Liang H, Yin J, Liu Z, et al. Twitter and Middle East respiratory syndrome, South Korea, 2015: A multi-lingual study. Infect Dis Health [Internet]. 2018;23(1):10–6. Available from: http://dx.doi.org/10.1016/j.idh.2017.08.005

1230. Xie Q, Cao Y, Su J, Wu J, Wu X, Wan C, et al. Two deletion variants of Middle East respiratory syndrome coronavirus found in a patient with characteristic symptoms. Arch Virol [Internet]. 2017;162(8):2445–9. Available from: http://dx.doi.org/10.1007/s00705-017-3361-x

1231. Te N, Rodon J, Ballester M, Pérez M, Pailler-García L, Segalés J, et al. Type I and III IFNs produced by the nasal epithelia and dimmed inflammation are features of alpacas resolving MERS-CoV infection. PLoS Pathog [Internet]. 2021;17(5):e1009229. Available from: http://dx.doi.org/10.1371/journal.ppat.1009229

1232. Nemr WA, Radwan NK. Typing of alpha and beta coronaviruses by DNA barcoding of NSP12 gene. J Med Virol [Internet]. 2022;94(5):1926–34. Available from: http://dx.doi.org/10.1002/jmv.27550

1233. Zhou Y, Zheng R, Liu D, Liu S, Disoma C, Li S, et al. UBR5 acts as an antiviral host factor against MERS-CoV via promoting ubiquitination and degradation of ORF4b. J Virol [Internet]. 2022;96(17):e0074122. Available from: http://dx.doi.org/10.1128/jvi.00741-22

1234. Niu P, Zhang S, Zhou P, Huang B, Deng Y, Qin K, et al. Ultrapotent human neutralizing antibody repertoires against middle east respiratory syndrome Coronavirus from a recovered patient. J Infect Dis [Internet]. 2018;218(8):1249–60. Available from: http://dx.doi.org/10.1093/infdis/jiy311

1235. Kazuya S, Nao N, Matsuyama S, Kageyama T. Ultra-rapid real-time RT-PCR method for detecting middle East respiratory syndrome Coronavirus using a mobile PCR device, PCR1100. Jpn J Infect Dis [Internet]. 2020;73(3):181–6. Available from: http://dx.doi.org/10.7883/yoken.JJID.2019.400

1236. Yang C-W, Shi Z-L. Uncovering potential host proteins and pathways that may interact with eukaryotic short linear motifs in viral proteins of MERS, SARS and SARS2 coronaviruses that infect humans. PLoS One [Internet]. 2021;16(2):e0246150. Available from: http://dx.doi.org/10.1371/journal.pone.0246150

1237. Da’ar OB, Ahmed AE. Underlying trend, seasonality, prediction, forecasting and the contribution of risk factors: an analysis of globally reported cases of Middle East Respiratory Syndrome Coronavirus. Epidemiol Infect [Internet]. 2018;146(11):1343–9. Available from: http://dx.doi.org/10.1017/S0950268818001541

1238. Walls AC, Xiong X, Park Y-J, Tortorici MA, Snijder J, Quispe J, et al. Unexpected receptor functional mimicry elucidates activation of Coronavirus fusion. Cell [Internet]. 2019;176(5):1026-1039.e15. Available from: http://dx.doi.org/10.1016/j.cell.2018.12.028

1239. Tan Y, Schneider T, Shukla PK, Chandrasekharan MB, Aravind L, Zhang D. Unification and extensive diversification of M/Orf3-related ion channel proteins in coronaviruses and other nidoviruses. Virus Evol [Internet]. 2021;7(1):veab014. Available from: http://dx.doi.org/10.1093/ve/veab014

1240. Trigueiro-Louro J, Correia V, Figueiredo-Nunes I, Gíria M, Rebelo-de-Andrade H. Unlocking COVID therapeutic targets: A structure-based rationale against SARS-CoV-2, SARS-CoV and MERS-CoV Spike. Comput Struct Biotechnol J [Internet]. 2020;18:2117–31. Available from: http://dx.doi.org/10.1016/j.csbj.2020.07.017

1241. Cauchemez S, Nouvellet P, Cori A, Jombart T, Garske T, Clapham H, et al. Unraveling the drivers of MERS-CoV transmission. Proc Natl Acad Sci U S A [Internet]. 2016;113(32):9081–6. Available from: http://dx.doi.org/10.1073/pnas.1519235113

1242. Amer H, Alqahtani AS, Alzoman H, Aljerian N, Memish ZA. Unusual presentation of Middle East respiratory syndrome coronavirus leading to a large outbreak in Riyadh during 2017. Am J Infect Control [Internet]. 2018;46(9):1022–5. Available from: http://dx.doi.org/10.1016/j.ajic.2018.02.023

1243. Chen X, Chughtai AA, Macintyre R. Use of a modified Grunow-Finke tool to assess the origin of MERS-CoV in Saudi Arabia. Int J Infect Dis [Internet]. 2020;101:253–4. Available from: http://dx.doi.org/10.1016/j.ijid.2020.11.098

1244. Hoteit R, Shammaa D, Mahfouz R. Use of the Human Coronavirus 2012 (MERS) GeneSig kit for MERS-CoV detection. Gene Rep [Internet]. 2016;4:67–9. Available from: http://dx.doi.org/10.1016/j.genrep.2016.04.004

1245. Qiang X-L, Xu P, Fang G, Liu W-B, Kou Z. Using the spike protein feature to predict infection risk and monitor the evolutionary dynamic of coronavirus. Infect Dis Poverty [Internet]. 2020;9(1):33. Available from: http://dx.doi.org/10.1186/s40249-020-00649-8

1246. Kim D-W, Kim Y-J, Park SH, Yun M-R, Yang J-S, Kang HJ, et al. Variations in spike glycoprotein gene of MERS-CoV, South Korea, 2015. Emerg Infect Dis [Internet]. 2016;22(1):100–4. Available from: http://dx.doi.org/10.3201/eid2201.151055

1247. Yu HC, Mui KW, Wong LT, Chu HS. Ventilation of general hospital wards for mitigating infection risks of three kinds of viruses including Middle East respiratory syndrome coronavirus. Indoor Built Environ [Internet]. 2017;26(4):514–27. Available from: http://dx.doi.org/10.1177/1420326x16631596

1248. Hecht L-S, Jurado-Jimenez A, Hess M, Halas HE, Bochenek G, Mohammed H, et al. Verification and diagnostic evaluation of the RealStar® Middle East respiratory syndrome coronavirus (N gene) reverse transcription-PCR kit 1.0. Future Microbiol [Internet]. 2019;14(11):941–8. Available from: http://dx.doi.org/10.2217/fmb-2019-0067

1249. Kim SY, Park SJ, Cho SY, Cha R-H, Jee H-G, Kim G, et al. Viral RNA in blood as indicator of severe outcome in middle east respiratory syndrome Coronavirus infection. Emerg Infect Dis [Internet]. 2016;22(10):1813–6. Available from: http://dx.doi.org/10.3201/eid2210.160218

1250. Corman VM, Albarrak AM, Omrani AS, Albarrak MM, Farah ME, Almasri M, et al. Viral shedding and antibody response in 37 patients with Middle East respiratory syndrome Coronavirus infection. Clin Infect Dis [Internet]. 2016;62(4):477–83. Available from: http://dx.doi.org/10.1093/cid/civ951

1251. Song JY, Cheong HJ, Choi MJ, Jeon JH, Kang SH, Jeong EJ, et al. Viral shedding and environmental cleaning in middle east respiratory syndrome Coronavirus infection. Infect Chemother [Internet]. 2015;47(4):252–5. Available from: http://dx.doi.org/10.3947/ic.2015.47.4.252

1252. Spanakis N, Tsiodras S, Haagmans BL, Raj VS, Pontikis K, Koutsoukou A, et al. Virological and serological analysis of a recent Middle East respiratory syndrome coronavirus infection case on a triple combination antiviral regimen. Int J Antimicrob Agents [Internet]. 2014;44(6):528–32. Available from: http://dx.doi.org/10.1016/j.ijantimicag.2014.07.026

1253. Siddharta A, Pfaender S, Vielle NJ, Dijkman R, Friesland M, Becker B, et al. Virucidal activity of World Health Organization-recommended formulations against enveloped viruses, including Zika, Ebola, and emerging coronaviruses. J Infect Dis [Internet]. 2017;215(6):902–6. Available from: http://dx.doi.org/10.1093/infdis/jix046

1254. Wiederkehr MA, Qi W, Schoenbaechler K, Fraefel C, Kubacki J. Virus diversity, abundance, and evolution in three different bat colonies in Switzerland. Viruses [Internet]. 2022;14(9):1911. Available from: http://dx.doi.org/10.3390/v14091911

1255. Yuan S, Chu H, Huang J, Zhao X, Ye Z-W, Lai P-M, et al. Viruses harness YxxØ motif to interact with host AP2M1 for replication: A vulnerable broad-spectrum antiviral target. Sci Adv [Internet]. 2020;6(35):eaba7910. Available from: http://dx.doi.org/10.1126/sciadv.aba7910

1256. Fedorov V, Kholina E, Khruschev S, Kovalenko I, Rubin A, Strakhovskaya M. What binds cationic photosensitizers better: Brownian dynamics reveals key interaction sites on spike proteins of SARS-CoV, MERS-CoV, and SARS-CoV-2. Viruses [Internet]. 2021;13(8):1615. Available from: http://dx.doi.org/10.3390/v13081615

1257. Jang K, Baek YM. When information from public health officials is untrustworthy: The use of online news, interpersonal networks, and social media during the MERS outbreak in South Korea. Health Commun [Internet]. 2019;34(9):991–8. Available from: http://dx.doi.org/10.1080/10410236.2018.1449552

1258. Khan S, El Morabet R, Khan RA, Bindajam A, Alqadhi S, Alsubih M, et al. Where we missed? Middle East Respiratory Syndrome (MERS-CoV) epidemiology in Saudi Arabia; 2012-2019. Sci Total Environ [Internet]. 2020;747(141369):141369. Available from: http://dx.doi.org/10.1016/j.scitotenv.2020.141369

1259. Coleman CM, Matthews KL, Goicochea L, Frieman MB. Wild-type and innate immune-deficient mice are not susceptible to the Middle East respiratory syndrome coronavirus. J Gen Virol [Internet]. 2014;95(Pt 2):408–12. Available from: http://dx.doi.org/10.1099/vir.0.060640-0

1260. Ro J-S, Lee J-S, Kang S-C, Jung H-M. Worry experienced during the 2015 Middle East Respiratory Syndrome (MERS) pandemic in Korea. PLoS One [Internet]. 2017;12(3):e0173234. Available from: http://dx.doi.org/10.1371/journal.pone.0173234

1261. Clasman JR, Báez-Santos YM, Mettelman RC, O’Brien A, Baker SC, Mesecar AD. X-ray Structure and Enzymatic Activity Profile of a Core Papain-like Protease of MERS Coronavirus with utility for structure-based drug design. Sci Rep [Internet]. 2017;7:40292. Available from: http://dx.doi.org/10.1038/srep40292

1262. Alhomoud F, Alhomoud F. “Your Health Essential for Your Hajj”: Muslim pilgrims’ knowledge, attitudes and practices regarding Middle East respiratory syndrome coronavirus (MERS-CoV) during Hajj season. J Infect Chemother [Internet]. 2017;23(5):286–92. Available from: http://dx.doi.org/10.1016/j.jiac.2017.01.006

1263. Paden CR, Yusof MFBM, Al Hammadi ZM, Queen K, Tao Y, Eltahir YM, et al. Zoonotic origin and transmission of Middle East respiratory syndrome coronavirus in the UAE. Zoonoses Public Health [Internet]. 2018;65(3):322–33. Available from: http://dx.doi.org/10.1111/zph.12435

1264. Zhang L, Lin D, Kusov Y, Nian Y, Ma Q, Wang J, et al. Α-ketoamides as broad-spectrum inhibitors of Coronavirus and Enterovirus replication: Structure-based design, synthesis, and activity assessment. J Med Chem [Internet]. 2020;63(9):4562–78. Available from: http://dx.doi.org/10.1021/acs.jmedchem.9b01828
